# The Evolution of Primary Aldosteronism and the Role of Norrin

**DOI:** 10.1101/2025.06.05.25329066

**Authors:** Stéfanie Parisien-La Salle, Mahyar Heydarpour, Cheng-Hsuan Tsai, Jenifer M Brown, Andrew J Newman, Sanan Mahrokhian, Isabelle Hanna, Brooke Honzel, Laura C Tsai, Sushrut Waikar, Kosuke Inoue, Maria-Christina Zennaro, Richard Auchus, Adina F. Turcu, Jonathan S Williams, Barry Sacks, Marwan Moussa, Anand Vaidya

**Author notes:** **Corresponding Author**: Anand Vaidya, MD MMSc; Center for Adrenal Disorders, Mass General Brigham, Harvard Medical School, Boston, MA, 02115.

## Abstract

Primary aldosteronism (PA) is renin-independent aldosteronism that causes hypertension and cardiovascular disease. In this human physiology study, deep-phenotyping maneuvers were conducted to characterize the spectrum of Subclinical PA (SubPA) in normotensive people, along with measurement of the proteomic evolution of PA as it spanned across the continuum of SubPA and in people with clinically overt PA. Differential abundance and dose-dependent aldosterone trend analyses identified a progressive proteomic signature that paralleled the severity of renin-independent aldosteronism that was enriched for cardiovascular disease pathways, including inflammation, redox imbalance, and vascular remodeling. Following multiple *in vivo* testing maneuvers to modulate aldosterone physiology, Norrin, a Wnt/β-catenin ligand previously identified as an inheritable risk locus for PA, was robustly validated. Candidate-gene validations in a separate cohort of participants that had undergone extensive *in vivo* aldosterone phenotyping identified *NDP* variants that were reproducibly associated with multiple physiologic aldosterone traits that were supported by adrenal eQTL data. These findings demonstrate that the clinical spectrum of PA originates in normotensive people and is characterized by progressive proteomic signatures involved in the pathogenesis of cardiovascular disease. Moreover, these convergent proteomic, physiologic, and genetic findings establish Norrin as a novel mediator in the pathogenesis of PA.

## INTRODUCTION

Primary aldosteronism (PA) was once considered to be a rare hormonal cause of hypertension; however, recent human physiology and epidemiology studies indicate that PA is a common disorder in people with hypertension (1, 2). Identification of pathogenic somatic variants that cause PA (3, 4), along with the discovery that adrenal glands frequently harbor aldosterone-producing cell clusters and micronodules that are visualized by CYP11B2 immunohistochemistry (5, 6), have redefined PA as a progressive and multi-factorial syndrome that evolves as a consequence of multiple molecular abnormalities (7).

In support of this evolution, aldosterone-producing micronodules harboring pathogenic somatic mutations can be detected in morphologically normal adrenal glands from normotensive people (5, 6, 8) and in a process that parallels aging (8–10), implying that the origins of PA precede the development of hypertension, and that the pathophysiology of PA may be an important contributor to age-related hypertension. Using deep phenotyping maneuvers, human physiology studies have revealed that features known to be operative in overt PA, such as renin- and angiotensin II-independent and ACTH-mediated aldosterone production, enhanced aldosterone synthase expression, increased mineralocorticoid receptor activity, and natriuretic hormone deficiency, can also be identified in normotensive individuals (1, 11–13). In parallel, prospective studies have consistently identified a continuum of PA pathophysiology (non-suppressible and renin-independent aldosterone production) that ranges from subclinical in normotensive individuals (SubPA) to overt in individuals with severe hypertension and heightened cardiovascular disease risk (overt PA) (1, 11–21).

Hypertension is the leading cause of morbidity and mortality, affecting 1.3 billion people worldwide (22). A better understanding of the pathogenesis of PA is essential since it is a prevalent mechanism of hypertension and adverse cardiovascular outcomes. To gain these insights, we combined detailed physiologic phenotyping of aldosterone production with large-scale proteomic profiling across the spectrum of PA, from SubPA in normotensive people to overt PA in hypertensive people, to identify proteomic signatures and pathways that characterize the evolution of PA. We leveraged dose-dependent aldosterone production trends to identify proteins implicated in PA pathophysiology and functional and genetic validation studies to replicate and confirm our findings.

## RESULTS

### Study Population

The overall study design (**Figure 1**) included participants with normal blood pressure who exhibited a spectrum of renin-independent aldosterone production (SubPA) following an oral sodium loading test (OSLT) (n=61) and those with clinical diagnosed overt PA (n=50) who had more florid renin-independent aldosterone production (**Table 1**). As expected, participants with overt PA were older, had higher body mass indexes (BMI) and blood pressure, used antihypertensive medications, had lower serum potassium and kidney function, and were predominantly male, when compared to normotensive participants. The median seated plasma aldosterone concentration (PAC) trended from 7.1 to 23.0 ng/dL from SubPA to overt PA.

**Figure 1:**
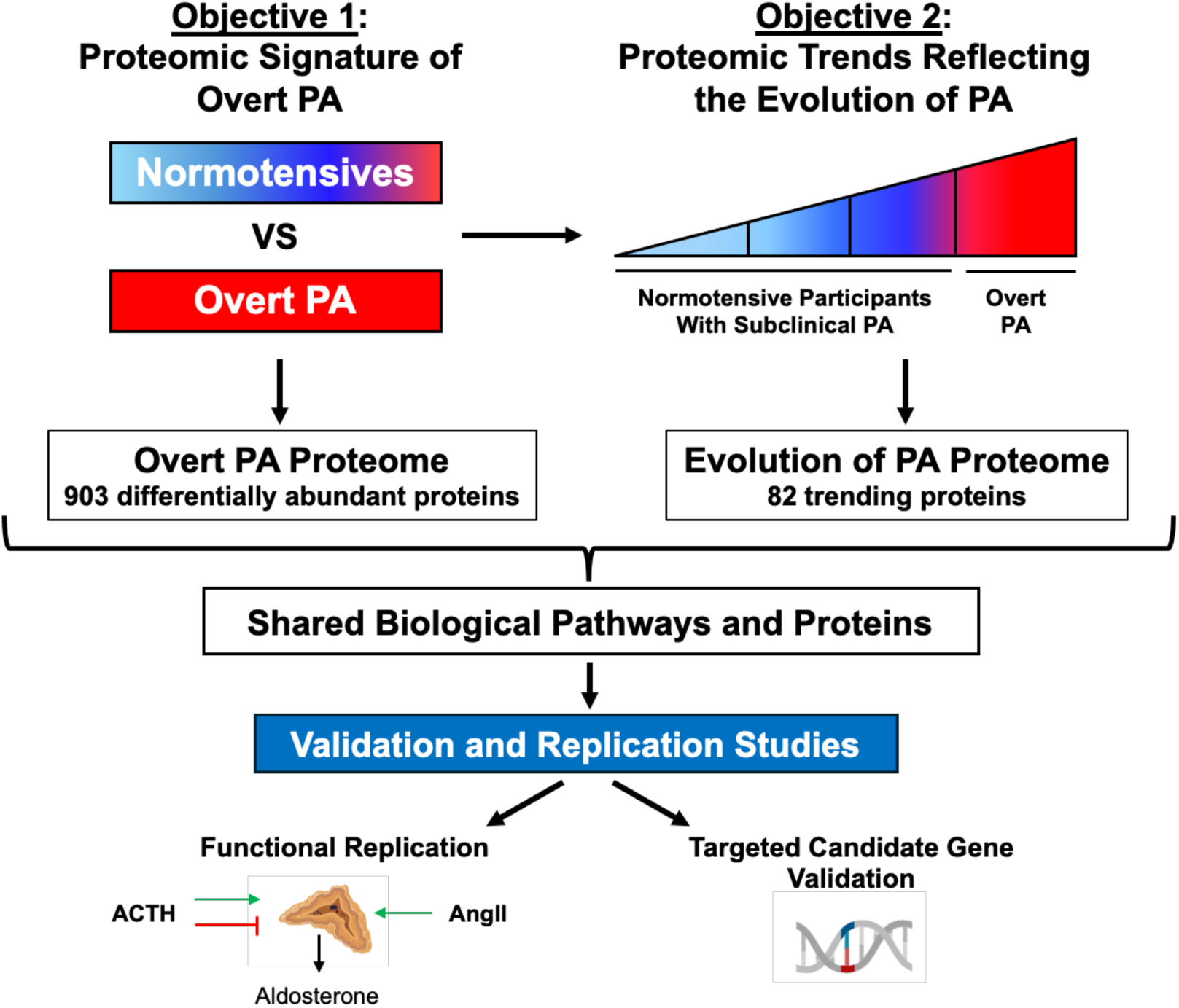
Overarching Study Design. Proteomic profiling was performed in normotensive participants who completed an oral sodium loading test to quantify renin-independent aldosterone production and in patients with overt PA (study objective 1). Subsequently, aldosterone dose-dependent trend analyses were conducted to identify proteins that paralleled the magnitude of PA pathophysiology from subclinical PA (SubPA), defined as renin-independent aldosterone production despite supine posture during an oral sodium loading test in normotensive people, to overt PA (study objective 2). To validate the results of these analyses, candidate proteins underwent separate physiological (ACTH modulated and angiotensin II-dependent) proteomic profiles for replication and a candidate gene association study was conducted in a separate cohort to confirm genotype-phenotype associations. Created using biorender.com.

**Table 1:**
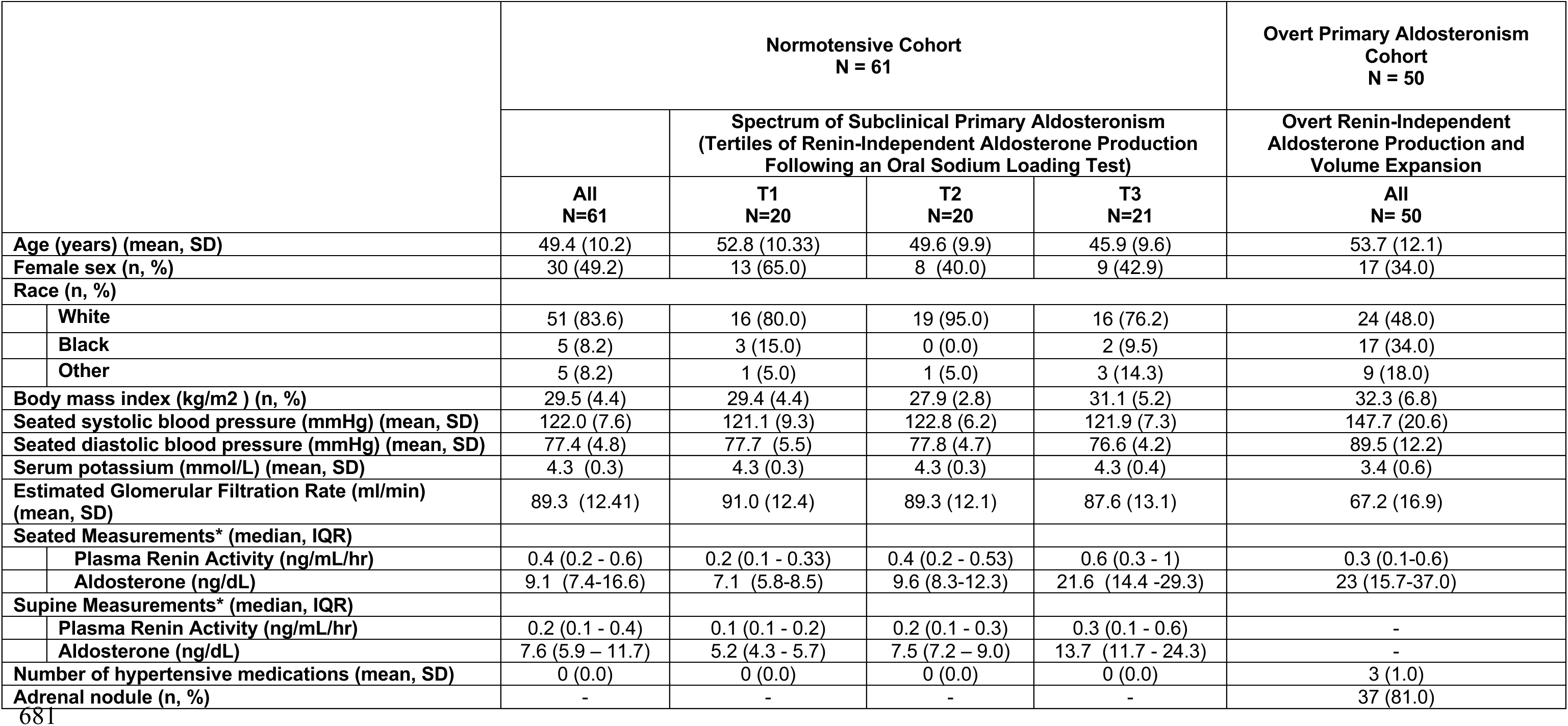
Characteristics of Study Participants. Community-dwelling normotensive participants are shown stratified by the degree of subclinical primary aldosteronism, defined as tertiles of renin-independent aldosterone production following volume expansion during an oral sodium loading test. Participants with Overt PA were clinical patients diagnosed with PA and had overt renin-independent aldosterone production and were in a state of volume expansion with suppressed renin at baseline.

|  | Normotensive Cohort<br>N = 61 |  |  |  | Overt Primary Aldosteronism<br>Cohort<br>N = 50 |
| --- | --- | --- | --- | --- | --- |
|  | Spectrum of Subclinical Primary Aldosteronism<br>(Tertiles of Renin-Independent Aldosterone Production<br>Following an Oral Sodium Loading Test) |  |  |  | Overt Renin-Independent<br>Aldosterone Production and<br>Volume Expansion |
|  | All<br>N=61 | T1<br>N=20 | T2<br>N=20 | T3<br>N=21 | All<br>N= 50 |
| Age (years) (mean, SD) | 49.4 (10.2) | 52.8 (10.33) | 49.6 (9.9) | 45.9 (9.6) | 53.7 (12.1) |
| Female sex (n, %) | 30 (49.2) | 13 (65.0) | 8 (40.0) | 9 (42.9) | 17 (34.0) |
| Race (n, %) |  |  |  |  |  |
| White | 51 (83.6) | 16 (80.0) | 19 (95.0) | 16 (76.2) | 24 (48.0) |
| Black | 5 (8.2) | 3 (15.0) | 0 (0.0) | 2 (9.5) | 17 (34.0) |
| Other | 5 (8.2) | 1 (5.0) | 1 (5.0) | 3 (14.3) | 9 (18.0) |
| Body mass index (kg/m<sup>2</sup>) (n, %) | 29.5 (4.4) | 29.4 (4.4) | 27.9 (2.8) | 31.1 (5.2) | 32.3 (6.8) |
| Seated systolic blood pressure (mmHg) (mean, SD) | 122.0 (7.6) | 121.1 (9.3) | 122.8 (6.2) | 121.9 (7.3) | 147.7 (20.6) |
| Seated diastolic blood pressure (mmHg) (mean, SD) | 77.4 (4.8) | 77.7 (5.5) | 77.8 (4.7) | 76.6 (4.2) | 89.5 (12.2) |
| Serum potassium (mmol/L) (mean, SD) | 4.3 (0.3) | 4.3 (0.3) | 4.3 (0.3) | 4.3 (0.4) | 3.4 (0.6) |
| Estimated Glomerular Filtration Rate (ml/min) (mean, SD) | 89.3 (12.41) | 91.0 (12.4) | 89.3 (12.1) | 87.6 (13.1) | 67.2 (16.9) |
| Seated Measurements* (median, IQR) |  |  |  |  |  |
| Plasma Renin Activity (ng/mL/hr) | 0.4 (0.2 - 0.6) | 0.2 (0.1 - 0.33) | 0.4 (0.2 - 0.53) | 0.6 (0.3 - 1) | 0.3 (0.1-0.6) |
| Aldosterone (ng/dL) | 9.1 (7.4-16.6) | 7.1 (5.8-8.5) | 9.6 (8.3-12.3) | 21.6 (14.4 -29.3) | 23 (15.7-37.0) |
| Supine Measurements* (median, IQR) |  |  |  |  |  |
| Plasma Renin Activity (ng/mL/hr) | 0.2 (0.1 - 0.4) | 0.1 (0.1 - 0.2) | 0.2 (0.1 - 0.3) | 0.3 (0.1 - 0.6) | - |
| Aldosterone (ng/dL) | 7.6 (5.9 - 11.7) | 5.2 (4.3 - 5.7) | 7.5 (7.2 - 9.0) | 13.7 (11.7 - 24.3) | - |
| Number of hypertensive medications (mean, SD) | 0 (0.0) | 0 (0.0) | 0 (0.0) | 0 (0.0) | 3 (1.0) |
| Adrenal nodule (n, %) | - | - | - | - | 37 (81.0) |

### The Proteomic Signature of Overt PA

Of the 1500 measured proteins (**Supplemental Table 1**), a total of 903 exhibited statistically significant differences in categorical analyses between overt PA and normotensive participants with SubPA (**Figure 2A**, **Supplemental Table 2**). The most statistically significant proteins that were more abundant in overt PA were primarily involved in glycolysis and glycation (PKLR, GLO1), coagulation and inflammation (F11, CD14), iron and hemoglobin metabolism (BPGM, CYBRD1), and calcium signaling (S100A6). In contrast, proteins that were significantly less abundant in overt PA were associated with mitochondrial function and energy production (ATP5PF, NAMPT, ECHS1, DECR1, IVD) as well as immune signaling and cell-matrix interactions (PTPN6, CCL20, ITGB1/ITGA2).

**Figure 2:**
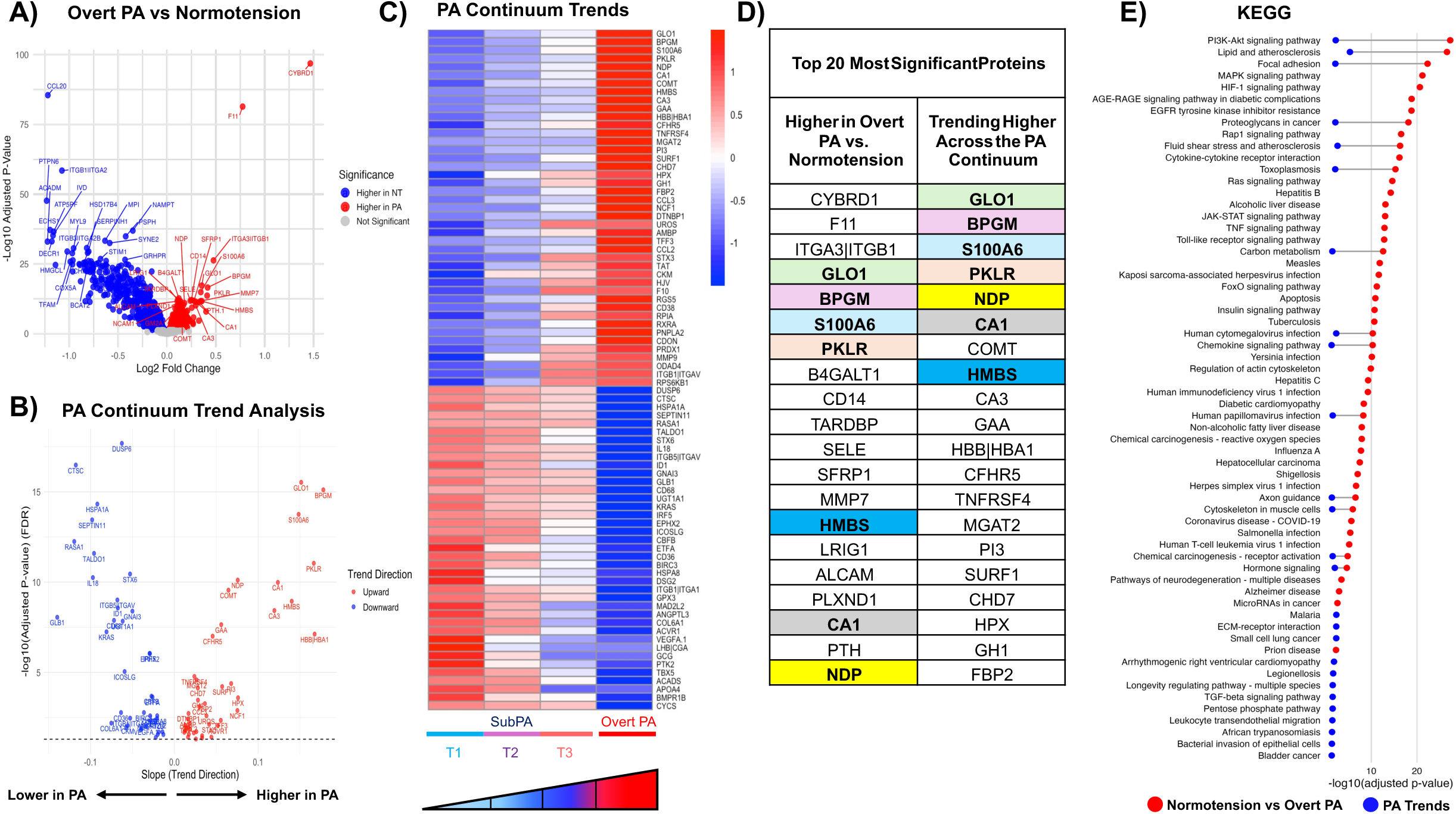
The Proteomic Signature of Primary Aldosteronism. A) Volcano plot showing 903 proteins differentially abundant between normotensive individuals (blue) and those with Overt PA (red). B) Volcano plot of the 82 proteins that exhibited consistent trends across the continuum of PA, from tertiles of SubPA to Overt PA. Proteins were required to exhibit consistently upward (red) or downward (blue) patterns and are ordered by slope magnitude. C) Heatmap of 82 proteins that exhibited consistent upward or downward trends across the PA continuum, ordered by trend direction and adjusted p-value. D) Table of the top 20 proteins most significantly abundant in overt PA (left column) and the top trending proteins across the SubPA to overt PA continuum (right column), ranked by adjusted p-value. Proteins that emerged as common between both analytic approaches are color-coded. E) KEGG pathway enrichment analysis of the overlap between the top 50 pathways implicated in overt PA and the 26 pathways associated with proteins exhibiting a stepwise trend across the PA continuum. 14 of these pathways overlapped with pathways identified in the overt PA signature, generally related to cardiovascular disease, hormone and cell signaling, and inflammation.

After adjustment for sex, BMI, systolic blood pressure, serum potassium, estimated glomerular filtration rate and age, the PA-associated proteomic signature remained broadly consistent with the unadjusted analysis (**Supplemental Figure 1, Supplemental Table 3**). As expected, covariate adjustment reduced the total number of proteins (from 903 to 526) reaching false discovery rate significance. However, the leading PA-associated proteins remained among the most significant findings, suggesting that the dominant proteomic signal was not solely explained by these clinical covariates.

Pathway enrichment analysis using KEGG identified 194 significant pathways (top 50 are shown in **Supplemental Figure 2**). Top KEGG pathways were implicated in cell signalling (PI3K-Akt signalling pathway (hsa04151), MAPK signalling pathway (hsa04010)), inflammation (cytokine-cytokine receptor interaction (hsa04060)), cell adhesion (focal adhesion (hsa04510)), and cardiovascular disease (lipid and atherosclerosis (hsa05417)).

### Proteomic Trends Across the Continuum of PA

Trend analyses requiring plasma proteins to exhibit progressive upward or downward trajectories across the continuum of normotensive SubPA and into the overt PA circulation identified 82 proteins (**Figure 2B & C**). Notably, the top proteins trending upward across this PA continuum, were also among the top 20 most significant proteins found to be more abundant in overt PA (GLO1, BPGM, S100A6, PKLR, NDP, CA1 and HMBS) (**Figure 2D**). Of the 26 KEGG pathways identified from these 82 trending proteins, 14 overlapped with pathways identified in the overt PA signature, generally related to cardiovascular disease, hormone and cell signaling, and inflammation (**Figure 2E**).

Covariate (sex, BMI, systolic blood pressure, serum potassium, estimated glomerular filtration rate and age) adjustment attenuated the number of significant proteins in the trend analysis (from 82 to 45) but preserved the dominant proteomic signal, with leading proteins remaining significant (**Supplemental Figure 3A-B**), supporting the robustness of the main phenotype-associated proteomic findings.

### Functional Replication

*In vivo* functional replication studies were performed by modulating ACTH-mediated (overnight dexamethasone suppression testing [DST] and ACTH-stimulation [ACTHstim] testing) and angiotensin II-mediated (upright posture following dietary sodium restriction) aldosterone production. Post-DST, post-ACTHstim, and post sodium-restricted upright proteomic profiles were used as separate physiological replication tools to determine whether the 82 proteins that exhibited consistent aldosterone dose-dependent trends emerged following functional testing. As expected, there was a continuum of aldosterone production following DST, ACTHstim, and sodium-restricted upright posture (11–13). In parallel, three of the 82 trending proteins: NDP, ID1, and ITGB1|ITGAV (**Supplementary Table 4**), demonstrated consistent aldosterone dose-dependent trends across all three functional replication conditions, with statistically significant trends in at least two of the three conditions. Among these, NDP showed the strongest overall replication profile, with significant trends following DST and ACTH stimulation and a near-significant trend following upright sodium-restricted testing (**Figure 3**). A complete list of trending proteins and functional replication results is provided in **Supplementary Table 4**.

**Figure 3:**
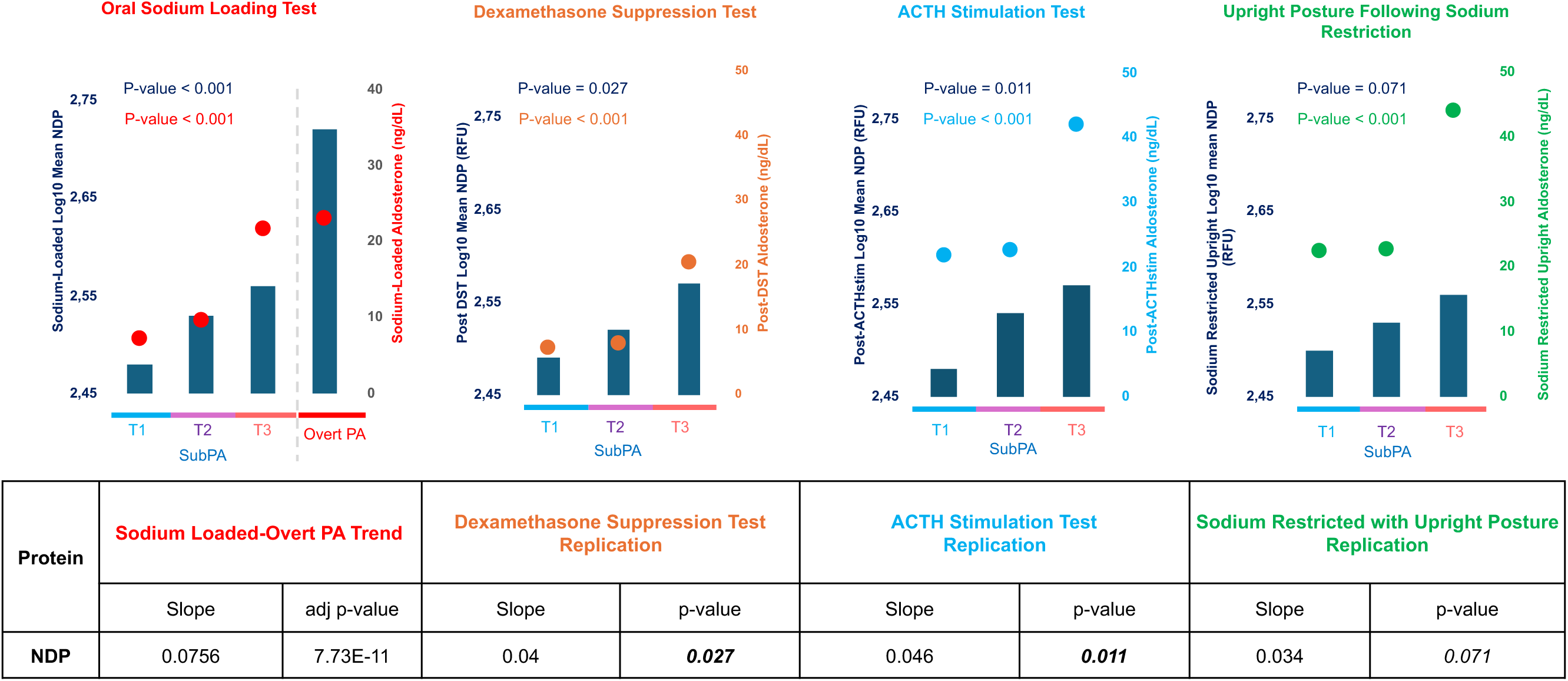
Physiologic Replication Studies. From left to right: Association between log10 mean of NDP with: the continuum of renin -independent aldosterone production, ranging from SubPA tertiles following oral salt-loading to overt PA; the continuum of ACTH-independent aldosterone production following DST; the continuum of ACTH-dependent aldosterone production following ACTHstim; and the continuum of angiotensin II-dependent aldosterone production following extreme dietary sodium restriction and upright posture.

### Candidate Gene Validation

Candidate gene validation of *NDP* as a pathogenic contributor to PA was performed using SNP association analyses with multiple well-phenotyped aldosterone traits in an independent cohort of participants from HyperPATH (**Table 2**). In normotensive participants, 2 SNPs were associated with higher PAC following infusion of exogenous AngII, 6 SNPs showed positive and significant dose-dependent associations with 24-hour urinary aldosterone excretion following OSLT, and 7 SNPs were significantly associated with upright PAC following extreme sodium restriction. In the combined cohort of normotensive and hypertensive participants, 6 SNPs were significantly associated with supine PAC following OSLT, 2 SNPs with PAC following infusion of exogenous AngII, 22 SNPs with 24-hour urinary aldosterone excretion following OSLT, and 12 SNPs with upright PAC following extreme sodium restriction. Notably, multiple SNPs were associated with more than one aldosterone trait and were supported by eQTL signals in adrenal tissue (**Table 2**).

**Table 2:**
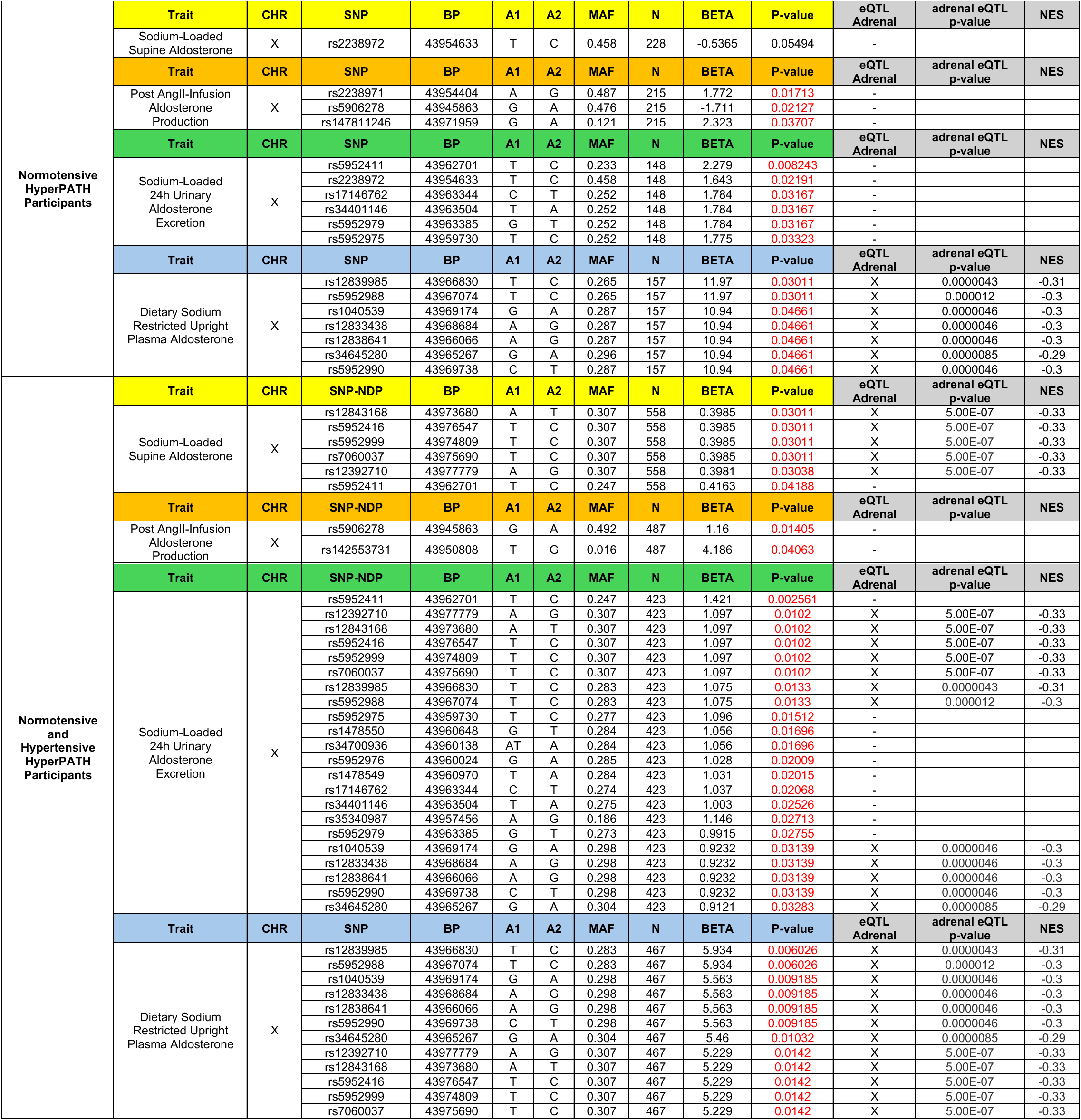
*NDP* gene SNP association analyses with multiple aldosterone traits from normotensive and hypertensive participants in the HyperPATH cohort.

The top associated SNP, rs5952411 (β = 1.42, P = 0.0025 for 24-hour urinary aldosterone), was functionally annotated using *VannoPortal* and found to lie in an enhancer region interacting with *NDP-AS1*, a long non-coding RNA adjacent to the *NDP* gene, suggesting it may influence *NDP* expression through regulatory chromatin interactions.

## DISCUSSION

PA causes cardiovascular disease via excessive activation of the mineralocorticoid receptor (MR) (19, 23, 24). The last decade has witnessed a maturing recognition of the widespread prevalence of PA and identification of its earliest manifestations (1, 2). Although PA is usually synonymous with hypertension, human physiology studies and large population-based cohort studies have consistently shown that a continuum of PA can even be detected in normotensive people (termed subclinical PA) where it increases risk for developing hypertension and cardiovascular disease (1, 12, 13, 15–18, 21, 25–28). Functional drivers of this continuum include pathogenic somatic mutations in the adrenal cortex, dysregulated aldosterone synthase expression, altered angiotensin II- and ACTH-mediated aldosterone production, and increased mineralocorticoid receptor activity (1, 11–13). Building on these prior studies, we now show that this continuum is mirrored at the proteomic level. Using large-scale proteomics and detailed phenotyping of aldosterone production, we identified proteins with robust dose-dependent abundance patterns that spanned from normotensive people with renin-independent aldosterone production to those with clinically overt PA, clustering in pathways known to be operative in cardiovascular disease. Among these implicated proteins, the evidence to support Norrin (NDP) in the pathogenesis of PA was strong and validated with multimodal phenotypic and genotypic assessments.

The revelation of a proteomic continuum of PA pathophysiology that originates in people with normal blood pressure has major impacts for understanding the pathogenesis of hypertension and cardiovascular disease and targeted treatments for these conditions. The top proteins trending upward through the PA continuum (GLO1, BPGM, S100A6, PKLR, NDP, CA1, HMBS) were also among the top 20 most significant proteins found to be higher in categorical analyses of overt PA, suggesting their key contribution to defining the PA proteomic signature. Several of the identified proteins (GLO1, PKLR) are involved in glycolysis, a pathway shown to be upregulated in aldosterone-producing adenomas (29, 30) and in a subsets of aldosterone-producing cell clusters (30) to support increased energy production and cell growth. The stepwise increase in glycolytic proteins across the continuum of PA suggests early metabolic reprogramming might be a feature of the pathogenesis of PA. We also show that hemoglobin and oxygen-related proteins (HBB|HBA1, HPX, HMBS, CA1, CA3, BPGM) trended across the PA continuum. Incidentally, increased hemoglobin was also recently identified in another proteomic characterization of PA (31). Several other proteins involved in aldosterone biosynthesis or adrenal cortex physiology also displayed stepwise trends across the PA continuum, but did not retain consistent patterns in physiological replication tests. This does not diminish their potential relevance; rather, it suggests that their abundance might be differentially influenced by ACTH and/or angiotensin II.

The *NDP* gene encodes for Norrin, a protein originally identified through its association with Norrie disease, a rare congenital disorder characterized by abnormal development of retinal blood vessels (32). In the eye, Norrin activates canonical Wnt signalling by binding to Frizzled4 (FZD4) and LRP5/6, with TSPAN12 acting as a co-receptor that enhances ligand capture and facilitates complex formation. This interaction leads to stabilization of β-catenin, to nuclear translocation, and to transcriptional activation of Wnt target genes (33). Support of the implications of our current work stems from recent genome-wide association studies where *NDP* was identified as a potential contributor to PA and hypertension. In 2022, Le Floch et al. identified *NDP* as a risk locus for PA in a large French discovery cohort and the lead SNP (rs5905587) was supported by adrenal-specific eQTL data (GTEx) linking genetic variation to local *NDP* RNA expression (34). In 2023, Naito et al. identified the *NDP* locus as significantly associated with risk for hypertension (35). Subsequently, Inoue et al. conducted a cross-ancestry meta-analysis combining data from these Japanese and French cohorts, as well as from the UK Biobank and Finnish cohorts, and affirmed the association of *NDP* with risk for PA (36). In a large X-chromosome-wide, multi-ancestry, association study of heart failure, *NDP* loci were associated with heart failure, a well-established consequence of PA that is also mitigated by aldosterone-directed therapy (37). Moreover, NDP has been identified as a factor secreted by heart tissue after injury (38); in a mouse model of myocardial infarction, *NDP* gene expression was increased in cardiac stromal cells following infarction and the resulting NDP protein was shown to directly stimulate the proliferation of fibroblasts(38). Collectively, our current findings add substantially to this prior evidence implicating NDP in the pathogenesis of PA and its comorbid consequences by demonstrating its role throughout the entire continuum of PA, detectable in normotensive people with SubPA, and strongly linked to germline variations in the *NDP* gene.

Although little is known about the interplay between Norrin signaling and adrenal function to fully explain the mechanisms underlying our genotype-phenotype-physiology findings, recent evidence has identified TSPAN12, a known co-receptor of Norrin, as a negative regulator of aldosterone production (39). TSPAN12 expression is reduced in aldosterone-producing adenomas, particularly those with *KCNJ5* pathogenic variants, and its loss is associated with increased *CYP11B2* expression and elevated basal and post-angiotensin II stimulated aldosterone production (39). Norrin has also been identified as a ligand for the R-spondin 4 receptor (LGR4), a widely expressed G protein-coupled receptor in the adrenals, capable of activating multiple downstream pathways depending on its ligand, including cAMP/PKA, Wnt/β-catenin, Gαq/GSK3β and Gαq/PKCα signalling cascades (40). LGR4 appears to play a critical role in adrenal cortex zonation and the structural integrity of the zona glomerulosa and possibly aldosterone production, as evidenced by a familial case of isolated hypoaldosteronism linked to a homozygous pathogenic variant in *LGR4* (41). Moreover, in an *in vivo* mouse model, conditional inactivation of *Lgr4* in steroidogenic adrenal cells led to disrupted zona glomerulosa differentiation, near-complete loss *of CYP11B2* expression, and markedly reduced aldosterone production (41), possibly due to decreased canonical Wnt signalling (41). Thus, many components of Norrin-associated signaling pathways have been linked to zona glomerulosa function and PA biology, which serve as a foundation for our current findings to extend hypothetical frameworks (**Figure 4**).

**Figure 4:**
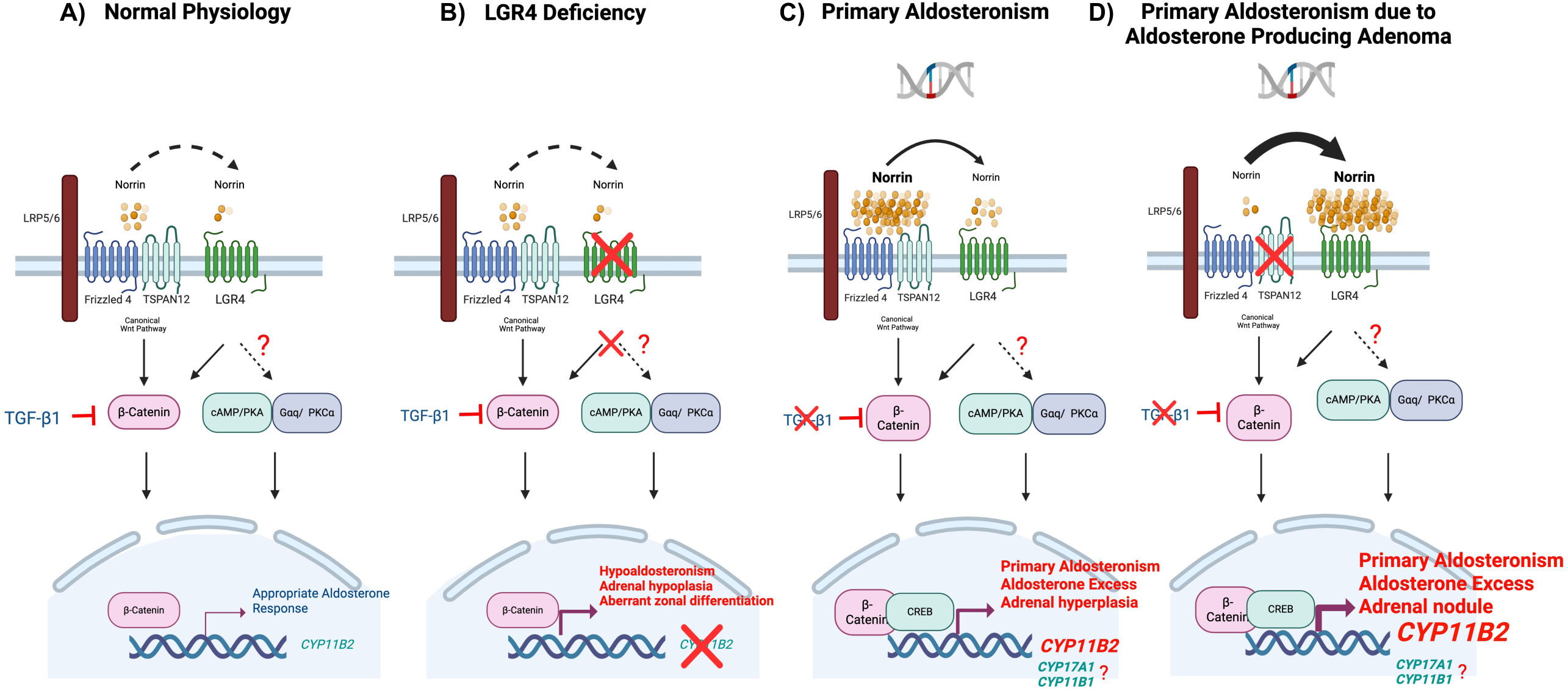
Hypothetical Model of the Role of Norrin in Primary Aldosteronism Pathophysiology. Created in biorender. A) In normal adrenal cortical and zona glomerulosa physiology, Norrin is expected to primarily signal through Frizzled-4 (FZD4) and LRP5/6, with TSPAN12 serving as a co-receptor that enhances canonical Wnt/β-catenin activation. Although Norrin can also bind LGR4, TSPAN12 preferentially facilitates its interaction with FZD4. In parallel, TGF-β1 suppresses aldosterone production by reducing β-catenin levels. B) Prior studies have shown that loss of LGR4 function in knockout models results in hypoaldosteronism, adrenal hypoplasia, and disrupted zona glomerulosa architecture due to insufficient Wnt/β-catenin signalling. C) In the pathophysiology of PA, genetic variation in the NDP locus might lead to increased local Norrin abundance, which could enhance paracrine activation of Wnt/β-catenin signaling primarily via FZD4 and potentially through LGR4, leading to increased CYP11B2 activity. Additionally, Norrin-induced inhibition of TGF-β1 signaling would potentially diminish its inhibitory effect on aldosterone production. Norrin–LGR4 interactions might also engage PKC and PKA pathways. D) In aldosterone producing adenomas, where prior studies have reported reduced expression of TSPAN12, higher levels of Norrin might shift signaling away from FZD4 toward LGR4, promoting Wnt/β-catenin activation and potentially engaging PKC and PKA pathways. This shift might enhance transcription of CYP11B2 and, more speculatively, CYP17A1 and CYP11B1. This combined signaling cascade might drive adrenal cell proliferation, aldosterone overproduction, and hybrid steroid synthesis. In parallel, Norrin-mediated inhibition of TGF-β1 signaling might further contribute to aldosterone excess.

While the strengths of this multimodal phenotyping study included comprehensive physiologic testing to characterize the continuum of PA and multiple filters to limit the possibility of false discovery, the results must be interpreted in the context of several limitations. First, the proteomic measurements included 1,500 proteins; therefore, thousands of proteins in the human proteome were unmeasured. Second, we compared a normotensive population with an overt PA group but did not study populations with hypertension or resistant hypertension. However, our approach permitted comparison of extreme phenotypes to more easily characterize the PA proteome; we then employed strict trend analyses to confirm that candidate proteins exhibited aldosterone dose-dependent patterns. We also included hypertensive participants in our validation candidate gene association studies to confirm that *NDP* variants were associated with aldosterone production traits in both normotensive and hypertensive people. Third, despite SNPs being associated with dose-dependent aldosterone production, there were negative normalized effect sizes for *NDP* expression in adrenal eQTL data, whereas Norrin abundance was seen to be higher across the PA continuum. This discrepancy could be explained by post-transcriptional or translational regulation, increased protein stability, or by the influence of tissue heterogeneity in eQTL datasets, which are derived from bulk adrenal samples containing cortex, medulla, and periadrenal fat, potentially masking zone-specific signals (42). Finally, although Norrin emerged as a strong candidate through proteomics, physiological replication, and genetic validation, its direct role in driving CYP11B2 expression was not directly evaluated. Further, our study (and priors) were not designed to determine the tissue source(s) of Norrin contributing to our findings. However, our results provide a mechanistic basis for prior GWAS and XWAS studies that identified *NDP* variants as potentially causal factors for PA and demonstrate that this relationship evolves over a continuum of PA pathophysiology beginning in normotensive people.

In conclusion, the evolution of PA is characterized by proteomic signatures that originate in normotensive individuals and involve biologic pathways known to be operative in the pathogenesis of cardiovascular disease. These results imply that a large proportion of essential hypertension might be driven by PA pathophysiology, and thus amenable to established and emerging therapies that target aldosterone and/or the mineralocorticoid receptor (43–46). Our data support the presence of a proteomic continuum that mirrors the spectrum of renin-independent aldosterone production, representing gradations of disease severity rather than classifying PA as a categorical or binary diagnosis. Within this continuum, Norrin demonstrated robust aldosterone dose-dependent trends across proteomic, physiologic, and genetic analyses, indicating its potential causal role in the pathophysiology of PA that might be a target for future therapeutics.

## METHODS

### Sex as a biological variable

Both female and male participants were included in this study. Sex was considered as a biological variable in the study design. Accordingly, sex was included as a covariate in adjusted proteomic analyses.

General Study Design:

Participants were recruited through two prospective studies: one enrolling a cohort of community-dwelling normotensive individuals to undergo deep-phenotyping studies to characterize subclinical PA (SubPA) (NCT03484130), and the other a prospective registry enrolling patients with overt PA: individuals with an established clinical diagnosis PA. The first objective was to characterize the peripheral plasma proteome of PA by comparing the circulating proteome of participants with established clinically overt PA with that of normotensive participants following an oral sodium loading test (OSLT). The second objective was to identify proteins that consistently and reliably trended across the continuum of PA severity (continuum of renin-independent aldosteronism) in an aldosterone dose-dependent manner. To facilitate this analysis, normotensive participants were stratified into unbiased categories of SubPA, defined as the magnitude of non-suppressible and renin-independent aldosterone production, despite supine posture and following the OSLT (**Figure 1**). Candidate proteins identified in the trend analysis were then subjected to multimodal *in vivo* replication and validation, including physiological testing (using ACTH- and Angiotensin II-modulated proteomics) and genetic testing (candidate gene SNP association studies) of an independent cohort that also underwent detailed aldosterone physiologic testing (**Figure 1**).

Normotensive Participants:

Normotensive participants with risk factors for developing incident hypertension (BMI ≥ 25 kg/m², family history of hypertension prior to the age of 60 years in a parent or sibling, diabetes with a hemoglobin A1c < 9%) were prospectively recruited from the Boston, MA metropolitan area between July 2018 and October 2022 as previously described (12, 13). All participants completed an OSLT to assess the magnitude or renin-independent aldosterone production (the signature of PA) and were then stratified into unbiased tertiles of SubPA, based on the magnitude of plasma aldosterone levels while in a supine position following an OSLT, to facilitate evaluation of the presence and severity of SubPA and subsequent trend analyses. The OSLT involved consuming >200 mmol/d of sodium for 5-7 days to induce intravascular volume expansion and physiologic suppression of renin and angiotensin II, thereby facilitating interrogation of the degree of renin-independent aldosteronism. The combination of supine posture and sodium loading was used to induce a physiologic nadir in aldosterone production to characterize the spectrum of SubPA (12, 13). This maneuver permitted comparison of normotensive participants with SubPA to individuals with clinically overt PA, all of whom had severe renin-independent aldosteronism despite volume expansion, by definition.

### Overt PA Participants

50 patients with established clinically overt PA were included after undergoing informed consent to participate.

### Laboratory Assays

Normotensive participants had plasma aldosterone concentration (PAC) and plasma renin activity (PRA) measured as previously described (12, 13) and participants with overt PA had PAC measured as previously described (47).

### Plasma Proteomics

Normotensive participants had proteomic measurements performed on four separate occasions: following OSLT, following an overnight 1mg dexamethasone suppression test (DST), following an ACTH stimulation (ACTHstim) test, and following an extreme sodium restriction intervention while in upright posture (protocols described below). Patients with clinically overt PA underwent proteomic profiling of peripheral blood samples only.

Proteomic measurements were performed using the SomaScan® assay, an aptamer-based platform that quantifies proteins using chemically modified single-stranded DNA aptamers called SOMAmers (Slow Ofrate Modifed Aptamer) (48). Detailed methods of the SomaScan® assay have been previously described (49, 50). This platform is well-validated and widely used in large-scale proteomic studies (51–53). Proteomic profiles were characterized using the 7k SomaScan assay v4.1 for the normotensive participants and the SomaScan® 11K Assay v5.0 (SomaLogic, Inc.; Boulder, CO, USA) for overt PA participants. Both assays have been well validated (48, 54). 1500 proteins were measured to reflect cardiovascular, metabolic, and inflammation profiles and are shown in **Supplemental Table 1**. The SomaLogic normalization procedure, including adaptive normalization by maximum likelihood, was used for the SomaScan® data for all analyses. Data from the 11K SomaScan assay were bridged to the 7K format using the SomaDataIO R package for cross-platform comparability (54). Data were processed in R v4.3.2 using the SomaDataIO package for loading raw proteomic data. Normality of protein abundance distributions was evaluated using visual inspection (i.e., histograms, QQ plots), and log-transformation (log_10_) was applied to approximate normality and stabilize variance.

### Statistical Analysis

Categorical variables were reported as percentages, normally distributed variables were reported as mean ± standard deviation, and non-normally distributed variables were reported as median (25^th^-75^th^ percentile interquartile range).

Differential abundance of plasma proteins between normotensive participants and those with overt PA was assessed using the *limma* package in R, which applies linear modeling with empirical Bayes moderation to improve variance estimation. To establish an overt PA proteomic signature, Pathway enrichment was conducted using Kyoto Encyclopedia of Genes and Genomes (KEGG)(55). The top 50 pathways were used to represent the overt PA proteomic signature.

Proteomic trend analyses were performed using linear regression across 4 categories of the PA continuum: unbiased tertiles of SubPA (termed T1, T2, T3) and the peripheral circulation of participants with overt PA (**Figure 1**). Proteins that exhibited statistically significant aldosterone dose-dependent trends, and a consistent direction of change across all four categories (e.g. either progressively increasing or progressively decreasing across all four categories), were classified as relevant proteins of interest. Identified trending proteins were visualized using a volcano plot and heatmap. Pathway enrichment using Kyoto Encyclopedia of Genes and Genomes (55) analyses was performed to identify pathways overlapping with those identified in the overt PA proteomic signature.

All statistical analyses were conducted using R, version 4.3.2 (R Project for Statistical Computing). P-values were adjusted for multiple testing using the Benjamini-Hochberg method (56). An adjusted p-value less than 0.05 was considered statistically significant.

### Sensitivity Analyses

Sensitivity analyses were conducted to interrogate the robustness of the findings. Analyses evaluating the differential protein abundance between participants with overt PA versus those with normotensive SubPA were repeated using covariate-adjusted limma models. Protein abundances were modeled with adjustment for sex, BMI, systolic blood pressure, serum potassium, estimated glomerular filtration rate and age, with Benjamini-Hochberg correction applied for multiple testing. The aldosterone dose-dependent trend analyses described above were also repeated after adjustment for the same covariates.

### Validation and Replication Studies

#### Functional Replication: Proteomic Profiling Following ACTH- and Angiotensin II-Modulations

To validate the involvement of trending (from SubPA to overt PA) proteins in the pathogenesis of PA, proteomic profiles were measured across 3 independent physiological studies conducted in the same participants on two separate days. These maneuvers to interrogate ACTH-dependent, ACTH-independent, and AngII-dependent production have been previously used to characterize both normotensive SubPA and overt PA (11–13). On a separate study day, normotensive participants with SubPA underwent pharmacological modulation of ACTH-mediated aldosterone production. A dexamethasone suppression test (DST: 1mg of dexamethasone administered at 23h00-0h00 with blood sampling at 08h00 the following morning) was used to evaluate ACTH-independent aldosterone production, while an ACTH stimulation test (ACTHstim: 250 mcg bolus of cosyntropin with sampling 60 minutes later) was used to assess ACTH-dependent aldosterone secretion, as previously described (12, 13). Blood samples were collected after each intervention to measure serum PAC and to measure the plasma proteome. On a separate study day, participants underwent assessment of AngII-dependent aldosterone production using sodium restriction and upright posture interventions to induce renin- and AngII-dependent aldosteronism. Normotensive participants with SubPA completed 5 to 7 days of extreme dietary sodium restriction (∼10 mEq per day) prepared by professional dietary staff in the metabolic kitchen to achieve a mean urine sodium excretion of 16.1 ± 11.2 mEq/24h (50). They were then evaluated while fasting at 08h00 in the ambulatory clinical research center with venipuncture following 30 minutes of upright posture to measure PAC and the peripheral proteome. This combination of extreme sodium restriction and upright posture was used as a maneuver to maximally stimulate endogenous AngII, and consequently AngII-dependent aldosterone production (12, 13).

#### Independent Candidate Gene Validation: Dose-Dependent SNP Associations

To validate the most robustly identified protein that emerged from the aforementioned analyses, a candidate-gene association study was performed in an independent population of participants from the HyperPATH cohort. The HyperPATH cohort was designed to elucidate the genetic determinants of adrenal hypertension and has been previously described (57). Study participants included both normotensive (n=228) and hypertensive (n=330) individuals without clinically diagnosed PA, recruited from the community, studied without interfering anti-hypertensive medications, and who underwent strictly controlled dietary sodium interventions to evaluate PA pathophysiology as well as genotyping. The value of independent genotype-phenotype confirmation in the HyperPATH cohort was the availability of robust aldosterone measurements following multiple interventions to modulate aldosterone physiology: OSLT (to assess renin-independent aldosteronism), following an infusion of exogenous AngII (to assess AngII-dependent aldosteronism), and following extreme dietary sodium restriction and upright posture (to assess AngII-dependent aldosteronism via a distinct intervention).

Single nucleotide polymorphisms (SNPs) within the *NDP* gene region were selected based on chromosomal coordinates chrX:43,808,022–43,832,750 (GRCh37), including ±5 kb flanking regions to capture regulatory and promoter variation. Genotyping data were imputed using the TOPMed Imputation Server (https://imputation.biodatacatalyst.nhlbi.nih.gov) with the 1000 Genomes Project Phase 3 as the reference panel, thereby increasing genomic coverage and resolution beyond directly typed variants. To ensure accurate imputation and harmonization across genotyping platforms, we excluded SNPs with imputation quality scores (INFO) < 0.8 or minor allele frequency < 1%. Association analyses were conducted for 4 aldosterone production phenotypes, reflecting suppressed and stimulated angiotensin II conditions, and treated as continuous variables:

(1) Supine PAC following OSLT: a reflection of renin- and angiotensin II-independent aldosterone production;
(2) 24-hour urinary aldosterone excretion following an OSLT: another distinct reflection of renin- and angiotensin II-independent aldosterone production;
(3) PAC following an infusion of exogenous angiotensin II (3 ng/kg/min for 60 mins): a reflection of angiotensin II-dependent aldosterone production;
(4) Upright PAC following extreme dietary sodium restriction (10 mEq per day for 5-7 days): another reflection of angiotensin II-dependent aldosterone production.

Genotype-phenotype associations were assessed using additive linear regression models, implemented in the PLINK software (v1.9). SNP genotypes were coded additively based on the number of effect alleles (A1: 0, 1, or 2), under the assumption that the effect of each additional allele is additive on the trait scale. Effect sizes (β coefficients) were interpreted as the expected change in aldosterone level per additional copy of the A1 allele. To account for multiple testing, false discovery rate (FDR)-adjusted p-values were computed, and statistical significance was defined as FDR <0.05. To ensure robustness of findings, sensitivity analyses were conducted using quantile-normalized aldosterone traits and controlling for potential confounders such as age, sex, and BMI. These did not materially alter the associations. Given that *NDP* resides on the X chromosome, genotype coding was harmonized across sexes: male hemizygous genotypes were recoded as either 0 or 2 to match the dosage scale in females (0, 1, 2), thereby enabling uniform modeling of additive genetic effects. This approach preserves comparability between sexes while retaining statistical power. SNPs were analyzed for tissue-specific expression quantitative trait loci (eQTLs) using data from the GTEx Portal (https://gtexportal.org), accessed on April 14, 2025. To further elucidate functional consequences, we cross-referenced the top SNP signal at *NDP* (rs5952411; associated with higher urinary aldosterone excretion, β = 1.42, P = 0.0025) using the VannoPortal platform (http://www.mulinlab.org/vportal/index.html).

### Study Approval

All normotensive participants provided written informed consent prior to participating and the study protocol was approved and monitored by the Mass General Brigham Institutional Review Board. All participants with overt PA provided written informed consent prior to participating, and the study protocol was approved and monitored by the Beth Israel Deaconess Medical Center Institutional Review Board. Human participants in this study were involved in a prospective physiology protocol (NCT03484130) and from a longitudinal patient registry. All human study protocols were approved and supervised by the human ethics and research committees and Mass General Brigham and Beth Israel Deaconess Medical Center.

## Author contributions

Conceptualization: AV, SPL

Methodology: AV, SPL, MH

Investigation: AV, SPL, AFT, JSW, MM, BS

Visualization: AV, MH, SPL

Project administration: SM, IH, LCT, BH, AV

Funding acquisition: AV

Supervision: AV

Writing - original draft: SPL, AV, MH

Writing - review & editing: SPL, MH, CHT, JMB, AJN, SM, IH, BH, LCT, SW, KI, MCZ, RA, AFT, JSW, BS, MM, AV

## Data Availability

Data supporting the results are available from the corresponding author.

## Acknowledgments and Funding

We thank the National Institutes of Health for support through awards R01DK115392 (AV, RA), R01HL153004 (AV), and R01HL155834 (AV, AFT).

## List of Supplementary Materials

Fig. S1: Covariate-adjusted peripheral proteomic signature of overt PA

Fig. S2: Top 50 KEGG Pathways in the Overt PA Signature

Fig. S3: Volcano plot and heat map from trend analysis adjusted for covariates

Table S1: Proteomic dataset comprising 1500 measured proteins

Table S2: Volcano Plot Analysis: Differential Proteins in PA vs. Normotensives

Table S3: Trending Proteins with Supporting Functional Replication Results Table S4: Replication Studies

