## Supplementary figures and images for "The Evolution of Primary Aldosteronism and the Role of Norrin"

### Supplemental Figures

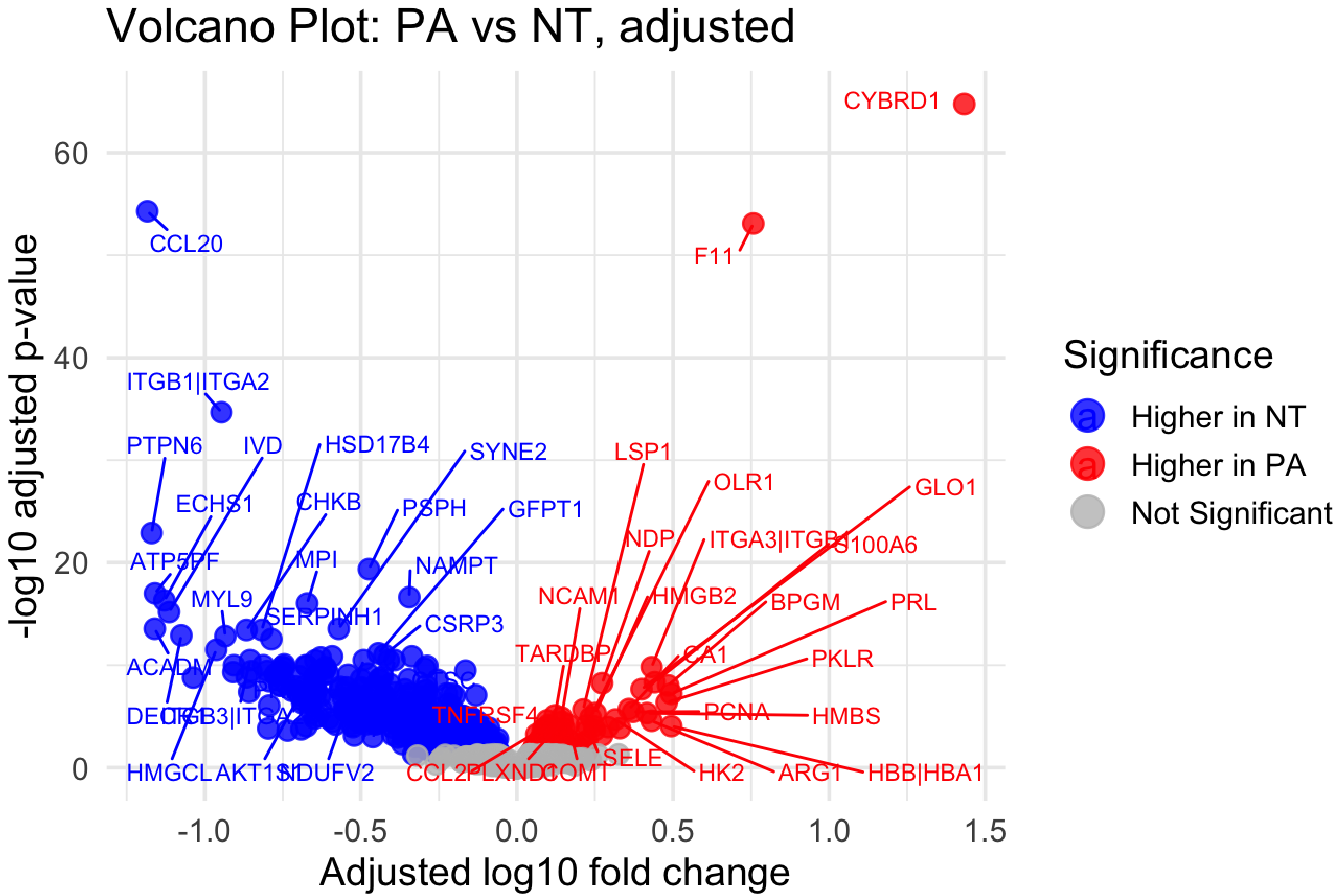

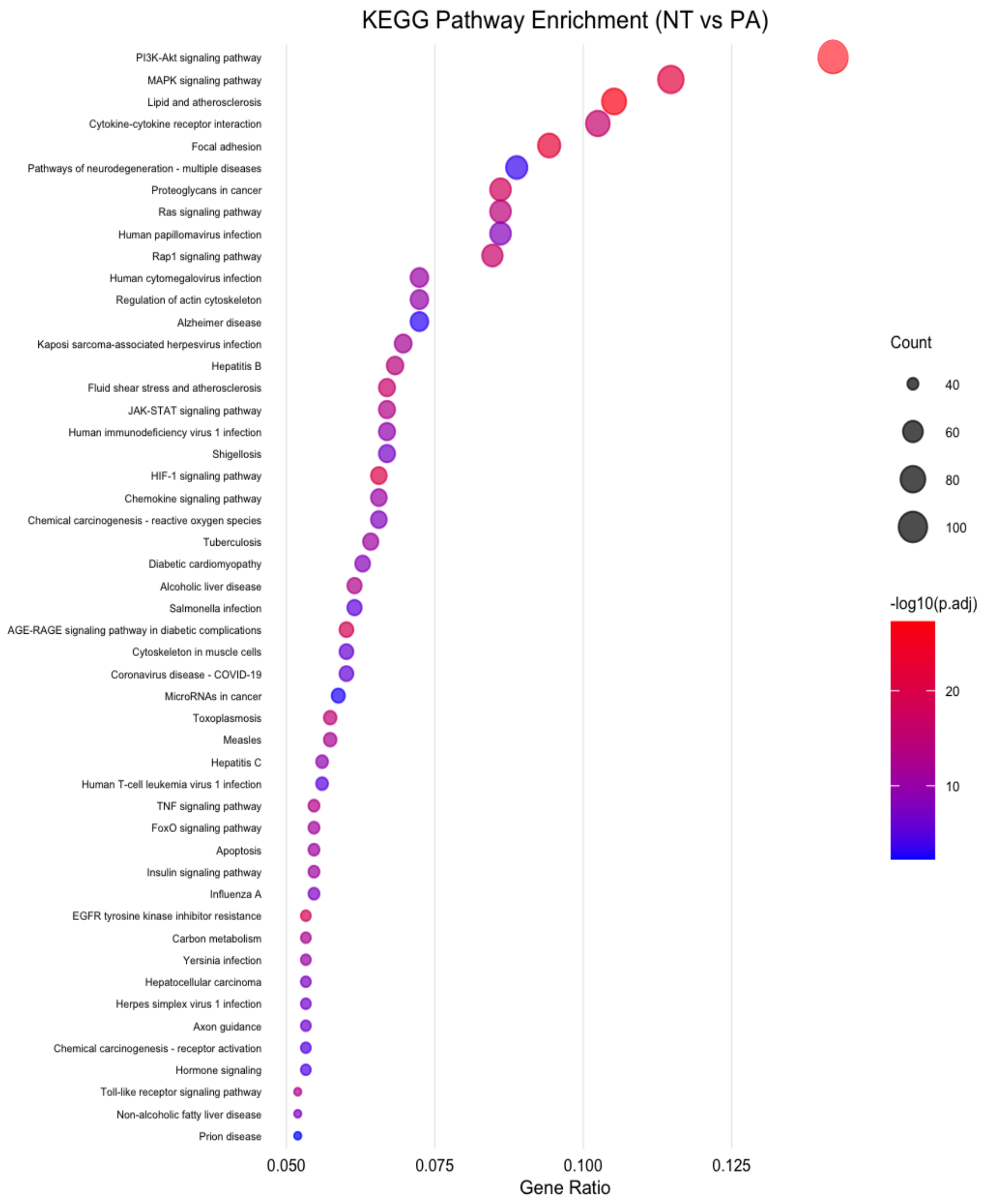

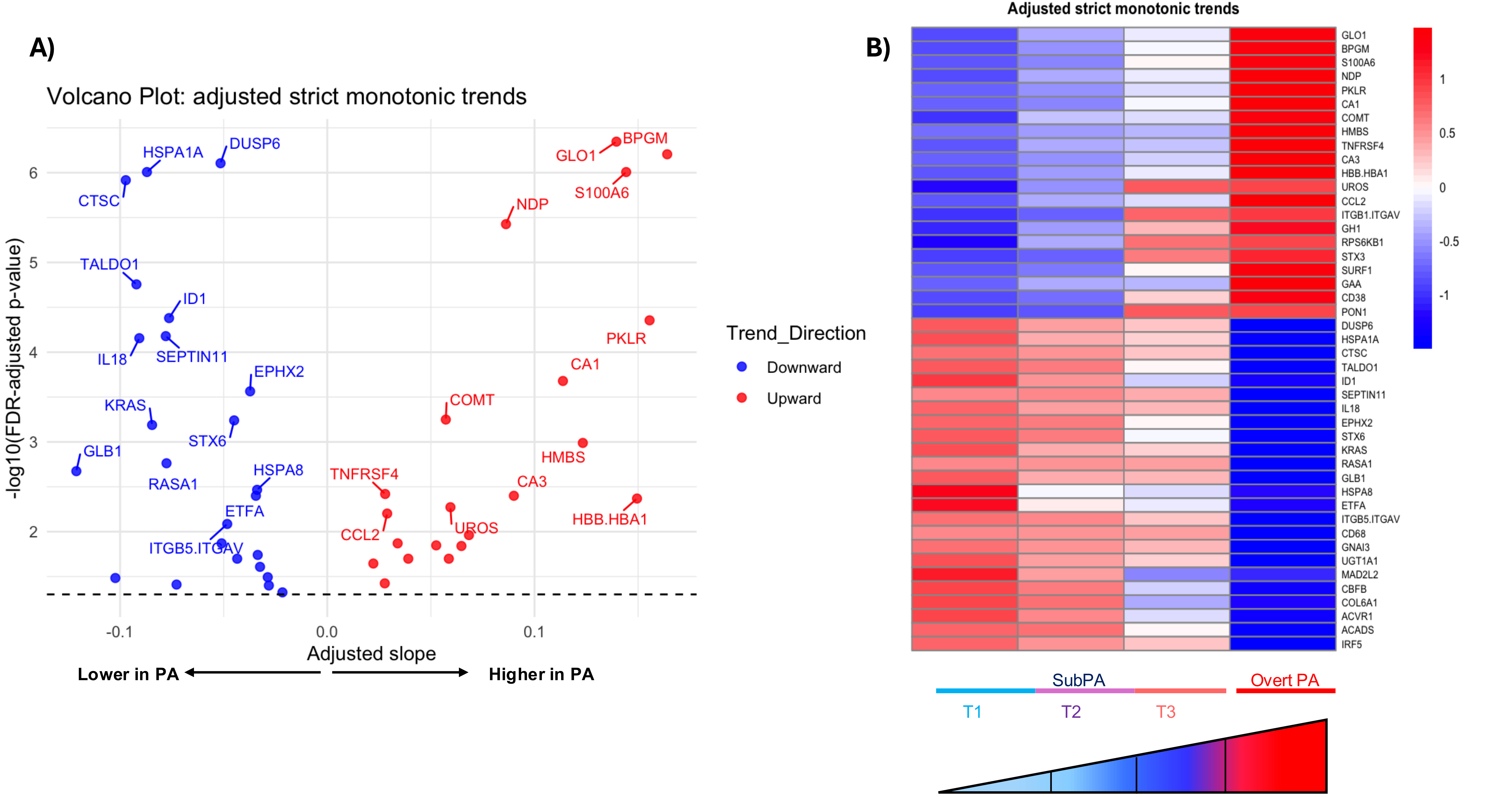
