## Supplemental Table 1 for "The Evolution of Primary Aldosteronism and the Role of Norrin"

| SeqId | Target | GeneSymbol |
|---|---|---|
| 10001-7 | c-Raf | RAF1 |
| 10011-65 | OCRL | OCRL |
| 10014-31 | SLUG | SNAI2 |
| 10022-207 | POLH | POLH |
| 10023-32 | VDR | VDR |
| 10024-44 | HOGA1 | HOGA1 |
| 10041-3 | HNF4A | HNF4A |
| 10043-31 | BRD4 | BRD4 |
| 10048-7 | PEBB | CBFB |
| 10053-5 | ILK1 | ILK |
| 10063-10 | FANCL | FANCL |
| 10078-5 | BAG3 | BAG3 |
| 10085-25 | STAR | STAR |
| 10086-39 | CBS | CBS |
| 10088-37 | APT | APRT |
| 10346-5 | STAT3 | STAT3 |
| 10351-51 | IRF1 | IRF1 |
| 10356-21 | c-Jun | JUN |
| 10362-35 | c-Myc | MYC |
| 10363-13 | SMAD3 | SMAD3 |
| 10364-6 | SMAD2 | SMAD2 |
| 10365-132 | IL-23 | IL12B\|IL23A |
| 10366-11 | PDGFRA | PDGFRA |
| 10367-62 | IL-12 | IL12A\|IL12B |
| 10391-1 | ANGL3 | ANGPTL3 |
| 10396-6 | Mcl-1 | MCL1 |
| 10426-21 | GOSR2 | GOSR2 |
| 10435-2 | ASPG | AGA |
| 10480-33 | CD59 | CD59 |
| 10521-10 | MXRA8:ECD | MXRA8 |
| 10531-18 | RASN | NRAS |
| 10534-40 | PARP:BRCT domain | PARP1 |
| 10550-37 | BMP RIB | BMPR1B |
| 10551-7 | LAT | LAT |
| 10552-88 | NKG2F | KLRC4 |
| 10554-23 | BGAL | GLB1 |
| 10574-10 | b2-Microglobulin | B2M |
| 10584-7 | NDUS4 | NDUFS4 |
| 10618-190 | megalin | LRP2 |
| 10623-19 | MUC1:region 1 | MUC1 |
| 10627-87 | APLP2 | APLP2 |
| 10638-1 | TINF2 | TINF2 |
| 10666-7 | GNPTG | GNPTG |
| 10667-78 | CA185 | C1orf185 |
| 10710-23 | ZPBP2 | ZPBP2 |
| 10714-7 | ACE | ACE |

| | | |
|---|---|---|
| 10761-5 | TMED2 | TMED2 |
| 10800-15 | Collagen-binding prote | SERPINH1 |
| 10818-36 | ASM | SMPD1 |
| 10889-2 | MRAP2 | MRAP2 |
| 10892-8 | OSMR | OSMR |
| 10945-11 | Syntaxin-6 | STX6 |
| 10955-4 | CLC10 | CLEC10A |
| 10956-82 | GCSH | GCSH |
| 10966-1 | a2-HS-Glycoprotein | AHSG |
| 10976-44 | MUC1:region 3 | MUC1 |
| 10980-11 | ACES | ACHE |
| 11067-13 | Osteocalcin | BGLAP |
| 11081-1 | GPDA | GPD1 |
| 11101-18 | TLR4 | TLR4 |
| 11103-24 | HSP 27 | HSPB1 |
| 11104-13 | YKL-40 | CHI3L1 |
| 11105-171 | Alpha enolase | ENO1 |
| 11138-16 | RUNX3 | RUNX3 |
| 11140-56 | CO1A1:C-term propep | COL1A1 |
| 11171-25 | filamin A:CH2 | FLNA |
| 11185-145 | GCH1 | GCH1 |
| 11193-27 | HNF1A | HNF1A |
| 11201-19 | TRUA | PUS1 |
| 11202-70 | TBX5 | TBX5 |
| 11203-97 | KPYR | PKLR |
| 11208-15 | NAGPA | NAGPA |
| 11211-7 | TBCE | TBCE |
| 11218-84 | TPMT | TPMT |
| 11231-12 | ADXL | FDX2 |
| 11237-49 | PCOC1 | PCOLCE |
| 11239-49 | TMPS6 | TMPRSS6 |
| 11241-8 | ARLY | ASL |
| 11242-33 | TGM1 | TGM1 |
| 11245-43 | filamin A:CH1 | FLNA |
| 11247-20 | NAGS | NAGS |
| 11248-43 | HEM4 | UROS |
| 11257-1 | DHPR | QDPR |
| 11264-33 | XDH | XDH |
| 11266-8 | SELPL:ECD | SELPLG |
| 11270-17 | CHC10 | CHCHD10 |
| 11287-14 | Cytochrome b5 | CYB5A |
| 11297-54 | NOTC2 | NOTCH2 |
| 11300-32 | SORT | SORT1 |
| 11303-7 | SAMH1 | SAMHD1 |
| 11311-79 | RAG-1 | RAG1 |
| 11312-40 | PMS2 | PMS2 |
| 11313-100 | PHS | PCBD1 |

| | | |
|---|---|---|
| 11319-106 | MRE11 | MRE11 |
| 11336-9 | FANCF | FANCF |
| 11338-49 | BLK | BLK |
| 11347-9 | Transaldolase | TALDO1 |
| 11351-233 | NHEJ1 | NHEJ1 |
| 11352-42 | TITIN | TTN |
| 11356-19 | DGC14 | ESS2 |
| 11364-18 | MFN1 | MFN1 |
| 11368-32 | KAD2 | AK2 |
| 11370-20 | INP5E | INPP5E |
| 11381-56 | PRPS1 | PRPS1 |
| 11383-41 | Keratin 7 | KRT7 |
| 11391-69 | MVK | MVK |
| 11396-39 | DNAI1 | DNAI1 |
| 11406-82 | ACAD8 | ACAD8 |
| 11424-4 | FAAA | FAH |
| 11432-11 | PQBP1 | PQBP1 |
| 11436-6 | AINX | INA |
| 11440-58 | SOCS-3 | SOCS3 |
| 11441-11 | PYGL | PYGL |
| 11448-34 | GALK1 | GALK1 |
| 11450-110 | Protein disulfide-isom | P4HB |
| 11457-53 | GALE | GALE |
| 11516-7 | FABPL | FABP1 |
| 11530-37 | HEM3 | HMBS |
| 11537-12 | TFR2 | TFR2 |
| 11538-216 | DCMC | MLYCD |
| 11540-37 | FOXO3A | FOXO3 |
| 11616-9 | HSF1 | HSF1 |
| 11617-1 | LFA-1 alpha-L chain | ITGAL |
| 11696-7 | RABP2 | CRABP2 |
| 11699-16 | TP4A2 | PTP4A2 |
| 11709-29 | CPT1B | CPT1B |
| 11814-29 | Met | MET |
| 11816-84 | JAK2 | JAK2 |
| 11825-27 | PRGC1 | PPARGC1A |
| 11830-48 | SHP-2 | PTPN11 |
| 11833-83 | FRDA | FXN |
| 11838-130 | FA38A | PIEZO1 |
| 11949-25 | EGF:CD | EGF |
| 12008-3 | CD7 | CD7 |
| 12014-19 | PTPS | PTS |
| 12016-60 | CBL | CBL |
| 12020-39 | PMGE | BPGM |
| 12022-12 | SMAD4 | SMAD4 |
| 12030-82 | Desmin | DES |
| 12034-28 | CAP 1 | CAP1 |

| | | |
|---|---|---|
| 12046-51 | TADBP | TARDBP |
| 12077-32 | Myostatin | MSTN |
| 12333-87 | RPIA | RPIA |
| 12341-8 | DUS6 | DUSP6 |
| 12347-29 | CCM2 | CCM2 |
| 12348-46 | SYSM | SARS2 |
| 12351-25 | STAT1 | STAT1 |
| 12361-102 | RRAS2 | RRAS2 |
| 12367-52 | MACOI | MACO1 |
| 12378-71 | TPSN | TAPBP |
| 12381-26 | CBR1 | CBR1 |
| 12382-2 | DDX58 | DDX58 |
| 12387-7 | PDLI4 | PDLIM4 |
| 12395-86 | SYDM | DARS2 |
| 12396-19 | HIBCH | HIBCH |
| 12398-15 | PAX4 | PAX4 |
| 12400-25 | UBE2T | UBE2T |
| 12409-90 | RAB7B | RAB7B |
| 12420-10 | GPD1L | GPD1L |
| 12427-8 | MPIP2 | CDC25B |
| 12444-39 | NR5A2 | NR5A2 |
| 12446-49 | GST A1-1 | GSTA1 |
| 12456-5 | F263 | PFKFB3 |
| 12459-13 | PKHA1 | PLEKHA1 |
| 12466-7 | ROA1 | HNRNPA1 |
| 12469-19 | MARE1 | MAPRE1 |
| 12488-9 | MCTS1 | MCTS1 |
| 12501-10 | TBCA | TBCA |
| 12504-26 | LMOD1 | LMOD1 |
| 12507-16 | IP3KC | ITPKC |
| 12509-115 | COMD1 | COMMD1 |
| 12510-3 | STAP1 | STAP1 |
| 12521-3 | CDN2C | CDKN2C |
| 12527-50 | THA | THRA |
| 12529-32 | FKBP6 | FKBP6 |
| 12538-19 | RGS7 | RGS7 |
| 12574-36 | Endothelin 2 | EDN2 |
| 12580-7 | PSB5 | PSMB5 |
| 12585-39 | ERCC1 | ERCC1 |
| 12604-16 | SCMH1 | SCMH1 |
| 12612-37 | PSB1 | PSMB1 |
| 12620-3 | Septin-11 | SEPTIN11 |
| 12650-43 | GNAI3 | GNAI3 |
| 12651-21 | PDK2 | PDK2 |
| 12687-2 | DECR | DECR1 |
| 12700-9 | ACLY | ACLY |
| 12705-9 | HERC1 | HERC1 |

| | | |
|---|---|---|
| 12712-9 | HM20A | HMG20A |
| 12720-71 | UBQL4 | UBQLN4 |
| 12724-81 | CIRBP | CIRBP |
| 12726-3 | RFXAP | RFXAP |
| 12738-43 | NISCH | NISCH |
| 12740-55 | FEV | FEV |
| 12756-3 | BACH2 | BACH2 |
| 12825-18 | DP13A | APPL1 |
| 12869-68 | DDX6 | DDX6 |
| 12873-11 | LEUK | SPN |
| 12895-28 | DGKB | DGKB |
| 12904-180 | cubilin | CUBN |
| 12931-16 | MCR | NR3C2 |
| 12954-71 | PPARa | PPARA |
| 12957-62 | UCR6 | UQCRB |
| 12960-9 | HXK4 | GCK |
| 12988-49 | EWS | EWSR1 |
| 13027-20 | CBX7 | CBX7 |
| 13032-1 | BECN1 | BECN1 |
| 13053-6 | S6A14 | SLC6A14 |
| 13067-5 | KGP1B | PRKG1 |
| 13085-18 | GLP1R:CD | GLP1R |
| 13088-397 | BTC | BTC |
| 13089-6 | HIF-1a | HIF1A |
| 13090-17 | S100A6 | S100A6 |
| 13098-93 | VEGF-D | VEGFD |
| 13105-7 | SNP25 | SNAP25 |
| 13112-179 | FSTL1 | FSTL1 |
| 13113-7 | Osteopontin | SPP1 |
| 13119-26 | protein Z inhibitor | SERPINA10 |
| 13126-52 | DSC2 | DSC2 |
| 13129-40 | LDLR | LDLR |
| 13130-150 | HXK2 | HK2 |
| 13131-5 | HXK1 | HK1 |
| 13236-25 | WNT3A | WNT3A |
| 13381-49 | B4GT1 | B4GALT1 |
| 13384-110 | FUMH | FH |
| 13388-57 | NEC1 | PCSK1 |
| 13392-13 | AT1B1 | ATP1B1 |
| 13405-61 | ISK2 | SPINK2 |
| 13451-2 | FNDC4 | FNDC4 |
| 13465-5 | CCP1 | RCAN1 |
| 13470-43 | PTH1R | PTH1R |
| 13484-69 | CO1A1:N-term propep | COL1A1 |
| 13486-9 | PKD2:CD 1 | PKD2 |
| 13492-44 | HSPA9B | HSPA9 |
| 13499-30 | Coagulation Factor VIII | F8 |

| | | |
|---|---|---|
| 13510-7 | AT2A3 | ATP2A3 |
| 13556-28 | 5HT2A | HTR2A |
| 13576-15 | Glutathione S-transfer | GSTP1 |
| 13604-27 | NEUL1 | NEURL1 |
| 13609-11 | GTF2I | GTF2I |
| 13614-6 | CREB-binding protein | CREBBP |
| 13628-58 | GRB14 | GRB14 |
| 13634-209 | PIR | PIR |
| 13654-1 | ROCK2 | ROCK2 |
| 13655-34 | Nucleolin | NCL |
| 13657-2 | PNKP | PNKP |
| 13665-35 | PP2A, subunit B | PPP2R3A |
| 13669-6 | FGFR-3:ECD | FGFR3 |
| 13676-46 | Inhibin bB chain | INHBB |
| 13678-169 | Factor D | CFD |
| 13698-28 | K319L | KIAA0319L |
| 13700-10 | annexin II | ANXA2 |
| 13704-5 | HMCS2 | HMGCS2 |
| 13717-15 | FCN2 | FCN2 |
| 13726-4 | B7 | CD80 |
| 13730-18 | CATC | CTSC |
| 13732-79 | Cardiotrophin-1 | CTF1 |
| 13733-5 | IL-12 p40 | IL12B |
| 13741-36 | IGFBP-1 | IGFBP1 |
| 13745-10 | PKD2:CD 4 | PKD2 |
| 13941-82 | SYFM | FARS2 |
| 13967-14 | Thioredoxin reductase | TXNRD1 |
| 13977-28 | BARD1 | BARD1 |
| 13978-122 | Tbx3 | TBX3 |
| 13990-1 | pyruvate carboxylase | PC |
| 14005-2 | CHD7 | CHD7 |
| 14006-36 | GNMT | GNMT |
| 14025-18 | 4-1BB | TNFRSF9 |
| 14030-21 | OX40 Ligand | TNFSF4 |
| 14031-18 | FGF7 | FGF7 |
| 14035-13 | TIAM1 | TIAM1 |
| 14047-78 | BDNF | BDNF |
| 14051-54 | FOXC2 | FOXC2 |
| 14052-26 | UN13A | UNC13A |
| 14060-67 | NCF4 | NCF4 |
| 14061-48 | sRANKL | TNFSF11 |
| 14066-49 | MAGI2 | MAGI2 |
| 14067-6 | PKP2 | PKP2 |
| 14069-61 | Carbonic Anhydrase IV | CA4 |
| 14094-29 | HB-EGF | HBEGF |
| 14105-5 | Kallistatin | SERPINA4 |
| 14110-200 | ON | SPARC |

| | | |
|---|---|---|
| 14131-37 | EFNB2:ECD | EFNB2 |
| 14136-234 | C1QR1 | CD93 |
| 14158-17 | Annexin V | ANXA5 |
| 14284-23 | E2AK4 | EIF2AK4 |
| 14645-253 | GSTA4 | GSTA4 |
| 14756-29 | sICAM-2 | ICAM2 |
| 15295-81 | IGF-II:Mature | IGF2 |
| 15299-102 | MESD2 | MESD |
| 15303-63 | S100A5 | S100A5 |
| 15324-58 | Ferritin light chain | FTL |
| 15339-32 | COF2 | CFL2 |
| 15343-337 | Kininogen, HMW, Two | KNG1 |
| 15346-31 | IFN-g | IFNG |
| 15361-37 | ANKR1 | ANKRD1 |
| 15363-32 | Apo A-V | APOA5 |
| 15364-101 | Apo C-I | APOC1 |
| 15383-200 | Endothelin 3 | EDN3 |
| 15384-15 | KLOTHO | KL |
| 15385-116 | FABP2 | FABP2 |
| 15386-7 | FABPA | FABP4 |
| 15388-24 | FcRIIIa | FCGR3A |
| 15390-3 | Galanin | GAL |
| 15391-114 | GAS-6 | GAS6 |
| 15395-15 | GST M1-1 | GSTM1 |
| 15398-2 | HERV1 | GFER |
| 15413-3 | LCAT | LCAT |
| 15414-316 | LDHA | LDHA |
| 15431-31 | OTCase | OTC |
| 15433-4 | p60-Src | SRC |
| 15434-5 | PNOC | PNOC |
| 15435-4 | PNP | PNP |
| 15440-57 | NEC2 | PCSK2 |
| 15441-6 | SAP3 | GM2A |
| 15447-45 | Sorbitol dehydrogenas | SORD |
| 15448-47 | SQSTM | SQSTM1 |
| 15449-33 | TIM-4 | TIMD4 |
| 15452-5 | 5'-Nucleotidase | NT5E |
| 15453-3 | a1-Microglobulin | AMBP |
| 15457-14 | Aminopeptidase N | ANPEP |
| 15460-9 | CD26 | DPP4 |
| 15467-10 | CTHR1 | CTHRC1 |
| 15470-11 | Hexosaminidase B | HEXB |
| 15475-4 | PLTP | PLTP |
| 15476-6 | REG3G | REG3G |
| 15482-12 | A2ML1 | A2ML1 |
| 15486-126 | ABP1 | AOC1 |
| 15503-20 | Lefty-A | LEFTY2 |

| | | |
|---|---|---|
| 15509-2 | NAG | NAGLU |
| 15513-108 | Prostasin | PRSS8 |
| 15514-26 | Pseudocholinesterase | BCHE |
| 15515-2 | SAA | SAA1 |
| 15523-9 | HEM2 | ALAD |
| 15524-30 | PGAM2 | PGAM2 |
| 15526-33 | GSHB | GSS |
| 15527-90 | DLDH | DLD |
| 15530-33 | EphB4 | EPHB4 |
| 15535-3 | Marapsin | PRSS27 |
| 15540-6 | Vimentin | VIM |
| 15545-13 | Calcineurin B a | PPP3R1 |
| 15559-5 | ANTR2 | ANTXR2 |
| 15562-24 | BGLR | GUSB |
| 15567-2 | CD3-zeta | CD247 |
| 15570-99 | Complement receptor | CR2 |
| 15585-304 | fibulin 5 | FBLN5 |
| 15588-17 | galactosidase, alpha | GLA |
| 15589-1 | Gc-Globulin, Mixed Ty | GC |
| 15591-28 | Glutathione peroxidas | GPX1 |
| 15594-47 | HTRA1 | HTRA1 |
| 15603-20 | Integrin alpha-2 | ITGA2 |
| 15604-18 | JNK2 | MAPK9 |
| 15606-19 | Keratin 19 | KRT19 |
| 15608-5 | KS6B1 | RPS6KB1 |
| 15626-223 | Perlecan | HSPG2 |
| 15633-6 | RBP | RBP4 |
| 15635-4 | SMOC2 | SMOC2 |
| 15666-21 | BMP-2 | BMP2 |
| 15667-39 | BMP-4 | BMP4 |
| 15669-7 | BRAF1 | BRAF |
| 15692-300 | NODAL | NODAL |
| 16035-8 | VEGF sR3 | FLT4 |
| 16043-30 | SHC1:PID | SHC1 |
| 16057-6 | IGF-II receptor | IGF2R |
| 16288-17 | EPHA4 | EPHA4 |
| 16297-14 | ROBO4 | ROBO4 |
| 16318-12 | ALK-1 | ACVRL1 |
| 16320-139 | CD320 | CD320 |
| 16323-8 | NRX3A | NRXN3 |
| 16536-3 | HMGA1 | HMGA1 |
| 16551-14 | STA5B | STAT5B |
| 16588-10 | BiP | HSPA5 |
| 16593-3 | FADD | FADD |
| 16596-25 | GLRX3 | GLRX3 |
| 16597-11 | GLRX5 | GLRX5 |
| 16612-28 | SDC3 | SDC3 |

| | | |
|---|---|---|
| 16616-137 | ENOB | ENO3 |
| 16618-7 | CD69 | CD69 |
| 16754-40 | CSKP | CASK |
| 16768-3 | PLAK | JUP |
| 16803-4 | CALB2 | CALB2 |
| 16809-1 | NDKM | NME4 |
| 16814-13 | SLIM 1 | FHL1 |
| 16828-8 | Collagen a1(VI) | COL6A1 |
| 16850-5 | RGS5 | RGS5 |
| 16851-50 | SCO2 | SCO2 |
| 16865-62 | RBP56 | TAF15 |
| 16890-37 | ATL1 | ADAMTSL1 |
| 16914-104 | sCD14 | CD14 |
| 16915-153 | SEM4A | SEMA4A |
| 16916-19 | SLIK6 | SLITRK6 |
| 16926-44 | Alkaline phosphatase, | ALPL |
| 16927-9 | Coagulation factor XIII | F13B\|F13A1 |
| 17145-1 | S100A8/S100A9 | S100A9\|S100A8 |
| 17150-8 | HSP 10 | HSPE1 |
| 17161-1 | OST48 | DDOST |
| 17205-21 | RAB5A | RAB5A |
| 17333-20 | ACADM | ACADM |
| 17341-89 | THIC | ACAT2 |
| 17350-13 | CHM2B | CHMP2B |
| 17362-5 | VHL | VHL |
| 17377-1 | Aldose reductase-like ( | AKR1C3 |
| 17384-110 | K6PF | PFKM |
| 17393-13 | SCAD | ACADS |
| 17398-55 | HO-1 | HMOX1 |
| 17403-14 | acyl-Coenzyme A dehy | ACADSB |
| 17411-55 | UFD1 | UFD1 |
| 17435-43 | ETFA | ETFA |
| 17447-52 | SFRP4 | SFRP4 |
| 17451-13 | BIM | BCL2L11 |
| 17453-34 | Ceruloplasmin | CP |
| 17455-42 | FOLR1 | FOLR1 |
| 17460-51 | Mx1 | MX1 |
| 17495-141 | SIR3 | SIRT3 |
| 17509-6 | HPPD | HPD |
| 17513-11 | ANX11 | ANXA11 |
| 17514-48 | RAB21 | RAB21 |
| 17672-184 | Gastric intrinsic factor | CBLIF |
| 17682-1 | CD46 | CD46 |
| 17685-9 | Apo A-IV | APOA4 |
| 17691-1 | TPP1 | TPP1 |
| 17702-53 | UGT 1A1 | UGT1A1 |
| 17706-4 | PPR1A | PPP1R1A |

| | | |
|---|---|---|
| 17721-82 | GAS7 | GAS7 |
| 17734-13 | a-endosulfine | ENSA |
| 17737-7 | IVD | IVD |
| 17739-1 | HCDH | HADH |
| 17742-2 | RRAS | RRAS |
| 17746-77 | FIS1 | FIS1 |
| 17747-45 | TRAF1 | TRAF1 |
| 17758-79 | DCXR | DCXR |
| 17764-108 | RHOC | RHOC |
| 17766-5 | NCF-1 | NCF1 |
| 17768-50 | HAOX1 | HAO1 |
| 17772-7 | UBD | UBD |
| 17783-9 | MMAB | MMAB |
| 17787-1 | Enoyl-CoA hydratase | ECHS1 |
| 17792-158 | SSDH | ALDH5A1 |
| 17794-6 | Phosphomannomutas | PMM2 |
| 17799-9 | 6PGL | PGLS |
| 17804-102 | PNPO | PNPO |
| 17843-30 | PPCS | PPCS |
| 17850-42 | KLF4 | KLF4 |
| 17857-6 | ACADL | ACADL |
| 18158-45 | Caspase-8 | CASP8 |
| 18162-167 | IRAK4 | IRAK4 |
| 18170-46 | P5CR1 | PYCR1 |
| 18182-24 | PCKGC | PCK1 |
| 18183-3 | ARH | LDLRAP1 |
| 18185-118 | ALDOB | ALDOB |
| 18187-16 | ACY2 | ASPA |
| 18188-12 | GATM | GATM |
| 18214-2 | GSH0 | GCLM |
| 18216-22 | IL-11 RA | IL11RA |
| 18226-148 | COX5A | COX5A |
| 18241-18 | HEM6 | CPOX |
| 18291-8 | p21 | CDKN1A |
| 18295-102 | GRHPR | GRHPR |
| 18297-8 | RASM | MRAS |
| 18309-18 | CISY | CS |
| 18319-7 | ODPX | PDHX |
| 18324-61 | SUOX | SUOX |
| 18331-3 | GLCM | GBA |
| 18347-15 | Laminin-2 | LAMC1\|LAMB1\|LAMA2 |
| 18376-19 | Myosin light chain 1 | MYL3 |
| 18380-78 | Albumin | ALB |
| 18381-16 | ALDH-E2 | ALDH2 |
| 18382-109 | Catechol O-methyltrar | COMT |
| 18396-10 | AES | TLE5 |
| 18398-1 | AK1D1 | AKR1D1 |

| | | |
|---|---|---|
| 18415-16 | ARL6 | ARL6 |
| 18429-10 | Cadherin E | CDH1 |
| 18830-1 | Omentin | ITLN1 |
| 18831-6 | LRIG1 | LRIG1 |
| 18870-1 | CD3E | CD3E |
| 18874-66 | CEBPA | CEBPA |
| 18880-81 | Collagen Type III | COL3A1 |
| 18890-227 | Fibrinogen B | FGB |
| 18891-98 | GBP2 | GBP2 |
| 18892-48 | GPC4 | GPC4 |
| 18897-31 | HDAC2 | HDAC2 |
| 18900-37 | H-ras (WT) | HRAS |
| 18901-26 | HS71B | HSPA1B |
| 18916-25 | Inosine triphosphatase | ITPA |
| 18921-30 | PHEX | PHEX |
| 18925-24 | Proteasome subunit al | PSMA5 |
| 18931-40 | SLIT3 | SLIT3 |
| 18934-50 | Tissue transglutaminas | TGM2 |
| 18938-3 | VLDLR | VLDLR |
| 18950-13 | RAC2 | RAC2 |
| 19114-8 | KITM | TK2 |
| 19119-10 | DDIT3 | DDIT3 |
| 19120-33 | OXDA | DAO |
| 19121-3 | UBQL2 | UBQLN2 |
| 19127-1 | HSPB6 | HSPB6 |
| 19136-22 | MMSA | ALDH6A1 |
| 19144-9 | ITF2 | TCF4 |
| 19168-71 | PPIP1 | PSTPIP1 |
| 19176-27 | FA49B | CYRIB |
| 19187-21 | STABP | STAMBP |
| 19197-95 | THIL | ACAT1 |
| 19209-6 | IKB-alpha | NFKBIA |
| 19215-7 | NDRG1 | NDRG1 |
| 19230-12 | GSTT1 | GSTT1 |
| 19233-75 | ATOX1 | ATOX1 |
| 19238-12 | GLNA | GLUL |
| 19258-24 | GCDH | GCDH |
| 19262-219 | ACADV | ACADVL |
| 19273-3 | Glutathione reductase | GSR |
| 19274-80 | Alpha-1-syntrophin | SNTA1 |
| 19278-19 | RAB1B | RAB1B |
| 19289-29 | DCUP | UROD |
| 19290-5 | HPRT | HPRT1 |
| 19291-2 | CASQ2 | CASQ2 |
| 19293-6 | VP26A | VPS26A |
| 19296-51 | MLRA | MYL7 |
| 19297-4 | G6PD | G6PD |

| | | |
|---|---|---|
| 19303-64 | PT117 | PET117 |
| 19364-163 | PCNA | PCNA |
| 19365-11 | BCAT2 | BCAT2 |
| 19366-8 | PEX26 | PEX26 |
| 19561-216 | PLXD1 | PLXND1 |
| 19564-61 | IRF4 | IRF4 |
| 19568-17 | IL-15 | IL15 |
| 19570-12 | FGF-8 | FGF8 |
| 19602-36 | jun-D | JUND |
| 19637-9 | CRH | CRH |
| 19638-9 | Dynorphin A (1-17) | PDYN |
| 19639-53 | IAPP | IAPP |
| 19640-2 | Parathyroid Hormone: | PTH |
| 19751-21 | ADA | ADA |
| 19768-13 | Cystatin B | CSTB |
| 19823-75 | SIR1 | SIRT1 |
| 20073-22 | K-ras | KRAS |
| 20103-176 | N-glycosylase/DNA lya | OGG1 |
| 20107-11 | MYL4 | MYL4 |
| 20116-30 | OXSR1 | OXSR1 |
| 20128-1 | CREB1 | CREB1 |
| 20134-27 | MARCO | MARCO |
| 20165-4 | DPYS | DPYS |
| 20185-44 | ODP2 | DLAT |
| 20187-10 | Integrin aVb3 | ITGB3\|ITGAV |
| 20191-13 | Integrin aVb8 | ITGAV\|ITGB8 |
| 20195-13 | IFIH1 | IFIH1 |
| 20203-45 | C1s | C1S |
| 20211-75 | GBP5 | GBP5 |
| 20215-45 | Integrin aVb1 | ITGB1\|ITGAV |
| 20217-26 | Lamin-B2 | LMNB2 |
| 20225-119 | STA5A | STAT5A |
| 20241-9 | SNP23 | SNAP23 |
| 20243-26 | Profilin-1 | PFN1 |
| 20245-13 | Retinoic acid receptor | RXRA |
| 20374-41 | COQ9 | COQ9 |
| 20378-110 | LZTL1 | LZTFL1 |
| 20425-12 | HHEX | HHEX |
| 20428-5 | PHOP1 | PHOSPHO1 |
| 20458-22 | NDUA2 | NDUFA2 |
| 20516-11 | SCN3B | SCN3B |
| 20528-23 | CD68 | CD68 |
| 20533-39 | IL-35 | IL12A\|EBI3 |
| 20534-6 | SCRB1 | SCARB1 |
| 20542-47 | CALCR | CALCR |
| 20553-2 | P3IP1 | PIK3IP1 |
| 20564-53 | THY1 | THY1 |

| | | |
|---|---|---|
| 20572-6 | MER | MERTK |
| 20581-42 | GLP1R:ECD | GLP1R |
| 20590-13 | NPY | NPY |
| 20931-156 | AUHM | AUH |
| 20953-34 | MMAC | MMACHC |
| 20996-107 | SDHF1 | SDHAF1 |
| 21107-5 | CIA30 | NDUFAF1 |
| 21120-3 | MIMIT | NDUFAF2 |
| 21126-27 | GFPT1 | GFPT1 |
| 21207-1 | COASY | COASY |
| 21227-18 | SULT 1A1*2 | SULT1A1 |
| 21239-31 | CYH1 | CYTH1 |
| 21240-6 | GCST | AMT |
| 21247-16 | METK1 | MAT1A |
| 21269-198 | HYES | EPHX2 |
| 21271-53 | AK1C2 | AKR1C2 |
| 21280-13 | T22D3 | TSC22D3 |
| 21330-13 | SYYM | YARS2 |
| 21355-4 | USF1 | USF1 |
| 21384-2 | Alpha-L-fucosidase I | FUCA1 |
| 21393-62 | SAHH | AHCY |
| 21430-4 | cPLA2-alpha | PLA2G4A |
| 21441-20 | gp75 | TYRP1 |
| 21452-3 | DNM1L | DNM1L |
| 21477-105 | MAEA | MAEA |
| 21478-20 | SMAD5 | SMAD5 |
| 21483-155 | KAP0 | PRKAR1A |
| 21487-20 | BL1S6 | BLOC1S6 |
| 21491-7 | VAP-1 | AOC3 |
| 21510-24 | GRK5 | GRK5 |
| 21513-1 | Caspase-7 | CASP7 |
| 21514-1 | GP1BB | GP1BB |
| 21533-51 | LSP1 | LSP1 |
| 21552-9 | ACOX1 | ACOX1 |
| 21572-91 | XYLT2 | XYLT2 |
| 21579-35 | MYPC3 | MYBPC3 |
| 21599-6 | CGL | CTH |
| 21643-8 | RS20 | RPS20 |
| 21651-9 | TXN4A | TXNL4A |
| 21660-4 | SAR1B | SAR1B |
| 21685-29 | DCC | DCC |
| 21703-31 | Activin RIA | ACVR1 |
| 21705-33 | METRL | METRNL |
| 21724-22 | GALNS | GALNS |
| 21739-7 | GLCNE | GNE |
| 21766-50 | SPHM | SGSH |
| 21796-43 | Glutathione peroxidas | GPX3 |

| | | |
|---|---|---|
| 21802-53 | NEIL1 | NEIL1 |
| 21814-13 | MMAD | MMADHC |
| 21817-5 | MYL9 | MYL9 |
| 21833-6 | sirtuin | SIRT6 |
| 21856-59 | KHK | KHK |
| 21875-31 | ZFAN3 | ZFAND3 |
| 21901-14 | Integrin a3b1 | F11 |
| 21903-6 | Integrin aLb2 | ITGA3\|ITGB1 |
| 21905-10 | Integrin alpha-2/ b1 | ITGB2\|ITGAL |
| 2190-55 | Coagulation Factor XI | ITGB1\|ITGA2 |
| 21909-2 | Integrin a5b1 | ITGA5\|ITGB1 |
| 21911-17 | NDP | NDP |
| 21967-20 | DHI1 | HSD11B1 |
| 21969-5 | DPP6 | DPP6 |
| 21976-4 | IKK-gamma | IKBKG |
| 21981-2 | Integrin alpha-M | ITGAM |
| 21987-76 | LPL | LPL |
| 21995-20 | NOS | NOS3 |
| 22003-4 | Paraoxonase-2 | PON2 |
| 22005-8 | PDP1 | PDP1 |
| 22009-1 | PPM1B | PPM1B |
| 2201-17 | Endostatin | COL18A1 |
| 22019-21 | SH2B3 | SH2B3 |
| 22043-174 | HSP70 protein 2 | HSPA2 |
| 22045-8 | SUMO4 | SUMO4 |
| 22047-46 | CO5A1 | COL5A1 |
| 22049-24 | 14-3-3 eta | YWHAH |
| 22057-9 | AL1B1 | ALDH1B1 |
| 22075-16 | ATF3 | ATF3 |
| 22078-4 | ATTY | TAT |
| 22088-3 | BTG1 | BTG1 |
| 22091-14 | BUP1 | UPB1 |
| 22116-9 | CREM | CREM |
| 2212-69 | tPA | PLAT |
| 22148-135 | FOXP3 | FOXP3 |
| 22154-37 | GATD1 | GATAD1 |
| 22383-21 | MITF | MITF |
| 22394-8 | TCF21 | TCF21 |
| 22395-7 | Tristetraproline | ZFP36 |
| 22397-2 | XBP1 | XBP1 |
| 22405-61 | PCKGM | PCK2 |
| 22485-1 | KCTD7 | KCTD7 |
| 22488-17 | KLF9 | KLF9 |
| 22505-24 | NAKD2 | NADK2 |
| 22508-2 | NDUBA | NDUFB10 |
| 22509-9 | NDUF5 | NDUFAF5 |
| 22510-6 | NF2L2 | NFE2L2 |

| | | |
|---|---|---|
| 22526-81 | PDHB | PDHB |
| 22527-4 | PDLI3 | PDLIM3 |
| 22529-31 | PHF6 | PHF6 |
| 22531-44 | PITX2 | PITX2 |
| 22540-129 | PSB7 | PSMB7 |
| 22544-10 | RA51C | RAD51C |
| 22569-55 | TGFR-1 | TGFBR1 |
| 22584-2 | LRP6 | LRP6 |
| 22773-69 | SDHB | SDHB |
| 22776-40 | SHP | NR0B2 |
| 2278-61 | TIMP-2 | TIMP2 |
| 22800-24 | TELT | TCAP |
| 22808-59 | THB | THRB |
| 22818-4 | TXTP | SLC25A1 |
| 22821-50 | UNG | UNG |
| 22847-18 | STK4 | STK4 |
| 22953-85 | CEBPB | CEBPB |
| 22958-6 | DEST | DSTN |
| 22969-12 | MCP-3 | CCL7 |
| 22978-13 | HMG-2 | HMGB2 |
| 23022-5 | RhoGDI | GDI1 |
| 23024-25 | SAR1A | SAR1A |
| 23038-63 | UNC5B | UNC5B |
| 23039-56 | MEOX2 | MEOX2 |
| 23173-3 | TIMP-1 | TIMP1 |
| 23213-21 | CK070 | CFAP300 |
| 23254-31 | NDUF3 | NDUFAF3 |
| 23263-9 | KCD15 | KCTD15 |
| 23275-9 | SSPN | SSPN |
| 23294-19 | MLX | MLX |
| 2330-2 | SDF-1 | CXCL12 |
| 23302-19 | AKTS1 | AKT1S1 |
| 23314-46 | CC103 | CCDC103 |
| 2333-72 | TGF-b1 | TGFB1 |
| 23352-9 | DTBP1 | DTNBP1 |
| 23369-17 | PAHX | PHYH |
| 23393-56 | ABHD5 | ABHD5 |
| 23404-16 | MD2L2 | MAD2L2 |
| 23418-66 | CCD92 | CCDC92 |
| 23555-11 | RAD | RRAD |
| 23557-110 | CAPZB | CAPZB |
| 23567-37 | GNAQ | GNAQ |
| 23568-41 | MEIS2 | MEIS2 |
| 23643-22 | DHB4 | HSD17B4 |
| 23652-15 | SOX | PIPOX |
| 23654-6 | ALG2 | ALG2 |
| 23658-1 | SYNM | NARS2 |

| | | |
|---|---|---|
| 23666-35 | PUR9 | ATIC |
| 23672-10 | PSMD6 | PSMD6 |
| 23673-9 | ODB2 | DBT |
| 23680-1 | MCPIP | ZC3H12A |
| 23694-3 | ASNS | ASNS |
| 23703-8 | PDLI5 | PDLIM5 |
| 23903-3 | GOT2 | GOT2 |
| 23915-18 | Rab-7 | RAB7A |
| 23981-172 | CBPA1 | CPA1 |
| 24023-35 | sEPCR | PROCR |
| 24050-26 | GSK-3 beta | GSK3B |
| 2418-55 | Apo E | APOE |
| 24211-25 | HMGCL | HMGCL |
| 24215-8 | PRRX1 | PRRX1 |
| 24221-3 | PPAR gamma | PPARG |
| 24223-5 | GST2 | GSTT2 |
| 24252-85 | CL065 | MTRFR |
| 24263-6 | OCLN | OCLN |
| 24276-171 | TTC25 | ODAD4 |
| 24278-4 | UBP3 | USP3 |
| 2436-49 | CXCL16, soluble | CXCL16 |
| 24407-31 | KIF3C | KIF3C |
| 24414-3 | Glycogen phosphoryla | PYGB |
| 24419-3 | MOCOS | MOCOS |
| 24426-15 | CTNA1 | CTNNA1 |
| 24459-15 | COPB2 | COPB2 |
| 24463-1 | NDRG2 | NDRG2 |
| 24469-10 | LPIN1 | LPIN1 |
| 24487-95 | BL1S3 | BLOC1S3 |
| 24646-47 | SPYA | AGXT |
| 24651-1 | RTN1 | RTN1 |
| 24659-6 | DJB13 | DNAJB13 |
| 24671-15 | ASC | PYCARD |
| 24676-105 | CU059 | CFAP298 |
| 24681-2 | SERB | CCL20 |
| 24682-35 | CYBR1 | PSPH |
| 2468-62 | MIP-3a | CYBRD1 |
| 24689-9 | PTRF | CAVIN1 |
| 24694-158 | NHERF | SLC9A3R1 |
| 24701-21 | DTNA | DTNA |
| 2474-54 | SAP | APCS |
| 2475-1 | SCF sR | KIT |
| 24898-39 | SCFD1 | SCFD1 |
| 24902-84 | GEPH | GPHN |
| 24910-18 | NonO protein | NONO |
| 24940-3 | IKK-beta | IKBKB |
| 24958-3 | RFX5 | RFX5 |

| | | |
|---|---|---|
| 24960-48 | HIP1R | HIP1R |
| 25033-194 | TER ATPase | VCP |
| 25043-74 | SCOT | OXCT1 |
| 25051-104 | FTO | FTO |
| 25060-18 | MAOX | ME1 |
| 25061-8 | ELMO2 | ELMO2 |
| 25066-32 | GLYG2 | GYG2 |
| 25076-2 | Selenium-binding prot | SELENBP1 |
| 25088-42 | PHKA1 | PHKA1 |
| 25089-21 | KRIP-1 | TRIM28 |
| 25100-11 | CMIP | CMIP |
| 25105-70 | EIF2A | EIF2A |
| 25209-15 | ATNG | FXYD2 |
| 25233-2 | MPI | MPI |
| 2524-56 | HMG-1 | HMGB1 |
| 25253-17 | GNAS | GNAS |
| 25264-102 | SHIP | INPP5D |
| 25283-2 | CTNA3 | CTNNA3 |
| 25288-16 | FMO3 | FMO3 |
| 25291-27 | PKHM2 | PLEKHM2 |
| 25292-4 | EST3 | CES3 |
| 25297-11 | SPD2B | SH3PXD2B |
| 25300-39 | MALT1 | MALT1 |
| 25308-8 | LIMK1 | LIMK1 |
| 25460-36 | IRF5 | IRF5 |
| 25463-3 | KAP3 | PRKAR2B |
| 25464-1 | CP2CJ | CYP2C19 |
| 2570-72 | IGFBP-2 | IGFBP2 |
| 2571-12 | IGFBP-3 | IGFBP3 |
| 2578-67 | MCP-1 | CCL2 |
| 2579-17 | MMP-9 | MMP9 |
| 2580-83 | Myeloperoxidase | MPO |
| 2585-2 | PRL | PRL |
| 25880-14 | 5-Lipoxygenase | ALOX5 |
| 25886-11 | ABCD4 | ABCD4 |
| 25902-27 | DHE3 | GLUD1 |
| 25913-17 | FRIH | FTH1 |
| 25917-12 | Hexosaminidase A | HEXA |
| 25922-7 | LRP5 | LRP5 |
| 25938-13 | NGN3 | NEUROG3 |
| 25951-17 | PRAME | PRAME |
| 25962-51 | WNK1 | WNK1 |
| 2597-8 | VEGF | VEGFA |
| 2609-59 | Cystatin C | CST3 |
| 2611-72 | Dtk | TYRO3 |
| 2615-60 | Ephrin-A5 | EFNA5 |
| 2618-10 | ERBB4 | ERBB4 |

| | | |
|---|---|---|
| 2619-72 | GA733-1 protein | TACSTD2 |
| 2620-4 | gp130, soluble | IL6ST |
| 2622-18 | HO-2 | HMOX2 |
| 2631-50 | IL-10 Rb | IL10RB |
| 2632-5 | IL-12 Rb1 | IL12RB1 |
| 2633-52 | IL-13 Ra1 | IL13RA1 |
| 2634-2 | IL-2 sRg | IL2RG |
| 2636-10 | Lymphotoxin b R | LTBR |
| 2637-77 | Macrophage mannose | MRC1 |
| 2644-11 | PKC-A | PRKCA |
| 2645-54 | PKC-Z | PRKCZ |
| 2649-77 | sICAM-3 | ICAM3 |
| 2652-15 | suPAR | PLAUR |
| 2654-19 | TNF sR-I | TNFRSF1A |
| 2666-53 | Bone proteoglycan II | DCN |
| 2670-67 | CK-MM | CKM |
| 2677-1 | ERBB1 | EGFR |
| 2681-23 | HGF | HGF |
| 2682-68 | HSP 60 | HSPD1 |
| 2686-67 | IGFBP-6 | IGFBP6 |
| 2692-74 | NPS-PLA2 | PLA2G2A |
| 2695-25 | PECAM-1 | PECAM1 |
| 2697-7 | PF-4 | PF4 |
| 2700-56 | Protein S | PROS1 |
| 2704-74 | TACI | TNFRSF13B |
| 2706-69 | Thyroxine-Binding Glo | SERPINA7 |
| 2730-58 | MICA | MICA |
| 2731-29 | NADPH-P450 Oxidored | POR |
| 2750-3 | Apo A-I | APOA1 |
| 2753-2 | C1q | C1QC\|C1QB\|C1QA |
| 2754-50 | C3 | C3 |
| 2765-4 | GDF-11/8 | MSTN\|GDF11 |
| 2768-56 | Hemopexin | HPX |
| 2773-50 | IL-10 | IL10 |
| 2774-10 | IL-16 | IL16 |
| 2778-10 | IL-22 | IL22 |
| 2780-35 | Lactoferrin | LTF |
| 2788-55 | MMP-3 | MMP3 |
| 2789-26 | MMP-7 | MMP7 |
| 2790-54 | NAP-2 | PPBP |
| 2794-60 | SOD | SOD1 |
| 2796-62 | Fibrinogen | FGG\|FGB\|FGA |
| 2797-56 | Apo B | APOB |
| 2805-6 | ACE2 | ACE2 |
| 2811-27 | Angiopoietin-1 | ANGPT1 |
| 2813-11 | ART | AGRP |
| 2819-23 | Cadherin-5 | CDH5 |

| | | |
|---|---|---|
| 2827-23 | Fractalkine/CX3CL-1 | CX3CL1 |
| 2829-19 | IL-27 | EBI3\|IL27 |
| 2831-29 | Kallikrein 11 | KLK11 |
| 2834-54 | kallikrein 8 | KLK8 |
| 2836-68 | Lipocalin 2 | LCN2 |
| 2843-13 | SPINT2 | SPINT2 |
| 2855-49 | ERK-1 | MAPK3 |
| 2857-70 | Glucocorticoid recepto | NR3C1 |
| 2859-69 | HDAC8 | HDAC8 |
| 2864-2 | MEK1 | MAP2K1 |
| 2867-52 | PKB | AKT1 |
| 2869-68 | PKC-D | PRKCD |
| 2870-29 | RAC1 | RAC1 |
| 2871-73 | RAD51 | RAD51 |
| 2878-66 | YES | YES1 |
| 2900-53 | HCC-1 | CCL14 |
| 2906-55 | IL-4 | IL4 |
| 2925-9 | PAI-1 | SERPINE1 |
| 2942-50 | Cytochrome c | CYCS |
| 2943-5 | Cytochrome P450 3A4 | CYP3A4 |
| 2948-58 | Growth hormone rece | GHR |
| 2953-31 | Luteinizing hormone | LHB\|CGA |
| 2961-1 | Protein C | PROC |
| 2962-50 | PTHrP | PTHLH |
| 2967-8 | VCAM-1 | VCAM1 |
| 2970-60 | AREG | AREG |
| 2973-15 | CD36 ANTIGEN | CD36 |
| 2974-61 | contactin-1 | CNTN1 |
| 2975-19 | CTGF | CCN2 |
| 2976-58 | Desmoglein-1 | DSG1 |
| 2985-35 | Gro-a | CXCL1 |
| 2986-49 | Gro-g | CXCL3 |
| 2991-9 | IL-1 sRI | IL1R1 |
| 2992-59 | IL-17 sR | IL17RA |
| 2997-8 | JAM-B | JAM2 |
| 3000-66 | MBL | MBL2 |
| 3005-5 | PTP-1B | PTPN1 |
| 3009-3 | TGF-b R III | TGFBR3 |
| 3022-4 | CTLA-4 | CTLA4 |
| 3025-50 | bFGF | FGF2 |
| 3032-11 | FSH | FSHB\|CGA |
| 3033-57 | Galectin-2 | LGALS2 |
| 3034-1 | GFAP | GFAP |
| 3035-80 | IL-19 | IL19 |
| 3037-62 | IL-1b | IL1B |
| 3038-9 | I-TAC | CXCL11 |
| 3040-59 | MIP-1a | CCL3 |

| | | |
|---|---|---|
| 3044-3 | PARC | CCL18 |
| 3046-31 | resistin | RETN |
| 3049-61 | Trypsin | PRSS1 |
| 3050-7 | vWF | VWF |
| 3052-8 | Fas ligand, soluble | FASLG |
| 3054-3 | Haptoglobin, Mixed Ty | HP |
| 3055-54 | IL-4 sR | IL4R |
| 3059-50 | BAFF | TNFSF13B |
| 3065-65 | FGF-5 | FGF5 |
| 3066-12 | Galectin-3 | LGALS3 |
| 3070-1 | IL-2 | IL2 |
| 3073-51 | IL-18 BPa | IL18BP |
| 3074-6 | LBP | LBP |
| 3077-66 | Coagulation Factor Xa | F10 |
| 3078-1 | PlGF | PGF |
| 3079-62 | TIG2 | RARRES2 |
| 3115-64 | MK01 | MAPK1 |
| 3122-6 | SMAC | DIABLO |
| 3143-3 | sCD4 | CD4 |
| 3148-49 | Gro-b | CXCL2 |
| 3151-6 | IL-2 sRa | IL2RA |
| 3169-70 | IDUA | IDUA |
| 3171-57 | amyloid precursor pro | APP |
| 3172-28 | ARSB | ARSB |
| 3175-51 | ATS13 | ADAMTS13 |
| 3179-51 | Cathepsin A | CTSA |
| 3181-50 | Cathepsin S | CTSS |
| 3184-25 | Coagulation Factor VII | F7 |
| 3186-2 | C2 | C2 |
| 3189-61 | Enterokinase | TMPRSS15 |
| 3198-4 | IDS | IDS |
| 3204-2 | LKHA4 | LTA4H |
| 3206-4 | LYVE1 | LYVE1 |
| 3209-69 | MEPE | MEPE |
| 3212-30 | ASAH2 | ASAH2 |
| 3213-65 | Nidogen | NID1 |
| 3220-40 | RET | RET |
| 3221-54 | SARP-2 | SFRP1 |
| 3232-28 | TrATPase | ACP5 |
| 3283-21 | BGH3 | TGFBI |
| 3285-23 | C1r | C1R |
| 3294-55 | CFC1 | CFC1 |
| 3309-2 | FCG2A | FCGR2A |
| 3310-62 | FCG2B | FCGR2B |
| 3311-27 | FCG3B | FCGR3B |
| 3312-64 | FCGR1 | FCGR1A |
| 3316-58 | Heparin cofactor II | SERPIND1 |

| | | |
|---|---|---|
| 3317-33 | HTRA2 | HTRA2 |
| 3320-49 | IGFBP-7 | IGFBP7 |
| 3323-37 | LRP8 | LRP8 |
| 3327-27 | NET4 | NTN4 |
| 3332-57 | RGM-C | HJV |
| 3336-50 | TFPI | TFPI |
| 3339-33 | TSP2 | THBS2 |
| 3341-33 | ABL1 | ABL1 |
| 3343-1 | Aminoacylase-1 | ACY1 |
| 3344-60 | Antithrombin III | SERPINC1 |
| 3347-9 | BARK1 | GRK2 |
| 3350-53 | CAMK2A | CAMK2A |
| 3359-11 | CDK8/cyclin C | CDK8\|CCNC |
| 3360-50 | Chk2 | CHEK2 |
| 3367-8 | FETUB | FETUB |
| 3389-7 | PCI | SERPINA5 |
| 3390-72 | PIK3CA/PIK3R1 | PIK3R1\|PIK3CA |
| 3391-10 | PK3CG | PIK3CG |
| 3396-54 | Renin | REN |
| 3400-49 | TBK1 | TBK1 |
| 3412-7 | Bcl-2 | BCL2 |
| 3419-49 | CAMK2D | CAMK2D |
| 3423-59 | Chymase | CMA1 |
| 3432-21 | EPHA3 | EPHA3 |
| 3435-53 | FN1.4 | FN1 |
| 3437-80 | Flt-3 | FLT3 |
| 3447-64 | IL-8 | CXCL8 |
| 3448-13 | IR | INSR |
| 3452-17 | LCK | LCK |
| 3459-49 | PDGF Rb | PDGFRB |
| 3470-1 | sE-Selectin | SELE |
| 3474-19 | Thrombospondin-1 | THBS1 |
| 3484-60 | Angiotensinogen | AGT |
| 3486-58 | b-ECGF | FGF1 |
| 3488-64 | Catalase | CAT |
| 3489-9 | CNTF | CNTF |
| 3497-13 | IFN-aA | IFNA2 |
| 3498-53 | IL-17 | IL17A |
| 3503-4 | Integrin a1b1 | ITGB1\|ITGA1 |
| 3504-58 | LEAP-1 | HAMP |
| 3505-6 | Lymphotoxin a1/b2 | LTA\|LTB |
| 3514-49 | Proteinase-3 | PRTN3 |
| 3518-54 | TAFI | CPB2 |
| 3519-3 | TARC | CCL17 |
| 3520-58 | TGF-b3 | TGFB3 |
| 3521-16 | TSH | TSHB\|CGA |
| 3522-57 | Vasoactive Intestinal P | VIP |

| | | |
|---|---|---|
| 3534-14 | CD40 ligand, soluble | CD40LG |
| 3535-84 | DKK1 | DKK1 |
| 3538-26 | dopa decarboxylase | DDC |
| 3554-24 | Adiponectin | ADIPOQ |
| 3580-25 | a1-Antitrypsin | SERPINA1 |
| 3585-54 | BASI | BSG |
| 3593-72 | Caspase-3 | CASP3 |
| 3600-2 | Chitotriosidase-1 | CHIT1 |
| 3601-54 | CHL1 | CHL1 |
| 3605-77 | MASP3:Light | MASP1 |
| 3611-70 | Endothelin-converting | ECE1 |
| 3623-84 | LY86 | LY86 |
| 3628-3 | MP2K2 | MAP2K2 |
| 3642-4 | SLAF5 | CD84 |
| 3647-49 | TLR4:MD-2 complex | TLR4\|LY96 |
| 3651-50 | VEGF sR2 | KDR |
| 3666-17 | complement factor H-r | CFHR5 |
| 3708-62 | a2-Macroglobulin | A2M |
| 3709-4 | ALT | GPT |
| 3710-49 | Angiostatin | PLG |
| 3714-49 | CK-MB | CKM\|CKB |
| 3719-2 | p27Kip1 | CDKN1B |
| 3727-35 | PYY | PYY |
| 3728-52 | Secretin | SCT |
| 3730-81 | TNR4 | TNFRSF4 |
| 3736-60 | BMP-6 | BMP6 |
| 3739-72 | gpIIbIIIa | ITGB3\|ITGA2B |
| 3758-63 | Activated Protein C | PROC |
| 3761-4 | COX-2 | PTGS2 |
| 3773-15 | sTie-2 | TEK |
| 3796-79 | ANGL4 | ANGPTL4 |
| 3797-1 | Cadherin-2 | CDH2 |
| 3799-11 | Carbonic anhydrase III | CA3 |
| 3800-71 | CK-BB | CKB |
| 3807-1 | FGF23 | FGF23 |
| 3808-76 | FGFR-2 | FGFR2 |
| 3809-1 | FGFR-3:CD | FGFR3 |
| 3817-18 | KPCT | PRKCQ |
| 3825-18 | MK08 | MAPK8 |
| 3831-21 | pTEN | PTEN |
| 3835-11 | TLR2 | TLR2 |
| 3837-6 | ZAP70 | ZAP70 |
| 3847-56 | ETHE1 | ETHE1 |
| 3848-14 | GAPDH, liver | GAPDH |
| 3853-56 | MDHC | MDH1 |
| 3854-24 | NACA | NACA |
| 3855-56 | Peroxiredoxin-1 | PRDX1 |

| | | |
|---|---|---|
| 3860-7 | PSA6 | PSMA6 |
| 3881-49 | DLC8 | DYNLL1 |
| 3889-64 | Lamin-B1 | LMNB1 |
| 3890-8 | LDH-H 1 | LDHB |
| 4125-52 | sRAGE | AGER |
| 4127-75 | C6 | C6 |
| 4128-27 | Eotaxin-2 | CCL24 |
| 4130-71 | FGF-6 | FGF6 |
| 4131-72 | Fibronectin | FN1 |
| 4132-27 | FST | FST |
| 4138-25 | IL-20 | IL20 |
| 4139-71 | IL-6 sRa | IL6R |
| 4141-79 | IP-10 | CXCL10 |
| 4148-49 | PAPP-A | PAPPA |
| 4149-8 | PDGF-BB | PDGFB |
| 4150-75 | Plasmin | PLG |
| 4151-6 | Plasminogen | PLG |
| 4152-58 | Prekallikrein | KLKB1 |
| 4153-11 | alpha-1-antichymotryp | KLK3\|SERPINA3 |
| 4154-57 | P-Selectin | SELP |
| 4156-74 | TGF-b2 | TGFB2 |
| 4157-2 | Thrombin | F2 |
| 4158-54 | uPA | PLAU |
| 4160-49 | MMP-2 | MMP2 |
| 4162-54 | Transferrin | TF |
| 4232-19 | IGF-I sR | IGF1R |
| 4234-8 | IL-1 R4 | IL1RL1 |
| 4240-31 | M2-PK | PKM |
| 4245-80 | MDM2 | MDM2 |
| 4261-55 | paraoxonase 1 | PON1 |
| 4276-10 | prostatic binding prote | PEBP1 |
| 4280-47 | PSA2 | PSMA2 |
| 4306-4 | Transketolase | TKT |
| 4309-59 | Triosephosphate isom | TPI1 |
| 4318-12 | PTP-1C | PTPN6 |
| 4322-28 | AMNLS | AMN |
| 4328-2 | BOC | BOC |
| 4337-49 | CRP | CRP |
| 4342-10 | sICAM-1 | ICAM1 |
| 4374-45 | MIC-1 | GDF15 |
| 4413-3 | SLPI | SLPI |
| 4414-69 | SP-D | SFTPD |
| 4423-77 | BCL2-like 1 protein | BCL2L1 |
| 4430-44 | Collectin Kidney 1 | COLEC11 |
| 4455-89 | MFGM | MFGE8 |
| 4460-8 | PDPK1 | PDPK1 |
| 4467-49 | SPARCL1 | SPARCL1 |

| | | |
|---|---|---|
| 4468-21 | SPHK2 | SPHK2 |
| 4479-14 | C1-Esterase Inhibitor | SERPING1 |
| 4481-34 | C4 | C4B\|C4A |
| 4482-66 | C5b, 6 Complex | C6\|C5 |
| 4493-92 | IL-11 | IL11 |
| 4496-60 | MMP-12 | MMP12 |
| 4498-62 | NCAM-120 | NCAM1 |
| 4499-21 | PDGF-AA | PDGFA |
| 4500-50 | SCGF-alpha | CLEC11A |
| 4541-49 | CDON | CDON |
| 4542-24 | Clusterin | CLU |
| 4545-53 | DnaJ homolog | DNAJC19 |
| 4546-27 | EMR2 | ADGRE2 |
| 4559-64 | KYNU | KYNU |
| 4673-13 | IL-6 | IL6 |
| 4697-59 | GM-CSF | CSF2 |
| 4703-87 | TNF-b | LTA |
| 4719-58 | Protein disulfide isome | PDIA3 |
| 4721-54 | TFF3 | TFF3 |
| 4775-34 | Gelsolin | GSN |
| 4807-13 | CO8A1 | COL8A1 |
| 4831-4 | sL-Selectin | SELL |
| 4832-75 | TRAIL R1 | TNFRSF10A |
| 4834-61 | Epithelial cell kinase | EPHA2 |
| 4842-62 | Glypican 3 | GPC3 |
| 4851-25 | IL-1a | IL1A |
| 4859-6 | BMPR1A | BMPR1A |
| 4862-63 | BMP RII | BMPR2 |
| 4866-59 | TrkB | NTRK2 |
| 4867-15 | VEGF121 | VEGFA |
| 4874-3 | Angiogenin | ANG |
| 4876-32 | Coagulation Factor IX | F9 |
| 4880-21 | GDF2 | GDF2 |
| 4883-56 | Insulin | INS |
| 4889-82 | WNT7A | WNT7A |
| 4890-10 | ACTH | POMC |
| 4891-50 | Glucagon | GCG |
| 4903-72 | Calcineurin | PPP3R1\|PPP3CA |
| 4906-35 | Coagulation Factor V | F5 |
| 4908-6 | Endoglin | ENG |
| 4914-10 | HCG | CGB7\|CGB3\|CGA |
| 4915-64 | Hemoglobin | HBB\|HBA1 |
| 4917-62 | Integrin aVb5 | ITGB5\|ITGAV |
| 4920-10 | Lysozyme | LYZ |
| 4924-32 | MMP-1 | MMP1 |
| 4929-55 | SHBG | SHBG |
| 4931-59 | TF | F3 |

| | | |
|---|---|---|
| 4956-2 | EPI | EREG |
| 4960-72 | annexin I | ANXA1 |
| 4964-67 | ARTS1 | ERAP1 |
| 4969-2 | Carbonic anhydrase I | CA1 |
| 4970-55 | carbonic anhydrase II | CA2 |
| 4971-1 | CATZ | CTSZ |
| 4973-18 | cIAP-2 | BIRC3 |
| 4976-57 | CRK | CRK |
| 4979-34 | DERM | DPT |
| 4982-54 | Elafin | PI3 |
| 4985-11 | FABPE | FABP5 |
| 4986-59 | FAK1 | PTK2 |
| 4990-87 | GP1BA | GP1BA |
| 4992-49 | GRN | GRN |
| 5002-76 | MMP-14 | MMP14 |
| 5004-69 | MK11 | MAPK11 |
| 5007-1 | MAPK14 | MAPK14 |
| 5008-51 | Mn SOD | SOD2 |
| 5009-11 | Moesin | MSN |
| 5011-11 | PBEF | NAMPT |
| 5012-67 | Myokinase, human | AK1 |
| 5015-15 | PAFAH | PLA2G7 |
| 5018-68 | Peroxiredoxin-6 | PRDX6 |
| 5020-50 | phosphoglycerate kina | PGK1 |
| 5023-23 | PUR8 | ADSL |
| 5024-67 | Rb | RB1 |
| 5028-59 | sCD163 | CD163 |
| 5033-27 | Tropomyosin 1 alpha c | TPM1 |
| 5034-79 | Trypsin 2 | PRSS2 |
| 5036-50 | TSG-6 | TNFAIP6 |
| 5061-27 | B7-H2 | ICOSLG |
| 5080-131 | GPNMB:ECD | GPNMB |
| 5088-175 | IL-23 R | IL23R |
| 5089-11 | IL-7 Ra | IL7R |
| 5090-49 | ILT-2 | LILRB1 |
| 5092-51 | JAG1:ECD | JAG1 |
| 5100-53 | LIMP II | SCARB2 |
| 5107-7 | Notch 1 | NOTCH1 |
| 5108-72 | Notch-3 | NOTCH3 |
| 5111-15 | NRX3B | NRXN3 |
| 5117-14 | ROBO3 | ROBO3 |
| 5133-17 | TGF-b R II | TGFBR2 |
| 5138-50 | TWEAKR | TNFRSF12A |
| 5183-53 | AMPK a1b1g1 | PRKAG1\|PRKAB1\|PRKAA1 |
| 5223-59 | GCKR | GCKR |
| 5228-25 | KIF23 | KIF23 |
| 5230-99 | HMGR | HMGCR |

| | | |
|---|---|---|
| 5245-40 | AMPK a2b2g1 | PRKAG1\|PRKAB2\|PRKAA2 |
| 5254-69 | PDE3A | PDE3A |
| 5259-2 | TAK1-TAB1 | TAB1\|MAP3K7 |
| 5260-80 | TYK2 | TYK2 |
| 5264-65 | calreticulin | CALR |
| 5272-55 | SHC1:SH2 | SHC1 |
| 5301-7 | Eotaxin | CCL11 |
| 5312-49 | Apo E2 | APOE |
| 5315-22 | Troponin T | TNNT2 |
| 5316-54 | Prothrombin | F2 |
| 5350-14 | GPC6 | GPC6 |
| 5351-52 | hnRNP A2/B1 | HNRNPA2B1 |
| 5352-11 | HVEM | TNFRSF14 |
| 5353-89 | IL-1Ra | IL1RN |
| 5360-9 | PKB beta | AKT2 |
| 5400-52 | sLeptin R | LEPR |
| 5412-53 | CD27 | CD27 |
| 5424-55 | RANK | TNFRSF11A |
| 5430-66 | SHPS1 | SIRPA |
| 5441-67 | Troponin I | TNNI3 |
| 5443-62 | ANP | NPPA |
| 5451-1 | ALCAM | ALCAM |
| 5468-67 | IL-17 RC | IL17RC |
| 5475-10 | PKC-B-II | PRKCB |
| 5480-49 | RANTES | CCL5 |
| 5481-16 | RASA1 | RASA1 |
| 5508-62 | Cathepsin D | CTSD |
| 5509-7 | EGF:ECD | EGF |
| 5532-53 | bFGF-R | FGFR1 |
| 5534-49 | TRAIL R2:ECD | TNFRSF10B |
| 5542-22 | NRP1 | NRP1 |
| 5584-21 | Holo-TC II | TCN2 |
| 5620-13 | AMD | PAM |
| 5621-64 | THSD1 | THSD1 |
| 5626-20 | CTRC | CTRC |
| 5627-53 | GON1 | GNRH1 |
| 5635-66 | TREM2 | TREM2 |
| 5658-64 | coagulation factor XIII | F13B |
| 5659-11 | TRH | TRH |
| 5660-51 | SOD3 | SOD3 |
| 5661-15 | IL-18 | IL18 |
| 5666-64 | IL-1 sRII | IL1R2 |
| 5675-6 | Tenascin | TNC |
| 5682-13 | VASN | VASN |
| 5692-79 | TNF-a | TNF |
| 5698-60 | Tenascin-X | TNXB |
| 5707-55 | ELK3 | ELK3 |

| | | |
|---|---|---|
| 5743-82 | CART | CARTPT |
| 5746-37 | EFNB1 | EFNB1 |
| 5748-20 | Acid ceramidase | ASAH1 |
| 5757-45 | GNAS3 | GNAS |
| 5792-8 | AFP | AFP |
| 5801-72 | b-NGF | NGF |
| 5807-77 | CD70 | CD70 |
| 5810-25 | Cripto | TDGF1 |
| 5813-58 | Epo | EPO |
| 5825-49 | IFN-g R1 | IFNGR1 |
| 5854-60 | tau | MAPT |
| 5864-10 | aldolase A | ALDOA |
| 5867-60 | ARGI1 | ARG1 |
| 5870-23 | BAD | BAD |
| 5903-91 | HSP70 protein 8 | HSPA8 |
| 5909-51 | Nucleoside diphosphat | NME1 |
| 5915-58 | PEX5 | PEX5 |
| 5934-1 | Ferritin | FTL\|FTH1 |
| 5939-42 | TWEAK | TNFSF12 |
| 5954-62 | PTH | PTH |
| 5957-30 | Somatostatin-28 | SST |
| 5980-55 | BOLA3 | BOLA3 |
| 6003-28 | NEUR1 | NEU1 |
| 6020-52 | Urotensin-II | UTS2 |
| 6024-68 | CBPE | CPE |
| 6027-31 | REL3 | RLN3 |
| 6035-2 | SIAT1 | ST6GAL1 |
| 6049-64 | PTPRS | PTPRS |
| 6077-63 | CECR1 | ADA2 |
| 6123-69 | p53 | TP53 |
| 6207-10 | prosaposin | PSAP |
| 6232-54 | B7-2 | CD86 |
| 6245-4 | Nectin-2 | NECTIN2 |
| 6350-43 | Apo C-II | APOC2 |
| 6373-54 | DLK1 | DLK1 |
| 6375-75 | XXLT1 | XXYLT1 |
| 6379-62 | ATL2 | ADAMTSL2 |
| 6382-17 | MANBA | MANBA |
| 6383-90 | TLL1 | TLL1 |
| 6398-12 | LONM | LONP1 |
| 6420-4 | SDHF2 | SDHAF2 |
| 6433-57 | FA20A | FAM20A |
| 6440-31 | MFAP5 | MFAP5 |
| 6447-73 | PTX3 | PTX3 |
| 6448-36 | Sema E | SEMA3C |
| 6461-54 | Apo C-III | APOC3 |
| 6462-12 | TIMP-4 | TIMP4 |

| | | |
|---|---|---|
| 6495-14 | Endothelin 1 | EDN1 |
| 6496-60 | DLK1:ECD | DLK1 |
| 6504-65 | Lysyl oxidase-like prot | LOXL2 |
| 6518-85 | ghrelin | GHRL |
| 6520-87 | MGP | MGP |
| 6544-33 | NELL1 | NELL1 |
| 6545-58 | PRIO | PRNP |
| 6558-5 | COL10 | COLEC10 |
| 6563-78 | HSP 70 | HSPA1A |
| 6577-64 | LAMA4 | LAMA4 |
| 6584-1 | SRCH | HRC |
| 6593-5 | GALT3 | GALNT3 |
| 6599-5 | APCD1 | APCDD1 |
| 6605-17 | IGFALS | IGFALS |
| 6622-90 | APEL | APLN |
| 6649-51 | NET1 | NTN1 |
| 6895-1 | TR:CD | TFRC |
| 6909-40 | MGAT2 | MGAT2 |
| 6932-42 | ITA5 | ITGA5 |
| 6935-123 | BAMBI:CD | BAMBI |
| 6941-11 | SUMF1 | SUMF1 |
| 6948-82 | FIG4 | FIG4 |
| 6963-82 | SELPL:CD | SELPLG |
| 6973-111 | IGF-II:Pro-form | IGF2 |
| 7015-8 | LIRB5 | LILRB5 |
| 7045-4 | BNIP3 | BNIP3 |
| 7050-5 | NEGR1 | NEGR1 |
| 7100-31 | CD2 | CD2 |
| 7124-18 | IL-21 | IL21 |
| 7127-3 | Apo A-II | APOA2 |
| 7131-8 | CETP | CETP |
| 7140-1 | ELA2A | CELA2A |
| 7142-5 | CBPN | CPN1 |
| 7161-25 | G6PE | H6PD |
| 7174-15 | CD38 | CD38 |
| 7178-59 | DEPP | DEPP1 |
| 7181-17 | VAPB | VAPB |
| 7182-1 | carboxylesterase, liver | CES1 |
| 7186-111 | STX3 | STX3 |
| 7187-3 | PPM1L | PPM1L |
| 7191-32 | SLMAP | SLMAP |
| 7194-36 | NPTN | NPTN |
| 7201-5 | ISCU | ISCU |
| 7206-20 | F16P1 | FBP1 |
| 7247-1 | GORAB | GORAB |
| 7262-191 | CHSTE | CHST14 |
| 7624-19 | ANK2 | ANK2 |

| | | |
|---|---|---|
| 7628-40 | CREL1 | CRELD1 |
| 7655-11 | N-terminal pro-BNP | NPPB |
| 7660-21 | Tropomyosin 4 | TPM4 |
| 7735-17 | PEDF | SERPINF1 |
| 7747-47 | NDUBB | NDUFB11 |
| 7748-11 | NDUV2 | NDUFV2 |
| 7754-11 | JAG1:CD | JAG1 |
| 7761-125 | CHKB | CHKB |
| 7770-25 | NFU1 | NFU1 |
| 7784-1 | Kininogen, HMW | KNG1 |
| 7788-1 | CF6 | ATP5PF |
| 7789-182 | PRDX4 | PRDX4 |
| 7790-21 | LIF | LIF |
| 7806-33 | B4GT7 | B4GALT7 |
| 7849-3 | Glutaminyl cyclase | QPCT |
| 7850-1 | COX42 | COX4I2 |
| 7853-19 | SCO1 | SCO1 |
| 7857-22 | NEUT | NTS |
| 7870-8 | CNDP1 | CNDP1 |
| 7875-86 | PLEK | PLEK |
| 7887-57 | COX5B | COX5B |
| 7888-58 | CCD56 | COA3 |
| 7893-19 | OLR1 | OLR1 |
| 7895-108 | MRAP | MRAP |
| 7905-30 | HPT | HP |
| 7911-29 | SCN4B | SCN4B |
| 7922-5 | Adrenomedullin | ADM |
| 7924-7 | CD3G | CD3G |
| 7953-20 | SLAF1 | SLAMF1 |
| 7981-230 | B3GT6 | B3GALT6 |
| 8005-1 | MXRA7 | MXRA7 |
| 8009-121 | SURF1 | SURF1 |
| 8017-23 | FXRD1 | FOXRED1 |
| 8036-75 | FSHB | FSHB |
| 8038-41 | NSDHL | NSDHL |
| 8043-153 | COMP | COMP |
| 8051-10 | AGGF1 | AGGF1 |
| 8055-33 | IDD | DGCR2 |
| 8059-1 | Tpo | THPO |
| 8067-21 | KCNE3 | KCNE3 |
| 8068-43 | K1161:ECD | MYORG |
| 8070-88 | DLG4 | DLG4 |
| 8074-32 | TMM70 | TMEM70 |
| 8086-49 | ITM2B | ITM2B |
| 8089-173 | NR4A1 | NR4A1 |
| 8091-16 | MASP3:Heavy | MASP1 |
| 8093-13 | K1161:CD | MYORG |

| | | |
|---|---|---|
| 8094-20 | CISD2 | CISD2 |
| 8099-42 | SPON2 | SPON2 |
| 8100-15 | ADM2 | ADM2 |
| 8231-122 | VEGF sR1 | FLT1 |
| 8243-55 | TATI | SPINK1 |
| 8246-9 | PCSK9 | PCSK9 |
| 8250-2 | PTPRJ | PTPRJ |
| 8265-225 | LAP2B | TMPO |
| 8273-84 | IL31R | IL31RA |
| 8285-64 | PACA | ADCYAP1 |
| 8304-50 | OPG | TNFRSF11B |
| 8309-12 | HYAL1 | HYAL1 |
| 8314-71 | GNS | GNS |
| 8353-15 | SCN2B | SCN2B |
| 8356-88 | NEU1 | OXT |
| 8358-30 | Peroxiredoxin-3 | PRDX3 |
| 8364-74 | UST | UST |
| 8368-102 | TNF sR-II | TNFRSF1B |
| 8376-25 | LSHB | LHB |
| 8380-244 | DLK1:CD | DLK1 |
| 8403-18 | Fatty acid synthase | FASN |
| 8406-17 | IGF-I | IGF1 |
| 8429-16 | AXIN2 | AXIN2 |
| 8446-4 | PACAP-27 | ADCYAP1 |
| 8458-16 | a-Synuclein | SNCA |
| 8462-18 | HGH | GH1 |
| 8470-213 | RNase H1 | RNASEH1 |
| 8480-29 | FBLN3 | EFEMP1 |
| 8484-24 | Leptin | LEP |
| 8529-1 | TRAIL R2:Death | TNFRSF10B |
| 8587-21 | ISK52 | SPINK14 |
| 8644-46 | Cathepsin H | CTSH |
| 8645-257 | PEX14:C-term | PEX14 |
| 8687-26 | T106B | TMEM106B |
| 8690-25 | CAV3 | CAV3 |
| 8750-46 | Vinculin | VCL |
| 8756-41 | KCE1L:N-term | KCNE5 |
| 8767-44 | Endothelin-converting | ECE1 |
| 8772-5 | EFNB2:CD | EFNB2 |
| 8790-6 | Arylsulfatase A | ARSA |
| 8795-48 | TR:ECD | TFRC |
| 8810-26 | HEPACAM | HEPACAM |
| 8811-24 | BAMBI:ECD | BAMBI |
| 8834-58 | Calnexin | CANX |
| 8854-59 | RIR2B | RRM2B |
| 8863-3 | P85A | PIK3R1 |
| 8874-53 | CLN5:LD | CLN5 |

| | | |
|---|---|---|
| 8893-29 | PARP:region 1 | PARP1 |
| 8908-14 | KCE1L:CD | KCNE5 |
| 8916-32 | STIM1:CD | STIM1 |
| 8924-55 | CALY | CALY |
| 8932-1 | ENTP6 | ENTPD6 |
| 8935-22 | MRP6 | ABCC6 |
| 8946-38 | NR1H4 | NR1H4 |
| 8952-65 | G-CSF | CSF3 |
| 8955-60 | GL8D1 | GLT8D1 |
| 8985-13 | SRAC1 | SERAC1 |
| 8986-2 | SERA | PHGDH |
| 9000-177 | SLC31 | SLC3A1 |
| 9017-58 | LPH | LCT |
| 9021-1 | TIM-1 | HAVCR1 |
| 9076-25 | PENK | PENK |
| 9125-23 | MASP3:Sushi 1 and Su | MASP1 |
| 9171-11 | CSRP3 | CSRP3 |
| 9173-21 | PGM1 | PGM1 |
| 9176-3 | MUC1:region 2 | MUC1 |
| 9178-30 | NEUREGULIN-1 | NRG1 |
| 9185-15 | TFF1 | TFF1 |
| 9188-119 | MIG | CXCL9 |
| 9197-4 | LEG9 | LGALS9 |
| 9204-33 | Corticotropin-lipotropi | POMC |
| 9213-24 | FTCD | FTCD |
| 9237-54 | Lysosomal acid phosph | ACP2 |
| 9244-27 | PPT1 | PPT1 |
| 9253-52 | BGAT | ABO |
| 9257-14 | TACO1 | TACO1 |
| 9259-35 | DR6 | TNFRSF21 |
| 9269-7 | Biotinidase | BTD |
| 9271-101 | STIM1:ECD | STIM1 |
| 9283-8 | CD44 | CD44 |
| 9287-6 | SPRE | SPR |
| 9337-43 | TKN1 | TAC1 |
| 9340-17 | FKB14 | FKBP14 |
| 9341-1 | PDGFD | PDGFD |
| 9343-16 | IL-2 sRb | IL2RB |
| 9357-4 | CREG1 | CREG1 |
| 9366-54 | IL-21 sR | IL21R |
| 9377-25 | SCF | KITLG |
| 9385-4 | GAA | GAA |
| 9388-18 | MCEE | MCEE |
| 9412-52 | Activin RIIB | ACVR2B |
| 9436-2 | ID-1 | ID1 |
| 9443-137 | cathepsin K | CTSK |
| 9444-70 | DGCR6 | DGCR6 |

| | | |
|---|---|---|
| 9451-20 | Uromodulin | UMOD |
| 9457-3 | CAV2 | CAV2 |
| 9459-7 | Fas, soluble | FAS |
| 9484-75 | Desmoglein-2 | DSG2 |
| 9504-19 | RB27A | RAB27A |
| 9522-3 | AIF | AIFM1 |
| 9543-131 | COG8 | COG8 |
| 9576-58 | MNX1 | MNX1 |
| 9739-4 | MSH2 | MSH2 |
| 9750-7 | S100A4 | S100A4 |
| 9756-6 | TAGL | TAGLN |
| 9759-13 | INDO | IDO1 |
| 9786-310 | IEX1 | IER3 |
| 9789-52 | Nesprin-2 | SYNE2 |
| 9796-4 | CEL | CEL |
| 9800-20 | NDUB8 | NDUFB8 |
| 9802-27 | PLPL2 | PNPLA2 |
| 9823-2 | DYR | DHFR |
| 9830-109 | VAV3 | VAV3 |
| 9832-33 | HGD | HGD |
| 9834-62 | ADH1B | ADH1B |
| 9837-60 | NAD(P)H dehydrogena | NQO1 |
| 9838-4 | SMAD1 | SMAD1 |
| 9842-2 | b-Catenin | CTNNB1 |
| 9844-138 | ACTN2 | ACTN2 |
| 9845-33 | PARK7 | PARK7 |
| 9846-32 | Rho-GDI beta | ARHGDIB |
| 9849-13 | APEX1 | APEX1 |
| 9859-180 | SSAT-1 | SAT1 |
| 9867-23 | F16P2 | FBP2 |
| 9869-28 | NF-kappa-B p105 | NFKB1 |
| 9873-17 | ERRalpha | ESRRA |
| 9874-28 | p15-INK4b | CDKN2B |
| 9877-28 | CRKL | CRKL |
| 9878-3 | SULT 1E | SULT1E1 |
| 9880-33 | T23O | TDO2 |
| 9883-29 | Glyoxalase I | GLO1 |
| 9886-28 | XRCC4 | XRCC4 |
| 9895-77 | XPF | ERCC4 |
| 9897-9 | MCM6 | MCM6 |
| 9900-36 | NFH | NEFH |
| 9923-6 | SOS1 | SOS1 |
| 9937-7 | CXA1 | GJA1 |
| 9952-57 | TFAM | TFAM |
