## Supplemental Table 2 for "The Evolution of Primary Aldosteronism and the Role of Norrin"

| GeneID | logFC | AveExpr | t | P.Value | adj.P.Val | B | Significance |
|---|---|---|---|---|---|---|---|
| CYBRD1 | 1.46716 | 2.868703018 | 80.36145 | 9.0442E-101 | 1.35662E-97 | 218.6556901 | Higher in PA |
| CCL20 | -1.21918 | 2.851902307 | -62.8532 | 4.30423E-89 | 3.22817E-86 | 192.7163528 | Higher in NT |
| F11 | 0.77433 | 3.159581099 | 57.41089 | 7.97741E-85 | 3.98871E-82 | 183.1170865 | Higher in PA |
| ITGB1\|ITGA2 | -1.07484 | 3.108109991 | -34.7432 | 9.29734E-62 | 3.4865E-59 | 130.5685646 | Higher in NT |
| PTPN6 | -1.23145 | 3.335371748 | -27.0275 | 7.41664E-51 | 2.22499E-48 | 105.5016861 | Higher in NT |
| ECHS1 | -1.19612 | 3.942212338 | -20.8233 | 2.7719E-40 | 6.92975E-38 | 81.12861025 | Higher in NT |
| NAMPT | -0.34937 | 3.191042609 | -20.6631 | 5.51092E-40 | 1.18091E-37 | 80.44010943 | Higher in NT |
| ATP5PF | -1.15862 | 3.50071229 | -20.441 | 1.43607E-39 | 2.69262E-37 | 79.48050887 | Higher in NT |
| IVD | -1.16628 | 4.004069238 | -19.6822 | 3.96675E-38 | 6.61124E-36 | 76.15531382 | Higher in NT |
| PSPH | -0.42235 | 3.53229496 | -19.5012 | 8.8466E-38 | 1.32699E-35 | 75.35160146 | Higher in NT |
| MPI | -0.63404 | 3.437882484 | -18.6754 | 3.62156E-36 | 4.93849E-34 | 71.63191392 | Higher in NT |
| DECR1 | -1.17780 | 4.054489921 | -18.5454 | 6.54944E-36 | 8.1868E-34 | 71.03820519 | Higher in NT |
| ACADM | -1.22313 | 3.15183598 | -18.4869 | 8.55213E-36 | 9.86784E-34 | 70.77084968 | Higher in NT |
| SYNE2 | -0.58593 | 3.420986601 | -18.2087 | 3.06562E-35 | 3.28459E-33 | 69.4915352 | Higher in NT |
| HSD17B4 | -0.82103 | 3.767128777 | -17.3044 | 2.08061E-33 | 2.08061E-31 | 65.26520868 | Higher in NT |
| MYL9 | -0.95544 | 4.143596876 | -17.253 | 2.65252E-33 | 2.48674E-31 | 65.02185937 | Higher in NT |
| HMGCL | -1.02050 | 3.320190886 | -16.6839 | 3.98973E-32 | 3.52035E-30 | 62.30558006 | Higher in NT |
| SERPINH1 | -0.80141 | 3.128388277 | -16.6716 | 4.23402E-32 | 3.52835E-30 | 62.24603403 | Higher in NT |
| CHKB | -0.81203 | 3.439769439 | -16.4293 | 1.35949E-31 | 1.07328E-29 | 61.07719123 | Higher in NT |
| ITGB3\|ITGA2B | -0.96880 | 4.481427883 | -16.3581 | 1.91817E-31 | 1.43863E-29 | 60.73225394 | Higher in NT |
| STIM1 | -0.68008 | 3.98576837 | -15.3722 | 2.40284E-29 | 1.71632E-27 | 55.89285864 | Higher in NT |
| GRHPR | -0.42912 | 3.450928554 | -15.2297 | 4.87707E-29 | 3.32528E-27 | 55.18374667 | Higher in NT |
| COX5A | -0.96278 | 3.671721407 | -15.1319 | 7.93728E-29 | 5.17649E-27 | 54.69589892 | Higher in NT |
| ITGA3\|ITGB1 | 0.47654 | 2.581908101 | 15.10992 | 8.8591E-29 | 5.53694E-27 | 54.58584068 | Higher in PA |
| AKT1S1 | -0.62261 | 3.428849971 | -15.0098 | 1.4605E-28 | 8.76298E-27 | 54.08509753 | Higher in NT |
| BCAT2 | -0.80235 | 3.354960308 | -14.9946 | 1.5759E-28 | 9.09172E-27 | 54.00892463 | Higher in NT |
| BOLA3 | -0.77615 | 3.290574782 | -14.9113 | 2.39226E-28 | 1.32903E-26 | 53.59083573 | Higher in NT |
| TFAM | -0.98889 | 3.99081806 | -14.8702 | 2.93977E-28 | 1.57488E-26 | 53.38441244 | Higher in NT |
| DBT | -0.83726 | 3.187514242 | -14.7552 | 5.23917E-28 | 2.70992E-26 | 52.80566945 | Higher in NT |
| OXCT1 | -0.70102 | 4.185605302 | -14.6517 | 8.82112E-28 | 4.41056E-26 | 52.28388284 | Higher in NT |
| AMT | -0.85320 | 3.664306837 | -14.6403 | 9.34369E-28 | 4.52114E-26 | 52.22624352 | Higher in NT |
| HSPB1 | -0.76350 | 3.506560404 | -14.4884 | 2.01231E-27 | 9.43271E-26 | 51.45793363 | Higher in NT |
| GOT2 | -0.72198 | 3.635382429 | -14.4693 | 2.2165E-27 | 1.0075E-25 | 51.36114694 | Higher in NT |

| | | | | | | | |
|---|---|---|---|---|---|---|---|
| HIBCH | -0.89779 | 3.908918345 | -14.4318 | 2.68025E-27 | 1.18246E-25 | 51.17089246 | Higher in NT |
| GFPT1 | -0.41754 | 3.569576298 | -14.4116 | 2.96853E-27 | 1.27223E-25 | 51.06858936 | Higher in NT |
| HSPE1 | -0.83785 | 3.962647173 | -14.3835 | 3.42321E-27 | 1.42634E-25 | 50.92587469 | Higher in NT |
| HADH | -0.96714 | 4.178256435 | -14.3651 | 3.75874E-27 | 1.52381E-25 | 50.83223705 | Higher in NT |
| JAK2 | -0.62621 | 3.809342461 | -14.3077 | 5.03064E-27 | 1.98578E-25 | 50.54036498 | Higher in NT |
| CSRP3 | -0.39507 | 2.941152616 | -14.2994 | 5.24508E-27 | 2.01734E-25 | 50.4985637 | Higher in NT |
| PYGB | -0.75876 | 4.377778928 | -14.269 | 6.12312E-27 | 2.29617E-25 | 50.34356883 | Higher in NT |
| UFD1 | -0.82076 | 3.814792594 | -14.259 | 6.44271E-27 | 2.35709E-25 | 50.29262155 | Higher in NT |
| DARS2 | -1.14089 | 4.079040949 | -14.2498 | 6.75208E-27 | 2.40116E-25 | 50.24565678 | Higher in NT |
| NDUFAF2 | -0.89055 | 3.428754032 | -14.246 | 6.88332E-27 | 2.40116E-25 | 50.22637955 | Higher in NT |
| NDUFV2 | -0.46783 | 3.396655509 | -14.1475 | 1.1361E-26 | 3.87306E-25 | 49.72462425 | Higher in NT |
| PRKAR2B | -0.62212 | 3.460196949 | -14.1002 | 1.44657E-26 | 4.82189E-25 | 49.48271263 | Higher in NT |
| RAB21 | -0.66338 | 4.639719496 | -14.0945 | 1.48911E-26 | 4.8558E-25 | 49.45368775 | Higher in NT |
| NME4 | -0.77544 | 3.759522268 | -14.0262 | 2.1107E-26 | 6.73626E-25 | 49.10440279 | Higher in NT |
| CIRBP | -0.64169 | 4.235322609 | -13.9849 | 2.60664E-26 | 8.14576E-25 | 48.89309214 | Higher in NT |
| UBQLN2 | -0.49244 | 3.754954545 | -13.9736 | 2.76152E-26 | 8.45365E-25 | 48.83530014 | Higher in NT |
| AKT2 | -0.67716 | 3.898620934 | -13.833 | 5.67294E-26 | 1.70188E-24 | 48.11450375 | Higher in NT |
| MESD | -0.55726 | 3.906528502 | -13.7456 | 8.88601E-26 | 2.60627E-24 | 47.66521397 | Higher in NT |
| LTA4H | -0.36616 | 3.485300519 | -13.7423 | 9.03507E-26 | 2.60627E-24 | 47.64855957 | Higher in NT |
| PRDX3 | -0.75372 | 3.913809945 | -13.7076 | 1.08028E-25 | 3.0574E-24 | 47.46966612 | Higher in NT |
| FH | -0.75503 | 3.201270536 | -13.6505 | 1.44919E-25 | 4.02552E-24 | 47.1755612 | Higher in NT |
| HSPD1 | -0.84141 | 4.214126209 | -13.6197 | 1.69787E-25 | 4.63056E-24 | 47.01701558 | Higher in NT |
| LZTFL1 | -0.50509 | 3.319092385 | -13.5116 | 2.96471E-25 | 7.94119E-24 | 46.45903195 | Higher in NT |
| UBQLN4 | -0.51897 | 3.627556326 | -13.4764 | 3.5554E-25 | 9.35633E-24 | 46.27715964 | Higher in NT |
| ENO1 | -0.77629 | 4.770243145 | -13.4109 | 4.98587E-25 | 1.28945E-23 | 45.93869003 | Higher in NT |
| ALDH6A1 | -0.79026 | 3.673933295 | -13.4004 | 5.26478E-25 | 1.3385E-23 | 45.88420674 | Higher in NT |
| CTSC | -0.29797 | 3.952967605 | -13.331 | 7.53937E-25 | 1.88484E-23 | 45.52477299 | Higher in NT |
| COPB2 | -0.76437 | 4.24397099 | -13.2932 | 9.16849E-25 | 2.25455E-23 | 45.32896174 | Higher in NT |
| CS | -0.82727 | 4.641392842 | -13.2404 | 1.20563E-24 | 2.91685E-23 | 45.05490693 | Higher in NT |
| GSK3B | -0.81512 | 3.544163378 | -13.1765 | 1.68009E-24 | 4.0002E-23 | 44.72278585 | Higher in NT |
| HTRA2 | -0.42815 | 3.485271783 | -13.1422 | 2.00741E-24 | 4.70487E-23 | 44.54463956 | Higher in NT |
| DUSP6 | -0.15557 | 3.198628226 | -13.1232 | 2.2159E-24 | 5.1136E-23 | 44.44575145 | Higher in NT |
| ACADVL | -0.96498 | 3.866754392 | -13.1162 | 2.29791E-24 | 5.22251E-23 | 44.40938108 | Higher in NT |
| MAPK1 | -0.54072 | 3.266217791 | -13.0878 | 2.66333E-24 | 5.96269E-23 | 44.26168637 | Higher in NT |

| | | | | | | | |
|---|---|---|---|---|---|---|---|
| MCEE | -0.60825 | 3.554587185 | -13.0706 | 2.91186E-24 | 6.42322E-23 | 44.17240441 | Higher in NT |
| ACAT1 | -0.75813 | 3.658369872 | -13.027 | 3.65295E-24 | 7.82941E-23 | 43.94549294 | Higher in NT |
| PDIA3 | -0.69293 | 3.744144757 | -13.027 | 3.65373E-24 | 7.82941E-23 | 43.94528103 | Higher in NT |
| GHRL | -0.78524 | 3.407799423 | -13.0229 | 3.73216E-24 | 7.88485E-23 | 43.92402537 | Higher in NT |
| MMAB | -0.62737 | 3.612985812 | -12.9642 | 5.06688E-24 | 1.0556E-22 | 43.61807217 | Higher in NT |
| DDOST | -0.43820 | 3.292852516 | -12.7858 | 1.28529E-23 | 2.64101E-22 | 42.68664111 | Higher in NT |
| PTP4A2 | -0.41437 | 3.075042852 | -12.6072 | 3.27178E-23 | 6.63198E-22 | 41.75181986 | Higher in NT |
| FKBP14 | -0.42966 | 3.12238239 | -12.5939 | 3.50917E-23 | 7.01834E-22 | 41.68174344 | Higher in NT |
| PRKAR1A | -0.49950 | 4.166680407 | -12.5557 | 4.28665E-23 | 8.46049E-22 | 41.48153376 | Higher in NT |
| AKT1 | -0.56241 | 3.262175841 | -12.3414 | 1.32069E-22 | 2.57278E-21 | 40.35593395 | Higher in NT |
| ADSL | -0.48671 | 3.639394498 | -12.3121 | 1.54084E-22 | 2.96314E-21 | 40.20173215 | Higher in NT |
| PDHX | -0.67176 | 3.395429424 | -12.2249 | 2.43827E-22 | 4.62962E-21 | 39.74268732 | Higher in NT |
| ACADSB | -0.51105 | 3.150230856 | -12.1312 | 3.99506E-22 | 7.49074E-21 | 39.24886437 | Higher in NT |
| PRKCA | -0.74756 | 4.247539277 | -12.1277 | 4.06887E-22 | 7.53495E-21 | 39.23055591 | Higher in NT |
| PPP3R1\|PPP3CA | -0.56049 | 4.294363959 | -12.1192 | 4.25486E-22 | 7.78327E-21 | 39.18585784 | Higher in NT |
| CD69 | -0.56158 | 2.431776534 | -12.0753 | 5.36591E-22 | 9.69278E-21 | 38.95384512 | Higher in NT |
| VPS26A | -0.44217 | 4.315786419 | -12.0731 | 5.42796E-22 | 9.69278E-21 | 38.94234737 | Higher in NT |
| UROD | -0.40374 | 3.606034533 | -12.0507 | 6.10776E-22 | 1.07784E-20 | 38.82434901 | Higher in NT |
| SMAD3 | -0.45518 | 3.778484394 | -12.0383 | 6.52175E-22 | 1.13751E-20 | 38.75876778 | Higher in NT |
| SEPTIN11 | -0.25700 | 3.473983679 | -12.0126 | 7.46896E-22 | 1.28775E-20 | 38.62315783 | Higher in NT |
| CCDC92 | -0.59443 | 3.422052722 | -11.9927 | 8.29471E-22 | 1.41387E-20 | 38.51830082 | Higher in NT |
| FLNA.1 | -0.48089 | 4.332486502 | -11.9542 | 1.0169E-21 | 1.71387E-20 | 38.31459371 | Higher in NT |
| FIS1 | -0.29134 | 3.32955021 | -11.9497 | 1.04149E-21 | 1.73582E-20 | 38.29069451 | Higher in NT |
| SMAD5 | -0.67171 | 3.767091744 | -11.8586 | 1.68514E-21 | 2.7777E-20 | 37.80956807 | Higher in NT |
| AIFM1 | -0.37781 | 3.519596807 | -11.8562 | 1.70726E-21 | 2.78358E-20 | 37.79652747 | Higher in NT |
| RHOC | -0.66727 | 3.881388448 | -11.7735 | 2.64448E-21 | 4.26529E-20 | 37.35904206 | Higher in NT |
| SLC9A3R1 | -0.59267 | 4.440805258 | -11.7691 | 2.70715E-21 | 4.31992E-20 | 37.33562512 | Higher in NT |
| DDX6 | -0.62961 | 4.546029653 | -11.7224 | 3.46564E-21 | 5.47206E-20 | 37.0886955 | Higher in NT |
| NDUFA2 | -0.48118 | 2.74781765 | -11.7054 | 3.79354E-21 | 5.92741E-20 | 36.99831973 | Higher in NT |
| SNTA1 | -0.76257 | 3.914450915 | -11.6854 | 4.21783E-21 | 6.52241E-20 | 36.89233619 | Higher in NT |
| RASA1 | -0.32107 | 2.757443461 | -11.6702 | 4.57168E-21 | 6.99747E-20 | 36.81180402 | Higher in NT |
| ABHD5 | -0.53969 | 2.97340952 | -11.5665 | 7.92088E-21 | 1.20013E-19 | 36.26240849 | Higher in NT |
| PLA2G4A | -0.71222 | 3.506034305 | -11.552 | 8.55516E-21 | 1.28327E-19 | 36.1854125 | Higher in NT |
| GRK2 | -0.57875 | 4.186939879 | -11.5444 | 8.90675E-21 | 1.32278E-19 | 36.14515705 | Higher in NT |

| | | | | | | | |
|---|---|---|---|---|---|---|---|
| TPM4 | -0.88387 | 4.490917667 | -11.4917 | 1.17806E-20 | 1.73244E-19 | 35.8656585 | Higher in NT |
| ILK | -0.73337 | 4.013281945 | -11.4489 | 1.47853E-20 | 2.15319E-19 | 35.638607 | Higher in NT |
| ALG2 | -0.36417 | 3.146200056 | -11.4217 | 1.70806E-20 | 2.46355E-19 | 35.49437972 | Higher in NT |
| ENSA | -0.59918 | 3.462198807 | -11.363 | 2.33381E-20 | 3.33402E-19 | 35.1824392 | Higher in NT |
| PFN1 | -0.58571 | 4.598003992 | -11.3554 | 2.42919E-20 | 3.43753E-19 | 35.14241273 | Higher in NT |
| PPCS | -0.49516 | 3.998434649 | -11.2663 | 3.90145E-20 | 5.46932E-19 | 34.66897081 | Higher in NT |
| XRCC4 | -0.29661 | 2.713720298 | -11.2506 | 4.24221E-20 | 5.89196E-19 | 34.58530119 | Higher in NT |
| MEIS2 | -0.50612 | 3.131773543 | -11.2114 | 5.22486E-20 | 7.19017E-19 | 34.37712989 | Higher in NT |
| GNE | -0.24391 | 2.885487972 | -11.206 | 5.37591E-20 | 7.33079E-19 | 34.34865358 | Higher in NT |
| ELMO2 | -0.43019 | 3.457533698 | -11.1868 | 5.9564E-20 | 8.04918E-19 | 34.24620481 | Higher in NT |
| ELK3 | -0.49654 | 3.120531102 | -11.163 | 6.76087E-20 | 9.05474E-19 | 34.11963112 | Higher in NT |
| SCFD1 | -0.40333 | 3.030078408 | -11.1207 | 8.466E-20 | 1.12381E-18 | 33.89493365 | Higher in NT |
| CASP3 | -0.60992 | 4.004694124 | -11.1184 | 8.5713E-20 | 1.1278E-18 | 33.88258499 | Higher in NT |
| HSPA2 | -0.27416 | 3.507941559 | -11.0988 | 9.51144E-20 | 1.24062E-18 | 33.77860922 | Higher in NT |
| PLEK | -0.62443 | 4.87393643 | -11.091 | 9.91765E-20 | 1.28245E-18 | 33.7368302 | Higher in NT |
| MAPRE1 | -0.73196 | 4.329468558 | -11.0289 | 1.38011E-19 | 1.76937E-18 | 33.4067386 | Higher in NT |
| CRK | -0.35104 | 3.830931769 | -10.9739 | 1.85028E-19 | 2.35205E-18 | 33.11388581 | Higher in NT |
| PUS1 | -0.29930 | 2.400722797 | -10.9436 | 2.17402E-19 | 2.74036E-18 | 32.95282902 | Higher in NT |
| NFU1 | -0.34293 | 2.946962052 | -10.9325 | 2.30631E-19 | 2.88288E-18 | 32.89382989 | Higher in NT |
| ANXA11 | -0.56434 | 4.443464014 | -10.9135 | 2.55167E-19 | 3.16323E-18 | 32.79285015 | Higher in NT |
| CRKL | -0.67563 | 3.49820935 | -10.8247 | 4.09673E-19 | 5.03697E-18 | 32.32000693 | Higher in NT |
| GLO1 | 0.35636 | 3.775593797 | 10.79408 | 4.82268E-19 | 5.84833E-18 | 32.15708481 | Higher in PA |
| RAB7A | -0.55740 | 4.396535316 | -10.7936 | 4.83462E-19 | 5.84833E-18 | 32.15461582 | Higher in NT |
| MAPK3 | -0.45724 | 4.120987247 | -10.7017 | 7.88993E-19 | 9.46792E-18 | 31.66552758 | Higher in NT |
| PGAM2 | -0.44062 | 3.150949668 | -10.6818 | 8.77429E-19 | 1.04456E-17 | 31.55944801 | Higher in NT |
| GALK1 | -0.44997 | 3.860447999 | -10.6711 | 9.289E-19 | 1.09713E-17 | 31.50252987 | Higher in NT |
| SARS2 | -0.53539 | 3.4202486 | -10.668 | 9.44426E-19 | 1.10675E-17 | 31.48597867 | Higher in NT |
| GATM | -0.60510 | 3.856678158 | -10.6525 | 1.02616E-18 | 1.19321E-17 | 31.40310646 | Higher in NT |
| HSPA1A | -0.22251 | 3.359059637 | -10.6469 | 1.05703E-18 | 1.21965E-17 | 31.37350832 | Higher in NT |
| NEIL1 | -0.23755 | 4.012264689 | -10.6443 | 1.07178E-18 | 1.22723E-17 | 31.35967548 | Higher in NT |
| EIF2A | -0.16802 | 2.801808498 | -10.6148 | 1.25411E-18 | 1.42513E-17 | 31.20281651 | Higher in NT |
| STAT3 | -0.49230 | 4.220429173 | -10.6082 | 1.29931E-18 | 1.46538E-17 | 31.16747252 | Higher in NT |
| HSPA9 | -0.39668 | 3.567661829 | -10.5686 | 1.60485E-18 | 1.79647E-17 | 30.95661993 | Higher in NT |
| APRT | -0.75460 | 3.756036347 | -10.5592 | 1.68743E-18 | 1.87492E-17 | 30.90652816 | Higher in NT |

| | | | | | | | |
|---|---|---|---|---|---|---|---|
| SLMAP | -0.47201 | 2.962335832 | -10.533 | 1.94079E-18 | 2.14057E-17 | 30.76687678 | Higher in NT |
| KLF9 | -0.47502 | 3.504362759 | -10.5108 | 2.18411E-18 | 2.39136E-17 | 30.64896534 | Higher in NT |
| BPGM | 0.41619 | 3.026228553 | 10.46946 | 2.72331E-18 | 2.96012E-17 | 30.42872069 | Higher in PA |
| AK2 | -0.52687 | 4.517312435 | -10.4408 | 3.17317E-18 | 3.40656E-17 | 30.27611731 | Higher in NT |
| ARHGDIB | -0.50045 | 4.541618107 | -10.4404 | 3.17946E-18 | 3.40656E-17 | 30.27414069 | Higher in NT |
| PKM | -0.71466 | 4.450863409 | -10.3923 | 4.11088E-18 | 4.37328E-17 | 30.01769873 | Higher in NT |
| TBCE | -0.58009 | 3.862813785 | -10.3012 | 6.68215E-18 | 7.0586E-17 | 29.53286469 | Higher in NT |
| FLNA | -0.59144 | 3.078322642 | -10.278 | 7.56392E-18 | 7.93418E-17 | 29.40917219 | Higher in NT |
| PTEN | -0.32121 | 2.793122354 | -10.2487 | 8.84163E-18 | 9.21003E-17 | 29.25342101 | Higher in NT |
| ASAH1 | -0.36463 | 3.106776541 | -10.1167 | 1.78733E-17 | 1.84896E-16 | 28.55116511 | Higher in NT |
| MAPK14 | -0.50402 | 3.552258276 | -10.0823 | 2.14732E-17 | 2.20615E-16 | 28.36810455 | Higher in NT |
| SHC1 | -0.41614 | 3.812152471 | -10.0736 | 2.24957E-17 | 2.29548E-16 | 28.32169847 | Higher in NT |
| ACOX1 | -0.68228 | 4.320244048 | -10.0667 | 2.33339E-17 | 2.36492E-16 | 28.28520449 | Higher in NT |
| TAF15 | -0.31145 | 3.473329841 | -10.052 | 2.52398E-17 | 2.54092E-16 | 28.20688109 | Higher in NT |
| CBL | -0.41610 | 4.115611729 | -10.0185 | 3.01748E-17 | 3.01748E-16 | 28.02874202 | Higher in NT |
| CES3 | -0.51945 | 4.607602041 | -10.0157 | 3.06284E-17 | 3.04255E-16 | 28.01386044 | Higher in NT |
| TACO1 | -0.55213 | 3.320001579 | -9.99932 | 3.34234E-17 | 3.29836E-16 | 27.92675338 | Higher in NT |
| HGD | -0.39609 | 2.982511834 | -9.96663 | 3.97844E-17 | 3.90043E-16 | 27.75298302 | Higher in NT |
| PDPK1 | -0.45188 | 2.871510458 | -9.94462 | 4.47375E-17 | 4.35388E-16 | 27.63595279 | Higher in NT |
| CD84 | -0.18004 | 3.610931709 | -9.94356 | 4.49901E-17 | 4.35388E-16 | 27.63033786 | Higher in NT |
| MAPK8 | -0.27261 | 2.841659164 | -9.91304 | 5.29348E-17 | 5.08989E-16 | 27.46815277 | Higher in NT |
| PYGL | -0.49312 | 4.214389349 | -9.77269 | 1.11797E-16 | 1.06813E-15 | 26.72263773 | Higher in NT |
| S100A6 | 0.34606 | 3.501991806 | 9.730985 | 1.39592E-16 | 1.32524E-15 | 26.50126659 | Higher in PA |
| LONP1 | -0.40371 | 2.977336582 | -9.69381 | 1.70133E-16 | 1.60503E-15 | 26.30401305 | Higher in NT |
| FTH1 | -0.16965 | 3.345383065 | -9.68381 | 1.79434E-16 | 1.6822E-15 | 26.2509483 | Higher in NT |
| FXN | -0.21366 | 2.842194105 | -9.64626 | 2.1912E-16 | 2.04149E-15 | 26.0517704 | Higher in NT |
| STAMBP | -0.29802 | 3.32881084 | -9.64437 | 2.2133E-16 | 2.04935E-15 | 26.04176489 | Higher in NT |
| CFAP298 | -0.27128 | 2.669214531 | -9.60522 | 2.72575E-16 | 2.50836E-15 | 25.83417751 | Higher in NT |
| SUMO4 | -0.26984 | 3.188138883 | -9.59792 | 2.83358E-16 | 2.59169E-15 | 25.79550662 | Higher in NT |
| DLG4 | -0.30580 | 2.629089188 | -9.5795 | 3.12519E-16 | 2.84108E-15 | 25.69787507 | Higher in NT |
| FOXO3 | -0.23030 | 2.931055771 | -9.55691 | 3.52408E-16 | 3.18441E-15 | 25.57815265 | Higher in NT |
| KLRC4 | -0.23630 | 2.759763371 | -9.53651 | 3.92765E-16 | 3.52783E-15 | 25.47009764 | Higher in NT |
| IL20 | -0.27728 | 2.617530109 | -9.5351 | 3.95716E-16 | 3.53318E-15 | 25.46263673 | Higher in NT |
| RAB1B | -0.78317 | 3.587737569 | -9.51975 | 4.29365E-16 | 3.81093E-15 | 25.38130492 | Higher in NT |

| | | | | | | | |
|---|---|---|---|---|---|---|---|
| BAD | -0.39163 | 3.641710899 | -9.51257 | 4.46076E-16 | 3.93596E-15 | 25.34325545 | Higher in NT |
| PRKAG1\|PRKAB2 | -0.47672 | 3.507255696 | -9.50907 | 4.54426E-16 | 3.9862E-15 | 25.32477234 | Higher in NT |
| SMAD4 | -0.33849 | 2.629967305 | -9.50313 | 4.69013E-16 | 4.09023E-15 | 25.29328598 | Higher in NT |
| EGF.1 | -0.45227 | 3.191127445 | -9.48445 | 5.17957E-16 | 4.49096E-15 | 25.19437089 | Higher in NT |
| CAVIN1 | -0.32456 | 3.414281334 | -9.45406 | 6.08687E-16 | 5.2473E-15 | 25.03352974 | Higher in NT |
| CDK8\|CCNC | -0.19258 | 3.226770235 | -9.44969 | 6.22997E-16 | 5.33997E-15 | 25.01037463 | Higher in NT |
| GFER | -0.30056 | 3.065913978 | -9.37619 | 9.20343E-16 | 7.84383E-15 | 24.62159386 | Higher in NT |
| STAT1 | -0.44990 | 4.471150854 | -9.36836 | 9.59421E-16 | 8.13068E-15 | 24.58016548 | Higher in NT |
| VCP | -0.51804 | 4.416862645 | -9.34674 | 1.07605E-15 | 9.06786E-15 | 24.46587244 | Higher in NT |
| DSTN | -0.60705 | 3.620786975 | -9.33946 | 1.1184E-15 | 9.37204E-15 | 24.42742154 | Higher in NT |
| GNAQ | -0.40988 | 3.421362493 | -9.33792 | 1.12759E-15 | 9.39659E-15 | 24.41926641 | Higher in NT |
| G6PD | -0.63227 | 4.142221819 | -9.32413 | 1.21314E-15 | 1.00537E-14 | 24.34641259 | Higher in NT |
| CCM2 | -0.19476 | 3.508571898 | -9.31031 | 1.30537E-15 | 1.07586E-14 | 24.27342039 | Higher in NT |
| LDLRAP1 | -0.64538 | 4.36341861 | -9.21157 | 2.20312E-15 | 1.80584E-14 | 23.75211223 | Higher in NT |
| PKLR | 0.41125 | 3.922905999 | 9.161299 | 2.87508E-15 | 2.34382E-14 | 23.48700874 | Higher in PA |
| RRAS | -0.17124 | 2.980829263 | -9.12418 | 3.49924E-15 | 2.83722E-14 | 23.29137462 | Higher in NT |
| PSMD6 | -0.24691 | 3.283342209 | -9.11465 | 3.68013E-15 | 2.96785E-14 | 23.24118881 | Higher in NT |
| ALDOA | -0.47896 | 3.162097209 | -9.0918 | 4.15296E-15 | 3.33125E-14 | 23.1208392 | Higher in NT |
| ROCK2 | -0.48634 | 3.121387496 | -9.06488 | 4.78831E-15 | 3.82046E-14 | 22.97910468 | Higher in NT |
| PRKCB | -0.54080 | 4.606664018 | -9.0475 | 5.24902E-15 | 4.16589E-14 | 22.88764968 | Higher in NT |
| SRC | -0.80237 | 3.89668897 | -9.02681 | 5.85557E-15 | 4.62282E-14 | 22.77878688 | Higher in NT |
| BRAF | -0.17635 | 3.136766098 | -8.98205 | 7.41731E-15 | 5.82511E-14 | 22.54344003 | Higher in NT |
| CBR1 | -0.18832 | 3.374907613 | -8.95732 | 8.45167E-15 | 6.60287E-14 | 22.41349786 | Higher in NT |
| NACA | -0.41228 | 3.871655239 | -8.95262 | 8.66397E-15 | 6.73365E-14 | 22.38880476 | Higher in NT |
| IL18 | -0.24541 | 2.926119003 | -8.94916 | 8.82361E-15 | 6.82238E-14 | 22.37063245 | Higher in NT |
| MAP2K1 | -0.21547 | 2.607332807 | -8.9447 | 9.03358E-15 | 6.94891E-14 | 22.34722452 | Higher in NT |
| IRAK4 | -0.29974 | 2.819040851 | -8.94195 | 9.16581E-15 | 7.01465E-14 | 22.33276188 | Higher in NT |
| PGK1 | -0.19943 | 3.420502976 | -8.86953 | 1.34291E-14 | 1.02252E-13 | 21.95264969 | Higher in NT |
| SMAD1 | -0.24952 | 3.089918468 | -8.8506 | 1.48373E-14 | 1.12404E-13 | 21.85341571 | Higher in NT |
| FLT3 | -0.27019 | 3.334375796 | -8.78589 | 2.08616E-14 | 1.57248E-13 | 21.51435783 | Higher in NT |
| TALDO1 | -0.22703 | 4.007220794 | -8.76077 | 2.38091E-14 | 1.78569E-13 | 21.38287683 | Higher in NT |
| SULT1A1 | -0.49780 | 3.574316842 | -8.75261 | 2.48534E-14 | 1.85473E-13 | 21.34017475 | Higher in NT |
| LDHA | -0.24985 | 4.597666135 | -8.72144 | 2.928E-14 | 2.16637E-13 | 21.17711939 | Higher in NT |
| CXCL2 | -0.41289 | 3.777540366 | -8.72119 | 2.93182E-14 | 2.16637E-13 | 21.1758233 | Higher in NT |

| | | | | | | | |
|---|---|---|---|---|---|---|---|
| SHC1.1 | -0.32518 | 3.626899425 | -8.71555 | 3.02002E-14 | 2.2206E-13 | 21.14633754 | Higher in NT |
| B4GALT1 | 0.12162 | 4.310085612 | 8.712844 | 3.06332E-14 | 2.24146E-13 | 21.13217685 | Higher in PA |
| NONO | -0.26681 | 2.870939052 | -8.70297 | 3.22655E-14 | 2.34943E-13 | 21.0805386 | Higher in NT |
| SPARC | -0.18315 | 3.533087943 | -8.65221 | 4.21216E-14 | 3.05229E-13 | 20.81540974 | Higher in NT |
| S100A5 | -0.39188 | 3.386805355 | -8.49455 | 9.62401E-14 | 6.94039E-13 | 19.99380029 | Higher in NT |
| CD14 | 0.25130 | 4.123630358 | 8.449207 | 1.21992E-13 | 8.7554E-13 | 19.75810203 | Higher in PA |
| ALAD | -0.30228 | 4.145441992 | -8.44041 | 1.27733E-13 | 9.12377E-13 | 19.71239314 | Higher in NT |
| TARDBP | 0.10961 | 2.660136246 | 8.439053 | 1.2864E-13 | 9.14501E-13 | 19.70536025 | Higher in PA |
| POR | -0.26643 | 3.491369909 | -8.3806 | 1.74557E-13 | 1.23508E-12 | 19.40199261 | Higher in NT |
| SMAD2 | -0.40331 | 3.653003146 | -8.3485 | 2.06373E-13 | 1.45333E-12 | 19.23560806 | Higher in NT |
| NME1 | -0.27362 | 3.383865147 | -8.34526 | 2.09888E-13 | 1.47118E-12 | 19.21882797 | Higher in NT |
| TPI1 | -0.43224 | 4.118753026 | -8.34063 | 2.15017E-13 | 1.50012E-12 | 19.19483686 | Higher in NT |
| PTPN11 | -0.34897 | 4.4540227 | -8.33154 | 2.25443E-13 | 1.56558E-12 | 19.14778471 | Higher in NT |
| SELE | 0.23911 | 4.278986765 | 8.329297 | 2.28098E-13 | 1.57672E-12 | 19.13615076 | Higher in PA |
| CHMP2B | -0.52015 | 4.113405637 | -8.32841 | 2.29156E-13 | 1.57676E-12 | 19.13155448 | Higher in NT |
| SFRP1 | 0.27650 | 3.181427874 | 8.316029 | 2.44427E-13 | 1.67416E-12 | 19.06745389 | Higher in PA |
| GPT | -0.23925 | 3.479916221 | -8.29713 | 2.69712E-13 | 1.83894E-12 | 18.96964827 | Higher in NT |
| BCL2L1 | -0.20896 | 3.497499379 | -8.28153 | 2.92524E-13 | 1.98545E-12 | 18.88898396 | Higher in NT |
| MMP7 | 0.34645 | 3.926450676 | 8.270492 | 3.09827E-13 | 2.09343E-12 | 18.83188993 | Higher in PA |
| PDLIM5 | -0.36942 | 4.408459548 | -8.26517 | 3.18527E-13 | 2.14256E-12 | 18.80437972 | Higher in NT |
| HMBS | 0.35665 | 4.544200984 | 8.261484 | 3.24695E-13 | 2.1743E-12 | 18.78532562 | Higher in PA |
| LRIG1 | 0.13716 | 2.927312086 | 8.256098 | 3.3392E-13 | 2.22613E-12 | 18.75749499 | Higher in PA |
| FN1.1 | -0.83594 | 4.598183791 | -8.24242 | 3.58547E-13 | 2.37974E-12 | 18.68680558 | Higher in NT |
| ALCAM | 0.10596 | 3.997630295 | 8.237279 | 3.68253E-13 | 2.43339E-12 | 18.66027458 | Higher in PA |
| CD68 | -0.19452 | 3.214891985 | -8.22791 | 3.86631E-13 | 2.54363E-12 | 18.61189711 | Higher in NT |
| RAC1 | -0.54845 | 4.42233415 | -8.21897 | 4.0502E-13 | 2.65297E-12 | 18.56574381 | Higher in NT |
| IL6 | -0.32593 | 2.756546153 | -8.20236 | 4.41516E-13 | 2.87945E-12 | 18.48004623 | Higher in NT |
| FASN | -0.32904 | 3.317212785 | -8.17702 | 5.03627E-13 | 3.27031E-12 | 18.34931873 | Higher in NT |
| PLXND1 | 0.13622 | 3.724093216 | 8.174495 | 5.10261E-13 | 3.2991E-12 | 18.33632116 | Higher in PA |
| LDHB | -0.33532 | 3.759791617 | -8.17211 | 5.16606E-13 | 3.32579E-12 | 18.32404822 | Higher in NT |
| GBP2 | -0.17560 | 3.240625612 | -8.17098 | 5.19658E-13 | 3.33114E-12 | 18.31819777 | Higher in NT |
| ITGB5\|ITGAV | -0.17234 | 3.02010938 | -8.15607 | 5.61455E-13 | 3.58375E-12 | 18.24137035 | Higher in NT |
| RAB5A | -0.36645 | 4.013160299 | -8.14238 | 6.02763E-13 | 3.83112E-12 | 18.17086912 | Higher in NT |
| FN1 | -0.78916 | 3.965052978 | -8.12642 | 6.54774E-13 | 4.14414E-12 | 18.08868031 | Higher in NT |

| | | | | | | | |
|---|---|---|---|---|---|---|---|
| CA1 | 0.29664 | 4.417428456 | 8.124842 | 6.6015E-13 | 4.16061E-12 | 18.08056 | Higher in PA |
| CTNNA1 | -0.38016 | 3.225791718 | -8.09833 | 7.57349E-13 | 4.75324E-12 | 17.94417136 | Higher in NT |
| PTH.1 | 0.33001 | 3.078030264 | 8.097506 | 7.60598E-13 | 4.75374E-12 | 17.9399219 | Higher in PA |
| MLYCD | -0.18933 | 2.845311258 | -8.09234 | 7.81224E-13 | 4.86239E-12 | 17.91335406 | Higher in NT |
| SDHAF1 | -0.13615 | 3.038459486 | -8.08848 | 7.96972E-13 | 4.93991E-12 | 17.89353937 | Higher in NT |
| STAR | -0.20111 | 3.096257617 | -8.08048 | 8.30689E-13 | 5.12771E-12 | 17.85239861 | Higher in NT |
| TKT | -0.34092 | 4.629664944 | -8.07923 | 8.36098E-13 | 5.13995E-12 | 17.84595458 | Higher in NT |
| PPM1B | -0.30229 | 3.024136467 | -8.04635 | 9.91159E-13 | 6.06832E-12 | 17.67705262 | Higher in NT |
| NDP | 0.17888 | 2.604861057 | 7.938243 | 1.73196E-12 | 1.05488E-11 | 17.12307407 | Higher in PA |
| DLAT | -0.21931 | 2.867668143 | -7.93768 | 1.73704E-12 | 1.05488E-11 | 17.12017211 | Higher in NT |
| STX6 | -0.12416 | 3.325410108 | -7.91715 | 1.93083E-12 | 1.16541E-11 | 17.01521383 | Higher in NT |
| NFKB1 | -0.34244 | 3.68870653 | -7.91677 | 1.93458E-12 | 1.16541E-11 | 17.0132893 | Higher in NT |
| IL15 | -0.18275 | 3.048188162 | -7.91471 | 1.95525E-12 | 1.17315E-11 | 17.00274133 | Higher in NT |
| RAC2 | -0.44245 | 4.243532673 | -7.89323 | 2.18386E-12 | 1.3051E-11 | 16.89302336 | Higher in NT |
| SELP | -0.30507 | 4.408970086 | -7.88934 | 2.22805E-12 | 1.32622E-11 | 16.87314735 | Higher in NT |
| GCDH | -0.33807 | 2.919565866 | -7.87705 | 2.37351E-12 | 1.40722E-11 | 16.81040202 | Higher in NT |
| GNAI3 | -0.12861 | 2.803802467 | -7.85452 | 2.66503E-12 | 1.57384E-11 | 16.69546919 | Higher in NT |
| NCAM1 | 0.12595 | 3.896520137 | 7.846714 | 2.77415E-12 | 1.63185E-11 | 16.65565854 | Higher in PA |
| GNS | -0.23858 | 3.060150018 | -7.83476 | 2.94983E-12 | 1.72842E-11 | 16.59474501 | Higher in NT |
| YWHAH | -0.33412 | 4.846184596 | -7.82808 | 3.05292E-12 | 1.78186E-11 | 16.56067058 | Higher in NT |
| DIABLO | -0.12434 | 3.51061311 | -7.78061 | 3.89492E-12 | 2.26449E-11 | 16.31907065 | Higher in NT |
| PPP3R1 | -0.19423 | 3.697189381 | -7.77621 | 3.98377E-12 | 2.3072E-11 | 16.29670253 | Higher in NT |
| SDHB | -0.17317 | 3.198336008 | -7.72491 | 5.18105E-12 | 2.98907E-11 | 16.03610655 | Higher in NT |
| P4HB | -0.18084 | 3.627377078 | -7.69361 | 6.08073E-12 | 3.49467E-11 | 15.87734697 | Higher in NT |
| SPINT2 | -0.21200 | 2.83599287 | -7.66864 | 6.90815E-12 | 3.95505E-11 | 15.75086396 | Higher in NT |
| LSP1 | 0.16883 | 2.770721724 | 7.600778 | 9.76577E-12 | 5.56983E-11 | 15.40771503 | Higher in PA |
| PGLS | -0.37545 | 3.521402246 | -7.59486 | 1.00648E-11 | 5.71863E-11 | 15.37782426 | Higher in NT |
| GM2A | 0.13176 | 3.033611013 | 7.542597 | 1.31308E-11 | 7.43253E-11 | 15.11430523 | Higher in PA |
| CA3 | 0.29422 | 3.256816937 | 7.539769 | 1.33209E-11 | 7.51176E-11 | 15.10006597 | Higher in PA |
| HRAS | -0.16939 | 3.180223158 | -7.52914 | 1.40601E-11 | 7.89892E-11 | 15.04655247 | Higher in NT |
| PDGFB | -0.59371 | 4.421692739 | -7.48775 | 1.73477E-11 | 9.70952E-11 | 14.83838002 | Higher in NT |
| COMT | 0.15067 | 3.1051932 | 7.468727 | 1.91042E-11 | 1.06529E-10 | 14.74283505 | Higher in PA |
| NR0B2 | -0.40359 | 2.305833981 | -7.45409 | 2.05746E-11 | 1.14303E-10 | 14.66938721 | Higher in NT |
| GLB1 | -0.34869 | 1.32504848 | -7.45237 | 2.07547E-11 | 1.14878E-10 | 14.66075689 | Higher in NT |

| | | | | | | | |
|---|---|---|---|---|---|---|---|
| CBS | -0.18167 | 3.160252844 | -7.41697 | 2.48275E-11 | 1.36916E-10 | 14.48329112 | Higher in NT |
| PNP | -0.40647 | 3.785611415 | -7.38972 | 2.84945E-11 | 1.56563E-10 | 14.34687079 | Higher in NT |
| TXNRD1 | -0.32249 | 3.676168207 | -7.35876 | 3.33145E-11 | 1.82379E-10 | 14.1921263 | Higher in NT |
| IGFBP7 | 0.10265 | 4.485899264 | 7.341139 | 3.64109E-11 | 1.98605E-10 | 14.10413389 | Higher in PA |
| VWF | -0.41028 | 4.425396089 | -7.33134 | 3.82551E-11 | 2.07908E-10 | 14.05521992 | Higher in NT |
| IL2 | 0.13153 | 3.016000209 | 7.322227 | 4.00517E-11 | 2.16886E-10 | 14.0097865 | Higher in PA |
| GSN | -0.14194 | 2.979556909 | -7.27969 | 4.96116E-11 | 2.67688E-10 | 13.797906 | Higher in NT |
| QDPR | -0.30508 | 3.822378126 | -7.27722 | 5.02331E-11 | 2.7007E-10 | 13.78558514 | Higher in NT |
| GAA | 0.14184 | 4.041375724 | 7.27258 | 5.14177E-11 | 2.75452E-10 | 13.76251483 | Higher in PA |
| CXCL3 | -0.43156 | 3.638691445 | -7.25774 | 5.53977E-11 | 2.95717E-10 | 13.68872856 | Higher in NT |
| HNRNPA1 | -0.43862 | 3.592819404 | -7.17091 | 8.56116E-11 | 4.55381E-10 | 13.25803885 | Higher in NT |
| HAVCR1 | 0.25461 | 2.964302161 | 7.152563 | 9.38396E-11 | 4.97383E-10 | 13.16726389 | Higher in PA |
| ITPA | -0.25284 | 3.733128354 | -7.12054 | 1.10116E-10 | 5.816E-10 | 13.00905964 | Higher in NT |
| CMIP | -0.29331 | 3.27802458 | -7.05831 | 1.50157E-10 | 7.90299E-10 | 12.70237296 | Higher in NT |
| TNFRSF1B | 0.15029 | 3.773452803 | 7.057562 | 1.50715E-10 | 7.90465E-10 | 12.69870174 | Higher in PA |
| NOTCH1 | 0.08392 | 4.107326097 | 7.003747 | 1.96923E-10 | 1.02922E-09 | 12.43433888 | Higher in PA |
| UGT1A1 | -0.15046 | 3.009422413 | -6.99762 | 2.02998E-10 | 1.05728E-09 | 12.40431043 | Higher in NT |
| IL6ST | 0.10384 | 3.839797361 | 6.983785 | 2.17419E-10 | 1.12847E-09 | 12.33647451 | Higher in PA |
| NOTCH3 | 0.11023 | 4.575814179 | 6.98049 | 2.21E-10 | 1.1431E-09 | 12.32033199 | Higher in PA |
| SDHAF2 | -0.14768 | 2.399005894 | -6.97659 | 2.25317E-10 | 1.16143E-09 | 12.30121082 | Higher in NT |
| ERBB4 | -0.10085 | 2.246581318 | -6.97207 | 2.3042E-10 | 1.18366E-09 | 12.27907954 | Higher in NT |
| PRSS8 | 0.11508 | 3.135115582 | 6.929435 | 2.84545E-10 | 1.45672E-09 | 12.07058742 | Higher in PA |
| PYCARD | -0.13415 | 4.150535261 | -6.92721 | 2.87699E-10 | 1.46785E-09 | 12.05969911 | Higher in NT |
| F5 | -0.12812 | 4.360035303 | -6.87043 | 3.80755E-10 | 1.93604E-09 | 11.78285023 | Higher in NT |
| PF4 | -0.55195 | 4.516597289 | -6.85603 | 4.08744E-10 | 2.07134E-09 | 11.71278821 | Higher in NT |
| F3 | 0.13076 | 2.91768962 | 6.851597 | 4.17761E-10 | 2.1099E-09 | 11.69123943 | Higher in PA |
| NISCH | -0.10129 | 3.358106542 | -6.84561 | 4.30254E-10 | 2.16571E-09 | 11.66213627 | Higher in NT |
| CYRIB | -0.29964 | 3.346207112 | -6.83109 | 4.62103E-10 | 2.31824E-09 | 11.59161206 | Higher in NT |
| MANBA | -0.15386 | 3.505902314 | -6.81389 | 5.02872E-10 | 2.51436E-09 | 11.50812226 | Higher in NT |
| YES1 | -0.15769 | 2.826471538 | -6.80022 | 5.37799E-10 | 2.68006E-09 | 11.44182073 | Higher in NT |
| SERPINA1 | 0.12797 | 4.515474151 | 6.710135 | 8.36014E-10 | 4.15239E-09 | 11.00634579 | Higher in PA |
| ITGB2\|ITGAL | -0.13229 | 2.424479983 | -6.70429 | 8.60235E-10 | 4.25859E-09 | 10.97816147 | Higher in NT |
| HDAC8 | -0.12088 | 3.917282303 | -6.67653 | 9.85E-10 | 4.8602E-09 | 10.84452113 | Higher in NT |
| KRAS | -0.19654 | 2.167116328 | -6.65484 | 1.09476E-09 | 5.38406E-09 | 10.74028889 | Higher in NT |

| | | | | | | | |
|---|---|---|---|---|---|---|---|
| ATOX1 | -0.22598 | 2.971733229 | -6.65161 | 1.11214E-09 | 5.45165E-09 | 10.72475243 | Higher in NT |
| TNFRSF1A | 0.21017 | 3.913609545 | 6.624936 | 1.2662E-09 | 6.18663E-09 | 10.59677759 | Higher in PA |
| STK4 | -0.22419 | 3.607782422 | -6.60883 | 1.36922E-09 | 6.66828E-09 | 10.51962381 | Higher in NT |
| CAP1 | -0.48719 | 3.927724991 | -6.57368 | 1.62379E-09 | 7.88249E-09 | 10.35146141 | Higher in NT |
| CD27 | -0.10628 | 3.582627005 | -6.57004 | 1.65265E-09 | 7.99669E-09 | 10.33409149 | Higher in NT |
| ACE | 0.13719 | 2.777545893 | 6.566583 | 1.68056E-09 | 8.10557E-09 | 10.31758136 | Higher in PA |
| CTHRC1 | 0.13693 | 4.650248325 | 6.532555 | 1.9813E-09 | 9.52546E-09 | 10.15527468 | Higher in PA |
| SPP1 | 0.15103 | 2.804645468 | 6.52749 | 2.03038E-09 | 9.73025E-09 | 10.1311507 | Higher in PA |
| CD163 | 0.16159 | 3.329148287 | 6.510268 | 2.20645E-09 | 1.05404E-08 | 10.04917419 | Higher in PA |
| FEV | -0.19941 | 2.861929759 | -6.50727 | 2.23858E-09 | 1.06599E-08 | 10.03492524 | Higher in NT |
| TAGLN | 0.16941 | 3.271667265 | 6.500885 | 2.30863E-09 | 1.09587E-08 | 10.00455771 | Higher in PA |
| HBB\|HBA1 | 0.40006 | 2.935764063 | 6.480337 | 2.549E-09 | 1.20615E-08 | 9.906941958 | Higher in PA |
| GP1BB | -0.28880 | 3.400790035 | -6.4796 | 2.55811E-09 | 1.20665E-08 | 9.903428533 | Higher in NT |
| ATIC | -0.35140 | 3.577887552 | -6.4669 | 2.7193E-09 | 1.27867E-08 | 9.843209614 | Higher in NT |
| SCO2 | -0.27905 | 2.725198512 | -6.46368 | 2.76177E-09 | 1.29458E-08 | 9.827939222 | Higher in NT |
| PDLIM4 | 0.19794 | 2.757422305 | 6.438162 | 3.12235E-09 | 1.45904E-08 | 9.70702959 | Higher in PA |
| FAM20A | 0.16745 | 3.102092957 | 6.429768 | 3.2508E-09 | 1.51435E-08 | 9.66731263 | Higher in PA |
| PNPO | -0.20740 | 3.071366603 | -6.42733 | 3.28899E-09 | 1.5274E-08 | 9.655806275 | Higher in NT |
| TYK2 | -0.16312 | 3.438744224 | -6.40645 | 3.63552E-09 | 1.68311E-08 | 9.557130197 | Higher in NT |
| CXCL16 | 0.09653 | 3.674748583 | 6.394033 | 3.85851E-09 | 1.78085E-08 | 9.498497254 | Higher in PA |
| VDR | -0.09883 | 2.697219241 | -6.39007 | 3.93242E-09 | 1.80939E-08 | 9.479810841 | Higher in NT |
| CNTN1 | 0.10417 | 4.463469699 | 6.382563 | 4.07634E-09 | 1.86988E-08 | 9.444409025 | Higher in PA |
| PTPN1 | -0.20774 | 3.1230996 | -6.36885 | 4.35284E-09 | 1.99063E-08 | 9.379777706 | Higher in NT |
| LAMA4 | -0.07342 | 2.804832322 | -6.35184 | 4.72145E-09 | 2.15264E-08 | 9.299733282 | Higher in NT |
| DNAJC19 | -0.16030 | 2.831438136 | -6.33698 | 5.06866E-09 | 2.30394E-08 | 9.229865361 | Higher in NT |
| SPARCL1 | 0.11920 | 3.914875912 | 6.319103 | 5.51966E-09 | 2.50136E-08 | 9.145948247 | Higher in PA |
| KDR | 0.09251 | 3.756449357 | 6.316466 | 5.58946E-09 | 2.52536E-08 | 9.133577539 | Higher in PA |
| TAPBP | -0.29052 | 3.539398782 | -6.31513 | 5.62504E-09 | 2.5338E-08 | 9.127332051 | Higher in NT |
| ID1 | -0.14619 | 2.599592312 | -6.29292 | 6.25251E-09 | 2.80801E-08 | 9.0232349 | Higher in NT |
| APLN | -0.08117 | 2.772277488 | -6.26633 | 7.09505E-09 | 3.17689E-08 | 8.898823759 | Higher in NT |
| NRAS | -0.13244 | 3.245999875 | -6.2484 | 7.72501E-09 | 3.44867E-08 | 8.815118626 | Higher in NT |
| MET | 0.08662 | 3.199235288 | 6.166461 | 1.13816E-08 | 5.066E-08 | 8.433919019 | Higher in PA |
| IL2RG | -0.21200 | 3.567412369 | -6.14553 | 1.25615E-08 | 5.57463E-08 | 8.336931492 | Higher in NT |
| CFD | 0.12758 | 4.51861828 | 6.112024 | 1.4705E-08 | 6.50663E-08 | 8.182046586 | Higher in PA |

| | | | | | | | |
|---|---|---|---|---|---|---|---|
| MAPK9 | -0.16684 | 3.749329933 | -6.07236 | 1.77113E-08 | 7.8138E-08 | 7.999229647 | Higher in NT |
| IL21R | -0.12720 | 3.0330129 | -6.07139 | 1.77921E-08 | 7.82644E-08 | 7.994756161 | Higher in NT |
| CCN2 | -0.09958 | 3.129998687 | -6.05929 | 1.88283E-08 | 8.25803E-08 | 7.939133297 | Higher in NT |
| TEK | 0.09163 | 3.030317353 | 6.053557 | 1.93403E-08 | 8.45785E-08 | 7.912773159 | Higher in PA |
| DKK1 | -0.26005 | 3.829918332 | -6.02999 | 2.15914E-08 | 9.41487E-08 | 7.804601763 | Higher in NT |
| IRF5 | -0.07447 | 3.11360693 | -6.028 | 2.17926E-08 | 9.47504E-08 | 7.795492658 | Higher in NT |
| FKBP6 | -0.13283 | 3.343447153 | -5.96179 | 2.96601E-08 | 1.28584E-07 | 7.492790713 | Higher in NT |
| CCL17 | -0.18350 | 2.684280663 | -5.94059 | 3.27253E-08 | 1.41464E-07 | 7.396242554 | Higher in NT |
| DHFR | -0.18725 | 3.164426211 | -5.92864 | 3.45887E-08 | 1.49089E-07 | 7.341881607 | Higher in NT |
| AREG | -0.17290 | 3.331779067 | -5.92603 | 3.50093E-08 | 1.5047E-07 | 7.330018857 | Higher in NT |
| CNTF | 0.05929 | 2.227120957 | 5.918482 | 3.62538E-08 | 1.55374E-07 | 7.295734736 | Higher in PA |
| RFX5 | 0.10885 | 2.925910002 | 5.912175 | 3.73272E-08 | 1.59518E-07 | 7.267099424 | Higher in PA |
| JAG1 | 0.15704 | 3.203496266 | 5.884923 | 4.23353E-08 | 1.80406E-07 | 7.143556876 | Higher in PA |
| EPHA4 | 0.09320 | 4.19039827 | 5.860888 | 4.72961E-08 | 2.00975E-07 | 7.034846754 | Higher in PA |
| PLG.2 | -0.09149 | 3.885102594 | -5.84968 | 4.98009E-08 | 2.11021E-07 | 6.984226306 | Higher in NT |
| PLEKHM2 | 0.19867 | 3.492454429 | 5.826785 | 5.53281E-08 | 2.33781E-07 | 6.881000658 | Higher in PA |
| MDH1 | -0.22244 | 4.383136735 | -5.82506 | 5.57685E-08 | 2.3498E-07 | 6.873225385 | Higher in NT |
| MMP2 | 0.10193 | 4.186309364 | 5.809262 | 5.99614E-08 | 2.51939E-07 | 6.802138053 | Higher in PA |
| ABCC6 | 0.06574 | 2.83670623 | 5.789435 | 6.56643E-08 | 2.7513E-07 | 6.713059735 | Higher in PA |
| SNCA | -0.25153 | 3.90932072 | -5.7666 | 7.28937E-08 | 3.0457E-07 | 6.610673206 | Higher in NT |
| SCARB1 | 0.14849 | 3.034649904 | 5.75271 | 7.76678E-08 | 3.23616E-07 | 6.548496973 | Higher in PA |
| CYP3A4 | -0.12149 | 3.580666113 | -5.72378 | 8.86184E-08 | 3.6822E-07 | 6.419246843 | Higher in NT |
| PPP1R1A | 0.09224 | 2.990457721 | 5.716709 | 9.15149E-08 | 3.79206E-07 | 6.387734826 | Higher in PA |
| RRM2B | -0.11302 | 3.178481585 | -5.7098 | 9.44359E-08 | 3.90231E-07 | 6.356953609 | Higher in NT |
| EPHB4 | 0.18188 | 3.567410657 | 5.702671 | 9.75476E-08 | 4.01982E-07 | 6.325194344 | Higher in PA |
| CD93 | 0.09682 | 3.679716095 | 5.701711 | 9.79742E-08 | 4.02634E-07 | 6.320920225 | Higher in PA |
| KIAA0319L | -0.06474 | 2.906345007 | -5.6912 | 1.02765E-07 | 4.21168E-07 | 6.274156641 | Higher in NT |
| EPHX2 | -0.06978 | 2.414517658 | -5.66224 | 1.17186E-07 | 4.78963E-07 | 6.145544452 | Higher in NT |
| HBEGF | -0.30957 | 3.301038941 | -5.66077 | 1.1797E-07 | 4.80855E-07 | 6.139018791 | Higher in NT |
| NPY | -0.12289 | 3.223632491 | -5.62805 | 1.36779E-07 | 5.56013E-07 | 5.994175768 | Higher in NT |
| CTSS | 0.16667 | 4.063014401 | 5.622027 | 1.40547E-07 | 5.69783E-07 | 5.967578847 | Higher in PA |
| C1R | -0.15846 | 4.966484243 | -5.61713 | 1.43684E-07 | 5.80935E-07 | 5.945964813 | Higher in NT |
| CDH5 | 0.09035 | 4.12003771 | 5.590747 | 1.6182E-07 | 6.52502E-07 | 5.829629944 | Higher in PA |
| IGFBP6 | 0.09219 | 4.267192573 | 5.578839 | 1.70723E-07 | 6.86552E-07 | 5.777228232 | Higher in PA |

| | | | | | | | |
|---|---|---|---|---|---|---|---|
| PDYN | -0.14181 | 3.514550619 | -5.57695 | 1.72179E-07 | 6.90559E-07 | 5.768914021 | Higher in NT |
| MGAT2 | 0.07154 | 3.005291346 | 5.571218 | 1.76669E-07 | 7.06678E-07 | 5.743726525 | Higher in PA |
| BDNF | -0.43339 | 3.794554737 | -5.52603 | 2.16333E-07 | 8.63029E-07 | 5.545612769 | Higher in NT |
| PDGFA | -0.30599 | 3.72907768 | -5.52522 | 2.17117E-07 | 8.63859E-07 | 5.542074883 | Higher in NT |
| MDM2 | -0.14511 | 2.687029495 | -5.52024 | 2.22007E-07 | 8.80978E-07 | 5.520294721 | Higher in NT |
| PNKP | -0.24797 | 2.987688469 | -5.49158 | 2.52314E-07 | 9.98603E-07 | 5.395175981 | Higher in NT |
| SERPINA10 | 0.10975 | 3.823130582 | 5.469402 | 2.78507E-07 | 1.09937E-06 | 5.298628697 | Higher in PA |
| NODAL | 0.06797 | 2.442738775 | 5.455424 | 2.96365E-07 | 1.16679E-06 | 5.237892699 | Higher in PA |
| LGALS3 | 0.08530 | 3.8093149 | 5.450051 | 3.03523E-07 | 1.19184E-06 | 5.214569145 | Higher in PA |
| CFHR5 | 0.09780 | 4.740791976 | 5.447476 | 3.07013E-07 | 1.2024E-06 | 5.2033963 | Higher in PA |
| ARL6 | -0.27339 | 3.456855365 | -5.44074 | 3.16335E-07 | 1.23568E-06 | 5.174170213 | Higher in NT |
| IL16 | 0.12701 | 4.219469048 | 5.434258 | 3.25557E-07 | 1.26841E-06 | 5.14609253 | Higher in PA |
| GAPDH | -0.36926 | 3.77650311 | -5.43156 | 3.29469E-07 | 1.28032E-06 | 5.13442326 | Higher in NT |
| VIP | -0.09934 | 2.338327335 | -5.42968 | 3.32225E-07 | 1.2877E-06 | 5.126283929 | Higher in NT |
| CHL1 | 0.09330 | 3.961016037 | 5.424781 | 3.39523E-07 | 1.31259E-06 | 5.105055873 | Higher in PA |
| SERPING1 | 0.10858 | 3.787886698 | 5.4103 | 3.62002E-07 | 1.39589E-06 | 5.042434105 | Higher in PA |
| CCL5 | -0.32977 | 4.443316875 | -5.39654 | 3.8471E-07 | 1.47965E-06 | 4.983010924 | Higher in NT |
| GBA | 0.18261 | 3.634207017 | 5.386419 | 4.02288E-07 | 1.5433E-06 | 4.939378274 | Higher in PA |
| ARG1 | 0.27220 | 3.156227298 | 5.372354 | 4.28034E-07 | 1.63788E-06 | 4.878805844 | Higher in PA |
| LMNB2 | 0.17090 | 3.302803162 | 5.37047 | 4.31603E-07 | 1.64734E-06 | 4.8706974 | Higher in PA |
| RNASEH1 | 0.12167 | 2.166472283 | 5.351184 | 4.69847E-07 | 1.78876E-06 | 4.787812313 | Higher in PA |
| SPON2 | 0.21194 | 3.833097401 | 5.342954 | 4.87159E-07 | 1.84997E-06 | 4.752495295 | Higher in PA |
| TLR4\|LY96 | 0.12366 | 3.264989125 | 5.340229 | 4.93026E-07 | 1.86752E-06 | 4.74080983 | Higher in PA |
| PTPRS | 0.07920 | 4.651129854 | 5.335176 | 5.04092E-07 | 1.90463E-06 | 4.71914482 | Higher in PA |
| GLRX3 | -0.22447 | 4.033667521 | -5.33305 | 5.08808E-07 | 1.91762E-06 | 4.710056879 | Higher in NT |
| SPINK14 | 0.09481 | 3.034580535 | 5.331267 | 5.12817E-07 | 1.92788E-06 | 4.702397641 | Higher in PA |
| CD40LG | -0.10622 | 2.453432365 | -5.32451 | 5.28253E-07 | 1.98095E-06 | 4.67345679 | Higher in NT |
| PPBP | -0.38345 | 3.989962674 | -5.3126 | 5.56561E-07 | 2.0819E-06 | 4.622519298 | Higher in NT |
| DSC2 | 0.09202 | 3.002749194 | 5.311143 | 5.60126E-07 | 2.09002E-06 | 4.616288579 | Higher in PA |
| CD86 | 0.13093 | 3.498824462 | 5.308603 | 5.66393E-07 | 2.10816E-06 | 4.605432895 | Higher in PA |
| MCL1 | -0.11094 | 2.244122929 | -5.29892 | 5.90914E-07 | 2.19399E-06 | 4.564082037 | Higher in NT |
| THBS2 | 0.20275 | 3.794964334 | 5.277176 | 6.49822E-07 | 2.40675E-06 | 4.471386509 | Higher in PA |
| CR2 | 0.16072 | 3.776629506 | 5.27305 | 6.61627E-07 | 2.44444E-06 | 4.453826835 | Higher in PA |
| APP | -0.29494 | 4.469304519 | -5.27195 | 6.6481E-07 | 2.45016E-06 | 4.449146081 | Higher in NT |

| | | | | | | | |
|---|---|---|---|---|---|---|---|
| ICOSLG | -0.14250 | 3.691231749 | -5.26592 | 6.82518E-07 | 2.50926E-06 | 4.423510435 | Higher in NT |
| HSPB6 | 0.21063 | 3.899637881 | 5.264932 | 6.85476E-07 | 2.51397E-06 | 4.419293023 | Higher in PA |
| THSD1 | 0.09836 | 2.802650426 | 5.259129 | 7.03034E-07 | 2.57208E-06 | 4.394628439 | Higher in PA |
| TNFSF13B | 0.07372 | 2.964806145 | 5.254715 | 7.16683E-07 | 2.61563E-06 | 4.37587927 | Higher in PA |
| XXYLT1 | 0.09261 | 3.364248871 | 5.253331 | 7.21016E-07 | 2.62506E-06 | 4.370000967 | Higher in PA |
| PROC | 0.07597 | 4.875480606 | 5.236843 | 7.74641E-07 | 2.81347E-06 | 4.300057786 | Higher in PA |
| S100A9\|S100A8 | 0.20430 | 2.887716614 | 5.225373 | 8.14223E-07 | 2.95008E-06 | 4.251478086 | Higher in PA |
| TGFB1 | -0.16175 | 3.149299534 | -5.2236 | 8.20505E-07 | 2.96568E-06 | 4.243986421 | Higher in NT |
| INS | 0.14425 | 2.159459672 | 5.218385 | 8.39298E-07 | 3.02632E-06 | 4.221912824 | Higher in PA |
| PIK3R1\|PIK3CA | -0.14571 | 2.861347878 | -5.20403 | 8.9318E-07 | 3.21288E-06 | 4.161272193 | Higher in NT |
| TIMP2 | 0.07131 | 4.596651979 | 5.185801 | 9.66516E-07 | 3.46836E-06 | 4.08438169 | Higher in PA |
| PI3 | 0.17095 | 3.837815786 | 5.1638 | 1.06286E-06 | 3.805E-06 | 3.991812412 | Higher in PA |
| LAMC1\|LAMB1\|L | 0.09127 | 4.181041842 | 5.161262 | 1.07456E-06 | 3.83772E-06 | 3.981150853 | Higher in PA |
| FSTL1 | 0.07123 | 4.282078898 | 5.151154 | 1.12241E-06 | 3.99909E-06 | 3.938717779 | Higher in PA |
| GDF15 | 0.20292 | 4.218694925 | 5.143179 | 1.16163E-06 | 4.11996E-06 | 3.905271415 | Higher in PA |
| NR3C1 | -0.07597 | 2.557561455 | -5.14314 | 1.16183E-06 | 4.11996E-06 | 3.905104444 | Higher in NT |
| IL12B | 0.13398 | 2.408148065 | 5.139327 | 1.18104E-06 | 4.17822E-06 | 3.889131863 | Higher in PA |
| TCF4 | -0.23707 | 3.067928115 | -5.13395 | 1.20866E-06 | 4.26584E-06 | 3.866629756 | Higher in NT |
| MRC1 | 0.09992 | 3.636586127 | 5.127541 | 1.24244E-06 | 4.37479E-06 | 3.839786842 | Higher in PA |
| CCDC103 | -0.12891 | 2.41915087 | -5.11878 | 1.29006E-06 | 4.53183E-06 | 3.803169535 | Higher in NT |
| NRXN3.1 | 0.16099 | 3.398876124 | 5.115599 | 1.30782E-06 | 4.58348E-06 | 3.789860905 | Higher in PA |
| TNFRSF4 | 0.05939 | 2.82079537 | 5.114021 | 1.3167E-06 | 4.60386E-06 | 3.783270665 | Higher in PA |
| FASLG | 0.12438 | 2.777917204 | 5.106439 | 1.36023E-06 | 4.74498E-06 | 3.751614751 | Higher in PA |
| ASAH2 | 0.15201 | 3.23184861 | 5.099901 | 1.39888E-06 | 4.86849E-06 | 3.724343575 | Higher in PA |
| EGFR | 0.06772 | 4.130792112 | 5.093091 | 1.44028E-06 | 4.99555E-06 | 3.695959589 | Higher in PA |
| FUCA1 | 0.21892 | 3.782159626 | 5.092804 | 1.44205E-06 | 4.99555E-06 | 3.694762829 | Higher in PA |
| CTSZ | 0.09612 | 3.158777462 | 5.082873 | 1.50464E-06 | 5.20037E-06 | 3.653415221 | Higher in PA |
| IGF2R | 0.08525 | 3.791052719 | 5.08202 | 1.51014E-06 | 5.20738E-06 | 3.649865308 | Higher in PA |
| FMO3 | -0.09437 | 3.416521306 | -5.07567 | 1.55169E-06 | 5.33837E-06 | 3.623458958 | Higher in NT |
| SMPD1 | 0.10703 | 2.971439754 | 5.070685 | 1.58508E-06 | 5.44078E-06 | 3.602741312 | Higher in PA |
| FGF6 | -0.12113 | 2.657397006 | -5.05566 | 1.69002E-06 | 5.78775E-06 | 3.540376165 | Higher in NT |
| KLK3\|SERPINA3 | 0.12474 | 3.927139582 | 5.05465 | 1.69732E-06 | 5.79951E-06 | 3.53618491 | Higher in PA |
| CETP | 0.10343 | 2.804497048 | 5.051976 | 1.71678E-06 | 5.85265E-06 | 3.525098128 | Higher in PA |
| SNAP23 | -0.12870 | 3.612945006 | -5.05027 | 1.72932E-06 | 5.88203E-06 | 3.518019485 | Higher in NT |

| | | | | | | | |
|---|---|---|---|---|---|---|---|
| IL13RA1 | 0.12950 | 2.927853101 | 5.030146 | 1.88394E-06 | 6.38937E-06 | 3.434733417 | Higher in PA |
| CHD7 | 0.07254 | 2.577768835 | 5.029765 | 1.887E-06 | 6.38937E-06 | 3.433157766 | Higher in PA |
| CTSH | 0.20313 | 3.888432793 | 5.02117 | 1.95718E-06 | 6.61209E-06 | 3.39764875 | Higher in PA |
| COASY | -0.14773 | 3.172512128 | -5.00639 | 2.08383E-06 | 7.02415E-06 | 3.336688853 | Higher in NT |
| LPL | -0.09332 | 3.415295043 | -4.99662 | 2.17195E-06 | 7.30478E-06 | 3.296427202 | Higher in NT |
| CDC25B | -0.06312 | 3.589436574 | -4.9847 | 2.28429E-06 | 7.66541E-06 | 3.247415111 | Higher in NT |
| KL | 0.18977 | 3.386404213 | 4.983367 | 2.29724E-06 | 7.69164E-06 | 3.241924036 | Higher in PA |
| THRB | -0.20268 | 3.547657184 | -4.98042 | 2.32607E-06 | 7.77082E-06 | 3.229804142 | Higher in NT |
| PRTN3 | 0.20215 | 3.196510361 | 4.975103 | 2.37889E-06 | 7.92962E-06 | 3.207984734 | Higher in PA |
| DLD | -0.14981 | 3.730546545 | -4.96837 | 2.4475E-06 | 8.14026E-06 | 3.180354699 | Higher in NT |
| NRXN3 | 0.14369 | 3.565305213 | 4.954555 | 2.59436E-06 | 8.6096E-06 | 3.123743859 | Higher in PA |
| F8 | -0.17400 | 4.365240209 | -4.94618 | 2.68748E-06 | 8.89895E-06 | 3.089486952 | Higher in NT |
| FXYD2 | -0.09304 | 3.32853803 | -4.94104 | 2.74635E-06 | 9.07384E-06 | 3.068442582 | Higher in NT |
| EFNA5 | 0.12217 | 3.279616089 | 4.937043 | 2.79288E-06 | 9.2073E-06 | 3.052124515 | Higher in PA |
| CAMK2D | -0.16079 | 3.555670789 | -4.93297 | 2.84114E-06 | 9.34585E-06 | 3.035487484 | Higher in NT |
| HEXB | 0.10919 | 2.907335008 | 4.907447 | 3.16244E-06 | 1.038E-05 | 2.931457494 | Higher in PA |
| DLK1.2 | 0.04586 | 2.71833341 | 4.897482 | 3.29725E-06 | 1.07988E-05 | 2.890935489 | Higher in PA |
| TMPRSS6 | -0.19980 | 2.933275414 | -4.894 | 3.34572E-06 | 1.09337E-05 | 2.876771388 | Higher in NT |
| PARK7 | -0.10975 | 2.820345406 | -4.8931 | 3.35826E-06 | 1.09509E-05 | 2.873137738 | Higher in NT |
| SERPINE1 | -0.23690 | 3.666131096 | -4.89005 | 3.40137E-06 | 1.10674E-05 | 2.860759777 | Higher in NT |
| BRD4 | -0.08697 | 2.806868695 | -4.88408 | 3.48742E-06 | 1.13228E-05 | 2.836512312 | Higher in NT |
| MLX | -0.14064 | 3.057825772 | -4.87465 | 3.62747E-06 | 1.17521E-05 | 2.798302628 | Higher in NT |
| ACAT2 | -0.28735 | 3.731983177 | -4.86966 | 3.70389E-06 | 1.19738E-05 | 2.778073196 | Higher in NT |
| ZC3H12A | -0.13751 | 3.031794535 | -4.86627 | 3.75664E-06 | 1.21182E-05 | 2.764353745 | Higher in NT |
| MPO | 0.14333 | 4.008084871 | 4.85869 | 3.87725E-06 | 1.24804E-05 | 2.733692714 | Higher in PA |
| TMED2 | -0.04516 | 2.726553369 | -4.84699 | 4.07078E-06 | 1.30753E-05 | 2.686443546 | Higher in NT |
| ACP2 | 0.23214 | 2.515889979 | 4.842389 | 4.14953E-06 | 1.32998E-05 | 2.667860797 | Higher in PA |
| SFRP4 | 0.11457 | 3.877745223 | 4.836172 | 4.2582E-06 | 1.3619E-05 | 2.642787877 | Higher in PA |
| PHYH | -0.06530 | 3.318078074 | -4.8311 | 4.34879E-06 | 1.38791E-05 | 2.622371656 | Higher in NT |
| TGFBR1 | -0.17476 | 2.627840372 | -4.82897 | 4.38752E-06 | 1.3973E-05 | 2.613774461 | Higher in NT |
| IL2RB | 0.12731 | 3.271256858 | 4.817194 | 4.60724E-06 | 1.46417E-05 | 2.566392873 | Higher in PA |
| CCL14 | 0.16474 | 3.663419758 | 4.811205 | 4.72306E-06 | 1.4978E-05 | 2.542323289 | Higher in PA |
| FGF7 | 0.08237 | 2.472273799 | 4.798655 | 4.97497E-06 | 1.57436E-05 | 2.491951646 | Higher in PA |
| GLUL | -0.11537 | 3.57094547 | -4.789 | 5.17754E-06 | 1.63501E-05 | 2.45326939 | Higher in NT |

| | | | | | | | |
|---|---|---|---|---|---|---|---|
| HIF1A | -0.08795 | 2.540147779 | -4.7839 | 5.2878E-06 | 1.66632E-05 | 2.432848315 | Higher in NT |
| PEBP1 | -0.21434 | 3.613848606 | -4.78044 | 5.36393E-06 | 1.68677E-05 | 2.418995513 | Higher in NT |
| CDKN1A | -0.09123 | 2.947737708 | -4.76764 | 5.65476E-06 | 1.77451E-05 | 2.367836122 | Higher in NT |
| NDUFS4 | -0.26205 | 3.439891956 | -4.7385 | 6.37491E-06 | 1.99632E-05 | 2.251725432 | Higher in NT |
| CYTH1 | -0.08188 | 2.887457298 | -4.7364 | 6.43026E-06 | 2.00946E-05 | 2.243353062 | Higher in NT |
| OLR1 | 0.12833 | 3.05350649 | 4.729465 | 6.61579E-06 | 2.06314E-05 | 2.215810253 | Higher in PA |
| SH3PXD2B | -0.26202 | 3.035802219 | -4.71118 | 7.13056E-06 | 2.21905E-05 | 2.143266795 | Higher in NT |
| SAA1 | 0.32709 | 3.932254877 | 4.704109 | 7.33974E-06 | 2.27942E-05 | 2.115279412 | Higher in PA |
| C4B\|C4A | 0.06399 | 5.144095982 | 4.695691 | 7.59663E-06 | 2.35433E-05 | 2.081984836 | Higher in PA |
| BMP6 | 0.08732 | 3.136786538 | 4.693239 | 7.67308E-06 | 2.37312E-05 | 2.072295631 | Higher in PA |
| EPHA3 | 0.05160 | 2.123070006 | 4.675548 | 8.2472E-06 | 2.54543E-05 | 2.002477965 | Higher in PA |
| PCNA | 0.18708 | 2.701978597 | 4.674698 | 8.27579E-06 | 2.54901E-05 | 1.999129572 | Higher in PA |
| TAB1\|MAP3K7 | -0.09392 | 2.86810557 | -4.66283 | 8.68552E-06 | 2.66889E-05 | 1.952384827 | Higher in NT |
| PKP2 | -0.16263 | 2.827056875 | -4.6624 | 8.70059E-06 | 2.66889E-05 | 1.950708089 | Higher in NT |
| FOXRED1 | -0.11213 | 2.778980427 | -4.64851 | 9.20596E-06 | 2.81815E-05 | 1.89610249 | Higher in NT |
| THRA | -0.08372 | 2.923006062 | -4.64471 | 9.34901E-06 | 2.85611E-05 | 1.88119228 | Higher in NT |
| SAR1A | -0.08427 | 2.823952253 | -4.6441 | 9.37222E-06 | 2.85738E-05 | 1.878794094 | Higher in NT |
| GRN | 0.06256 | 4.115056176 | 4.618743 | 1.0386E-05 | 3.16003E-05 | 1.779502782 | Higher in PA |
| ENG | 0.16273 | 3.278466094 | 4.61721 | 1.04506E-05 | 3.17325E-05 | 1.773509653 | Higher in PA |
| FLT4 | 0.14407 | 3.799050365 | 4.607395 | 1.08733E-05 | 3.29036E-05 | 1.73518089 | Higher in PA |
| IFNG | -0.11336 | 2.573579902 | -4.60724 | 1.08801E-05 | 3.29036E-05 | 1.73457905 | Higher in NT |
| GUSB | -0.21688 | 3.527326818 | -4.59804 | 1.12916E-05 | 3.40792E-05 | 1.698707931 | Higher in NT |
| SURF1 | 0.13277 | 3.820570697 | 4.569513 | 1.26658E-05 | 3.81501E-05 | 1.587762489 | Higher in PA |
| GCLM | -0.23377 | 3.668013113 | -4.5644 | 1.29286E-05 | 3.88635E-05 | 1.567933198 | Higher in NT |
| PRL | 0.23102 | 3.767630242 | 4.563471 | 1.2977E-05 | 3.8931E-05 | 1.564323001 | Higher in PA |
| AOC3 | 0.08961 | 4.484363522 | 4.543811 | 1.40414E-05 | 4.204E-05 | 1.488213364 | Higher in PA |
| ISCU | -0.10462 | 2.707808235 | -4.51144 | 1.598E-05 | 4.77489E-05 | 1.363400794 | Higher in NT |
| MYL3 | 0.21165 | 2.921383415 | 4.509004 | 1.6136E-05 | 4.81194E-05 | 1.354023604 | Higher in PA |
| INSR | 0.15648 | 3.77985332 | 4.499936 | 1.67295E-05 | 4.97903E-05 | 1.319178311 | Higher in PA |
| IL18BP | 0.16080 | 3.534595591 | 4.493396 | 1.71706E-05 | 5.10017E-05 | 1.294078074 | Higher in PA |
| NTRK2 | 0.16383 | 3.858752235 | 4.486388 | 1.76556E-05 | 5.23387E-05 | 1.267213154 | Higher in PA |
| FABP1 | 0.10158 | 3.024414488 | 4.466044 | 1.91398E-05 | 5.66266E-05 | 1.189384457 | Higher in PA |
| EPHA2 | 0.12120 | 3.003904124 | 4.464 | 1.92954E-05 | 5.69747E-05 | 1.181577944 | Higher in PA |
| SCO1 | -0.06788 | 3.161176859 | -4.44716 | 2.06249E-05 | 6.07513E-05 | 1.117353236 | Higher in NT |

| | | | | | | | |
|---|---|---|---|---|---|---|---|
| ENO3 | -0.19978 | 3.174554885 | -4.44678 | 2.06554E-05 | 6.07513E-05 | 1.11592913 | Higher in NT |
| TFRC.1 | 0.10075 | 3.711580876 | 4.445744 | 2.07405E-05 | 6.08821E-05 | 1.111969422 | Higher in PA |
| HMOX1 | 0.08827 | 3.378110098 | 4.441502 | 2.1091E-05 | 6.179E-05 | 1.095821594 | Higher in PA |
| IL23R | -0.06162 | 3.184312439 | -4.4266 | 2.23675E-05 | 6.54021E-05 | 1.039203541 | Higher in NT |
| F9 | 0.04429 | 4.319617435 | 4.423232 | 2.26666E-05 | 6.61477E-05 | 1.026406894 | Higher in PA |
| FDX2 | -0.13131 | 2.18732116 | -4.32699 | 3.30289E-05 | 9.62008E-05 | 0.664066511 | Higher in NT |
| NEURL1 | 0.04580 | 3.018171575 | 4.324871 | 3.33017E-05 | 9.6632E-05 | 0.65615926 | Higher in PA |
| SLAMF1 | 0.09496 | 3.062603096 | 4.324839 | 3.33058E-05 | 9.6632E-05 | 0.656039743 | Higher in PA |
| B4GALT7 | -0.16448 | 3.49256298 | -4.32323 | 3.35149E-05 | 9.70508E-05 | 0.65002394 | Higher in NT |
| RLN3 | -0.06474 | 2.7499593 | -4.29334 | 3.76286E-05 | 0.000108753 | 0.538743538 | Higher in NT |
| IAPP | 0.14355 | 3.303156416 | 4.269946 | 4.11846E-05 | 0.000118802 | 0.451997552 | Higher in PA |
| OXSR1 | -0.21934 | 3.337115684 | -4.2668 | 4.16871E-05 | 0.00012002 | 0.440350147 | Higher in NT |
| LGALS9 | 0.06316 | 2.962375775 | 4.261169 | 4.26E-05 | 0.000122414 | 0.419545549 | Higher in PA |
| KIF23 | -0.12941 | 2.745675875 | -4.2526 | 4.40276E-05 | 0.000126274 | 0.387897009 | Higher in NT |
| EFNB2 | 0.15400 | 3.673310551 | 4.247066 | 4.49728E-05 | 0.000128739 | 0.367504129 | Higher in PA |
| NAGLU | 0.16960 | 3.661411655 | 4.242087 | 4.58404E-05 | 0.000130972 | 0.349162566 | Higher in PA |
| DCXR | -0.18569 | 3.193505615 | -4.2346 | 4.71762E-05 | 0.000134533 | 0.32159252 | Higher in NT |
| SERPINA7 | 0.04514 | 4.850583245 | 4.226088 | 4.87388E-05 | 0.000138725 | 0.290321889 | Higher in PA |
| CRP | 0.26642 | 4.380209705 | 4.221843 | 4.95367E-05 | 0.000140729 | 0.274739429 | Higher in PA |
| LYVE1 | 0.09167 | 4.638029538 | 4.214297 | 5.09865E-05 | 0.000143818 | 0.24706463 | Higher in PA |
| TAC1 | 0.08183 | 2.428782278 | 4.214237 | 5.09981E-05 | 0.000143818 | 0.246844813 | Higher in PA |
| HK2 | 0.15775 | 2.799217179 | 4.214053 | 5.10341E-05 | 0.000143818 | 0.24616813 | Higher in PA |
| IL1R1 | 0.16608 | 3.509769931 | 4.213935 | 5.1057E-05 | 0.000143818 | 0.245737755 | Higher in PA |
| BECN1 | 0.05421 | 2.300869676 | 4.213699 | 5.11032E-05 | 0.000143818 | 0.244870036 | Higher in PA |
| THY1 | 0.11108 | 2.748207459 | 4.20561 | 5.2706E-05 | 0.000148051 | 0.215246911 | Higher in PA |
| CASP7 | -0.07703 | 2.573214449 | -4.2042 | 5.29912E-05 | 0.000148351 | 0.210071827 | Higher in NT |
| SLITRK6 | 0.07074 | 2.666313476 | 4.204099 | 5.30107E-05 | 0.000148351 | 0.209718161 | Higher in PA |
| FAS | 0.08782 | 3.472541148 | 4.201839 | 5.34695E-05 | 0.000149356 | 0.201452947 | Higher in PA |
| NPPA | 0.11060 | 3.100096423 | 4.190752 | 5.57767E-05 | 0.000155511 | 0.160941769 | Higher in PA |
| FBP2 | 0.08898 | 2.87593563 | 4.157358 | 6.33172E-05 | 0.000176207 | 0.039401632 | Higher in PA |
| PPARA | -0.16760 | 3.125073979 | -4.15087 | 6.48913E-05 | 0.000180254 | 0.015873244 | Higher in NT |
| SSPN | -0.13677 | 3.633710215 | -4.14843 | 6.54924E-05 | 0.000181587 | 0.007040671 | Higher in NT |
| PDK2 | -0.10512 | 2.702472777 | -4.14594 | 6.61132E-05 | 0.00018297 | -0.001996942 | Higher in NT |
| PPT1 | -0.12241 | 2.969591478 | -4.12564 | 7.13777E-05 | 0.000197176 | -0.075374475 | Higher in NT |

| | | | | | | | |
|---|---|---|---|---|---|---|---|
| HMOX2 | -0.11048 | 2.610130993 | -4.12273 | 7.21654E-05 | 0.000198796 | -0.085883167 | Higher in NT |
| PLAUR | 0.12682 | 3.600031139 | 4.122495 | 7.22291E-05 | 0.000198796 | -0.086727824 | Higher in PA |
| S100A4 | -0.09027 | 2.869492249 | -4.11419 | 7.45227E-05 | 0.000204733 | -0.116654639 | Higher in NT |
| FGB | -0.08691 | 3.175865896 | -4.10371 | 7.75177E-05 | 0.000212571 | -0.154369318 | Higher in NT |
| FGFR3 | 0.16242 | 3.819094434 | 4.101992 | 7.80192E-05 | 0.000213556 | -0.160539954 | Higher in PA |
| AGRP | 0.12815 | 3.007882601 | 4.100121 | 7.85691E-05 | 0.00021467 | -0.167262338 | Higher in PA |
| HMGB2 | 0.11318 | 2.688034461 | 4.098967 | 7.89101E-05 | 0.000215209 | -0.171406541 | Higher in PA |
| TCN2 | 0.08530 | 3.838131611 | 4.087114 | 8.24964E-05 | 0.000224582 | -0.21393162 | Higher in PA |
| GOSR2 | -0.18941 | 2.035518507 | -4.08659 | 8.26586E-05 | 0.000224616 | -0.21581008 | Higher in NT |
| SEMA4A | 0.07094 | 3.942820292 | 4.08266 | 8.38839E-05 | 0.000227533 | -0.22988757 | Higher in PA |
| CXCL1 | -0.18220 | 3.372441382 | -4.0598 | 9.13646E-05 | 0.000247377 | -0.311580991 | Higher in NT |
| INPP5D | -0.13219 | 3.119548437 | -4.05643 | 9.25179E-05 | 0.000250048 | -0.323573448 | Higher in NT |
| CCL3 | 0.07439 | 2.796509072 | 4.053763 | 9.34438E-05 | 0.000252097 | -0.333092503 | Higher in PA |
| HIP1R | 0.03598 | 2.561115757 | 4.047136 | 9.57785E-05 | 0.000257931 | -0.356681377 | Higher in PA |
| C2 | 0.04621 | 3.898247653 | 4.042712 | 9.73683E-05 | 0.000261743 | -0.372415695 | Higher in PA |
| CD3E | 0.08904 | 2.952397818 | 4.039108 | 9.8682E-05 | 0.0002648 | -0.385223345 | Higher in PA |
| TNFAIP6 | 0.11429 | 3.056224289 | 4.024049 | 0.000104357 | 0.000279527 | -0.438644283 | Higher in PA |
| NHEJ1 | -0.10802 | 2.536898015 | -4.01595 | 0.000107537 | 0.000287532 | -0.467319342 | Higher in NT |
| TGM2 | -0.28021 | 3.292119076 | -4.00273 | 0.000112927 | 0.000301407 | -0.51402187 | Higher in NT |
| NPTN | 0.13169 | 3.549248696 | 3.99874 | 0.000114604 | 0.000305339 | -0.528094597 | Higher in PA |
| NT5E | 0.14563 | 3.894366009 | 3.993855 | 0.00011669 | 0.000310345 | -0.545311602 | Higher in PA |
| GSTP1 | -0.14615 | 3.517345069 | -3.98825 | 0.000119127 | 0.000316267 | -0.565046404 | Higher in NT |
| TNFRSF11B | 0.14994 | 3.702522327 | 3.986298 | 0.000119987 | 0.000317988 | -0.571912457 | Higher in PA |
| QPCT | 0.12026 | 2.949301137 | 3.978246 | 0.000123599 | 0.000326982 | -0.600214564 | Higher in PA |
| SULT1E1 | -0.04977 | 3.238146255 | -3.97441 | 0.000125357 | 0.000331048 | -0.613689362 | Higher in NT |
| ENTPD6 | -0.06892 | 2.961090061 | -3.97208 | 0.000126433 | 0.000333303 | -0.621843554 | Higher in NT |
| C1orf185 | 0.05128 | 2.185584277 | 3.970762 | 0.000127049 | 0.000334339 | -0.626481018 | Higher in PA |
| SOD3 | 0.10223 | 3.665283537 | 3.957282 | 0.000133497 | 0.000350692 | -0.673702859 | Higher in PA |
| LEP | 0.26768 | 4.291235668 | 3.955413 | 0.000134415 | 0.000352486 | -0.680237677 | Higher in PA |
| GLA | -0.13927 | 3.329790569 | -3.95348 | 0.000135371 | 0.000354375 | -0.687000154 | Higher in NT |
| TNFRSF21 | 0.14866 | 3.563197664 | 3.946514 | 0.00013887 | 0.000362901 | -0.711333062 | Higher in PA |
| SEMA3C | 0.05816 | 2.895903715 | 3.945519 | 0.000139377 | 0.000363591 | -0.714805349 | Higher in PA |
| CD320 | -0.21113 | 3.050893022 | -3.93611 | 0.000144259 | 0.000375673 | -0.747626857 | Higher in NT |
| AHCY | -0.12763 | 3.707148856 | -3.93394 | 0.000145405 | 0.000378004 | -0.755174361 | Higher in NT |

| | | | | | | | |
|---|---|---|---|---|---|---|---|
| CBFB | -0.06052 | 3.083252403 | -3.92227 | 0.000151732 | 0.000393768 | -0.795763943 | Higher in NT |
| NCF1 | 0.18473 | 2.862660656 | 3.921242 | 0.000152304 | 0.000394569 | -0.79934848 | Higher in PA |
| PDHB | -0.13666 | 3.222143925 | -3.91876 | 0.000153689 | 0.000397473 | -0.807978406 | Higher in NT |
| EIF2AK4 | -0.20803 | 2.027400034 | -3.90871 | 0.000159419 | 0.00041158 | -0.842847076 | Higher in NT |
| CXCL11 | -0.19271 | 3.238026768 | -3.90187 | 0.000163431 | 0.000421214 | -0.866523354 | Higher in NT |
| RGS7 | -0.12462 | 2.678235865 | -3.89943 | 0.000164884 | 0.000424231 | -0.874954939 | Higher in NT |
| SELPLG | 0.11803 | 3.290283458 | 3.886289 | 0.00017294 | 0.000444195 | -0.920372272 | Higher in PA |
| FLT1 | 0.15925 | 2.666639276 | 3.884229 | 0.000174236 | 0.000446759 | -0.927480174 | Higher in PA |
| NAGPA | 0.12470 | 3.700336581 | 3.882789 | 0.000175147 | 0.00044833 | -0.932447813 | Higher in PA |
| ADAMTSL1 | 0.13618 | 3.808952352 | 3.870956 | 0.00018281 | 0.000467147 | -0.973203185 | Higher in PA |
| ESS2 | -0.05461 | 2.682121206 | -3.86965 | 0.000183677 | 0.000468565 | -0.977706853 | Higher in NT |
| IL1RL1 | 0.15588 | 3.43040792 | 3.860932 | 0.000189551 | 0.000482727 | -1.007656687 | Higher in PA |
| INA | 0.06489 | 2.720925572 | 3.859305 | 0.000190667 | 0.000484746 | -1.013241461 | Higher in PA |
| GSTT2 | -0.09498 | 3.100505782 | -3.85532 | 0.000193424 | 0.000490923 | -1.026897043 | Higher in NT |
| PYCR1 | 0.09920 | 2.564484355 | 3.849012 | 0.000197874 | 0.00050137 | -1.048533955 | Higher in PA |
| MALT1 | 0.06575 | 2.443240529 | 3.84825 | 0.000198418 | 0.0005019 | -1.051144695 | Higher in PA |
| UNC5B | 0.11711 | 3.687373146 | 3.847182 | 0.000199182 | 0.000502986 | -1.05480218 | Higher in PA |
| APLP2 | -0.16667 | 3.167545302 | -3.8357 | 0.000207581 | 0.000523313 | -1.094072859 | Higher in NT |
| CA4 | 0.08151 | 2.938633539 | 3.830186 | 0.000211733 | 0.000532884 | -1.11289992 | Higher in PA |
| CASP8 | 0.05080 | 2.751110477 | 3.823484 | 0.000216885 | 0.000544938 | -1.135755171 | Higher in PA |
| ACHE | 0.13618 | 3.26644128 | 3.811424 | 0.00022646 | 0.000568044 | -1.176807003 | Higher in PA |
| MSTN | 0.08993 | 3.581789023 | 3.808491 | 0.000228849 | 0.000573077 | -1.18677525 | Higher in PA |
| VEGFA | 0.05906 | 3.775778239 | 3.796826 | 0.000238587 | 0.000596467 | -1.226360124 | Higher in PA |
| JAM2 | -0.04881 | 3.673906854 | -3.79445 | 0.000240622 | 0.000600555 | -1.234428517 | Higher in NT |
| FBLN5 | -0.13996 | 3.83164365 | -3.78601 | 0.000247966 | 0.000617855 | -1.262974761 | Higher in NT |
| ADM | 0.06076 | 2.694305564 | 3.776925 | 0.000256118 | 0.000637109 | -1.293682691 | Higher in PA |
| SGSH | 0.13309 | 3.829658779 | 3.757201 | 0.000274695 | 0.00068219 | -1.360133039 | Higher in PA |
| SAR1B | 0.07113 | 3.511751303 | 3.748429 | 0.000283363 | 0.000702535 | -1.389600196 | Higher in PA |
| IDS | 0.07802 | 3.345691433 | 3.747969 | 0.000283824 | 0.000702535 | -1.39114444 | Higher in PA |
| CCL18 | 0.08681 | 4.830192342 | 3.734393 | 0.000297773 | 0.000735849 | -1.436639694 | Higher in PA |
| PROCR | 0.10035 | 3.467416922 | 3.733495 | 0.000298719 | 0.000736971 | -1.439644643 | Higher in PA |
| TRAF1 | -0.07386 | 3.270244319 | -3.72879 | 0.000303721 | 0.000747256 | -1.455387066 | Higher in NT |
| DNM1L | 0.15363 | 3.685990474 | 3.728635 | 0.000303884 | 0.000747256 | -1.45589714 | Higher in PA |
| DTNBP1 | 0.04254 | 2.252444091 | 3.725931 | 0.000306795 | 0.000753179 | -1.464932657 | Higher in PA |

| | | | | | | | |
|---|---|---|---|---|---|---|---|
| PLAT | -0.20256 | 3.714594695 | -3.72493 | 0.000307877 | 0.0007546 | -1.468269262 | Higher in NT |
| FTO | 0.07592 | 3.1809653 | 3.723612 | 0.000309311 | 0.000756879 | -1.472674763 | Higher in PA |
| PRSS27 | 0.10443 | 3.155904708 | 3.721576 | 0.000311538 | 0.00076095 | -1.479473189 | Higher in PA |
| NID1 | -0.12731 | 3.299848194 | -3.72116 | 0.00031199 | 0.00076095 | -1.480846476 | Higher in NT |
| AMN | 0.05776 | 2.217077829 | 3.69871 | 0.000337605 | 0.000822091 | -1.555603744 | Higher in PA |
| GCKR | 0.05727 | 2.745311135 | 3.693865 | 0.000343388 | 0.000834818 | -1.57168882 | Higher in PA |
| TFF3 | 0.14388 | 3.807851834 | 3.686509 | 0.000352348 | 0.000855213 | -1.596076531 | Higher in PA |
| PROC.1 | 0.05467 | 4.345855934 | 3.674972 | 0.000366848 | 0.000888968 | -1.63424994 | Higher in PA |
| ETFA | -0.05502 | 3.285112678 | -3.66667 | 0.000377635 | 0.000913633 | -1.66167564 | Higher in NT |
| SERAC1 | -0.07282 | 3.118973701 | -3.65963 | 0.000387011 | 0.000934808 | -1.684877154 | Higher in NT |
| PRDX6 | -0.12106 | 4.648673337 | -3.65847 | 0.00038857 | 0.000937067 | -1.688682141 | Higher in NT |
| FABP4 | 0.16497 | 4.24746947 | 3.656679 | 0.000391003 | 0.000941419 | -1.694584509 | Higher in PA |
| PLEKHA1 | 0.04277 | 2.456987568 | 3.655348 | 0.000392817 | 0.000944272 | -1.698964464 | Higher in PA |
| NDUFB11 | -0.08499 | 3.212224584 | -3.65372 | 0.000395054 | 0.00094813 | -1.704335269 | Higher in NT |
| JUN | -0.06700 | 3.088455408 | -3.63099 | 0.000427479 | 0.001023706 | -1.77891771 | Higher in NT |
| BCL2 | -0.11053 | 2.98815907 | -3.6307 | 0.000427909 | 0.001023706 | -1.779867528 | Higher in NT |
| TIMP4 | 0.08996 | 3.476650929 | 3.627785 | 0.000432248 | 0.001032439 | -1.789402391 | Higher in PA |
| PFKM | -0.14257 | 3.65385623 | -3.60085 | 0.000474381 | 0.001131274 | -1.877267629 | Higher in NT |
| HMG20A | -0.05487 | 2.639106467 | -3.58656 | 0.000498267 | 0.00118635 | -1.923644757 | Higher in NT |
| ADAMTSL2 | 0.06555 | 3.038543404 | 3.58579 | 0.000499591 | 0.001187618 | -1.926149925 | Higher in PA |
| CEL | -0.16202 | 3.144476599 | -3.58032 | 0.000509067 | 0.001208229 | -1.943881984 | Higher in NT |
| CXCL10 | -0.09813 | 2.954922683 | -3.57844 | 0.000512351 | 0.001214101 | -1.949948728 | Higher in NT |
| GH1 | 0.05901 | 2.577924457 | 3.576785 | 0.000515269 | 0.00121909 | -1.95530774 | Higher in PA |
| MVK | -0.11639 | 3.474054606 | -3.57431 | 0.000519659 | 0.001227542 | -1.963313198 | Higher in NT |
| WNK1 | -0.14832 | 2.752194374 | -3.54916 | 0.000566316 | 0.00133565 | -2.044398275 | Higher in NT |
| IKBKG | -0.19492 | 2.906246714 | -3.51851 | 0.000628519 | 0.00148003 | -2.142592289 | Higher in NT |
| FGF8 | -0.06044 | 2.29425189 | -3.51768 | 0.000630299 | 0.001481895 | -2.14525552 | Higher in NT |
| PTHLH | -0.04596 | 2.948549336 | -3.49434 | 0.000682063 | 0.001601086 | -2.219552795 | Higher in NT |
| TCAP | 0.04422 | 3.061780409 | 3.492651 | 0.000685956 | 0.00160771 | -2.224909162 | Higher in PA |
| BIRC3 | -0.08225 | 2.412989643 | -3.46715 | 0.000747422 | 0.001749037 | -2.305619725 | Higher in NT |
| SCN3B | 0.09840 | 2.497325978 | 3.466124 | 0.000749992 | 0.001752319 | -2.30884738 | Higher in PA |
| IDUA | -0.12667 | 3.170877414 | -3.45925 | 0.000767474 | 0.001790374 | -2.330505005 | Higher in NT |
| GPX3 | -0.05052 | 3.548500452 | -3.45611 | 0.000775586 | 0.001806489 | -2.340386134 | Higher in NT |
| UMOD | 0.14919 | 3.711122957 | 3.451041 | 0.000788869 | 0.001834579 | -2.356342774 | Higher in PA |

| | | | | | | | |
|---|---|---|---|---|---|---|---|
| CTNNB1 | -0.19810 | 2.938185588 | -3.4465 | 0.000800949 | 0.001858687 | -2.370620857 | Higher in NT |
| ITGB1\|ITGA1 | -0.12563 | 3.956986363 | -3.44621 | 0.000801714 | 0.001858687 | -2.37151677 | Higher in NT |
| PMM2 | -0.23677 | 3.861426916 | -3.43869 | 0.000822114 | 0.001903042 | -2.395119104 | Higher in NT |
| GALE | -0.08252 | 2.834486665 | -3.43333 | 0.000836931 | 0.001934356 | -2.411893088 | Higher in NT |
| GPHN | 0.06229 | 2.366095294 | 3.424287 | 0.000862529 | 0.001990452 | -2.440177364 | Higher in PA |
| ME1 | -0.12048 | 3.161568088 | -3.42093 | 0.000872227 | 0.002009739 | -2.450671174 | Higher in NT |
| CD36 | -0.14141 | 4.135876918 | -3.41858 | 0.000879067 | 0.002022394 | -2.458003338 | Higher in NT |
| SPR | 0.08011 | 2.619736202 | 3.403992 | 0.000922667 | 0.002119449 | -2.503419971 | Higher in PA |
| ADGRE2 | 0.10832 | 3.081486511 | 3.395847 | 0.000947891 | 0.002174063 | -2.528715389 | Higher in PA |
| PKD2 | -0.08631 | 2.911677073 | -3.38195 | 0.000992417 | 0.00227271 | -2.571748292 | Higher in NT |
| JAG1.1 | 0.07108 | 2.642831568 | 3.37658 | 0.00101016 | 0.002309817 | -2.588355236 | Higher in PA |
| SMOC2 | 0.06786 | 3.062518366 | 3.369596 | 0.001033667 | 0.00235997 | -2.609907829 | Higher in PA |
| IGF1R | 0.10489 | 3.428659177 | 3.368079 | 0.001038837 | 0.00236817 | -2.614581978 | Higher in PA |
| PAM | 0.05040 | 3.445275 | 3.359686 | 0.001067899 | 0.002430726 | -2.640423588 | Higher in PA |
| THBS1 | -0.23703 | 3.380798653 | -3.35454 | 0.001086097 | 0.002468401 | -2.656245263 | Higher in NT |
| BMP4 | 0.09825 | 2.926918346 | 3.348441 | 0.001108029 | 0.002514438 | -2.674961532 | Higher in PA |
| NTN1 | 0.09903 | 2.657150113 | 3.347932 | 0.00110988 | 0.002514834 | -2.676524084 | Higher in PA |
| LBP | 0.08920 | 4.635553586 | 3.342442 | 0.001130013 | 0.002556592 | -2.693349901 | Higher in PA |
| EWSR1 | 0.07307 | 3.334848003 | 3.340476 | 0.001137303 | 0.002569209 | -2.699367571 | Higher in PA |
| MGP | -0.08113 | 3.833542991 | -3.33469 | 0.001159019 | 0.002614328 | -2.717064524 | Higher in NT |
| MAPT | -0.06037 | 2.310016878 | -3.32773 | 0.001185673 | 0.002670435 | -2.738333477 | Higher in NT |
| TTN | 0.06244 | 2.621688454 | 3.32141 | 0.00121035 | 0.002721927 | -2.757597516 | Higher in PA |
| PTH | 0.06428 | 2.509351904 | 3.3155 | 0.001233873 | 0.002770672 | -2.775594003 | Higher in PA |
| MAT1A | 0.13161 | 2.533504975 | 3.309055 | 0.001260007 | 0.002825128 | -2.795186149 | Higher in PA |
| ASPA | 0.06377 | 2.853317668 | 3.306155 | 0.001271938 | 0.002847623 | -2.803994075 | Higher in PA |
| HPX | 0.14324 | 3.363484116 | 3.303032 | 0.001284901 | 0.002872358 | -2.813469706 | Higher in PA |
| NFE2L2 | -0.11709 | 2.747408722 | -3.29169 | 0.001333039 | 0.002975534 | -2.847829342 | Higher in NT |
| COL3A1 | 0.05670 | 3.921252747 | 3.288074 | 0.001348724 | 0.003006072 | -2.858754098 | Higher in PA |
| CTNNA3 | 0.11532 | 2.463647336 | 3.285684 | 0.001359192 | 0.003024907 | -2.865973423 | Higher in PA |
| ZAP70 | 0.09991 | 2.850887637 | 3.281694 | 0.001376836 | 0.003059635 | -2.87801629 | Higher in PA |
| GCK | -0.09844 | 3.357851304 | -3.28046 | 0.001382345 | 0.003067334 | -2.881744442 | Higher in NT |
| PPP2R3A | 0.07427 | 2.767895557 | 3.279586 | 0.001386249 | 0.003071452 | -2.884377007 | Higher in PA |
| MMP12 | 0.10230 | 2.883044697 | 3.261631 | 0.001468855 | 0.00324968 | -2.938393186 | Higher in PA |
| PPARG | -0.07083 | 3.053258892 | -3.25803 | 0.001485987 | 0.003282739 | -2.949209646 | Higher in NT |

| | | | | | | | |
|---|---|---|---|---|---|---|---|
| GSR | 0.04439 | 4.353787601 | 3.253932 | 0.00150567 | 0.00332133 | -2.961482184 | Higher in PA |
| TNXB | 0.07539 | 3.956169039 | 3.251472 | 0.001517614 | 0.003342762 | -2.968850742 | Higher in PA |
| NEGR1 | 0.04905 | 2.703695365 | 3.243193 | 0.001558457 | 0.00342365 | -2.993610623 | Higher in PA |
| ACE2 | -0.05969 | 2.497208454 | -3.24309 | 0.001558991 | 0.00342365 | -2.993930114 | Higher in NT |
| AOC1 | 0.19990 | 3.37650238 | 3.242648 | 0.001561185 | 0.00342365 | -2.995240566 | Higher in PA |
| PENK | 0.13865 | 3.763418277 | 3.23862 | 0.001581458 | 0.003463046 | -3.007266367 | Higher in PA |
| MYBPC3 | 0.06492 | 2.322770239 | 3.221724 | 0.001669206 | 0.003649868 | -3.057577641 | Higher in PA |
| ICAM1 | 0.10886 | 3.236713646 | 3.216109 | 0.001699362 | 0.003710397 | -3.074250912 | Higher in PA |
| TIMD4 | 0.08426 | 2.711924158 | 3.215173 | 0.001704434 | 0.003716063 | -3.077026103 | Higher in PA |
| PDGFRB | 0.10082 | 3.36700851 | 3.202508 | 0.001774518 | 0.003863247 | -3.114534769 | Higher in PA |
| A2M | 0.06391 | 4.087687592 | 3.190513 | 0.001843356 | 0.004007297 | -3.149943367 | Higher in PA |
| TYRO3 | 0.06778 | 2.886660834 | 3.185343 | 0.001873787 | 0.004067554 | -3.165170643 | Higher in PA |
| NCF4 | -0.13006 | 2.900450168 | -3.18455 | 0.001878483 | 0.004071856 | -3.167498296 | Higher in NT |
| TPM1 | -0.06516 | 3.013884905 | -3.181 | 0.001899722 | 0.004111953 | -3.177952288 | Higher in NT |
| IL19 | 0.07105 | 3.757991911 | 3.16988 | 0.001967617 | 0.004252775 | -3.210592783 | Higher in PA |
| NQO1 | 0.10774 | 3.033358531 | 3.166708 | 0.001987395 | 0.004289343 | -3.219886534 | Higher in PA |
| CBLIF | 0.13495 | 3.186384111 | 3.14843 | 0.002105025 | 0.004533704 | -3.273294057 | Higher in PA |
| AGGF1 | 0.06340 | 2.136042807 | 3.147816 | 0.002109087 | 0.004533704 | -3.27508381 | Higher in PA |
| CCL2 | 0.04453 | 2.455250076 | 3.147726 | 0.002109684 | 0.004533704 | -3.275346493 | Higher in PA |
| IL2RA | -0.14429 | 2.885446769 | -3.14537 | 0.002125315 | 0.004560762 | -3.282199528 | Higher in NT |
| GAS6 | 0.04767 | 3.084766126 | 3.129682 | 0.002232375 | 0.00478366 | -3.327805158 | Higher in PA |
| SELPLG.1 | -0.10772 | 2.786085905 | -3.12915 | 0.002236086 | 0.004784777 | -3.329345923 | Higher in NT |
| NOS3 | 0.04071 | 2.782750812 | 3.12083 | 0.002294959 | 0.004903758 | -3.353448284 | Higher in PA |
| ACP5 | 0.06129 | 3.664339912 | 3.118455 | 0.002312021 | 0.004933188 | -3.360316259 | Higher in PA |
| PC | -0.06162 | 2.756522128 | -3.11749 | 0.002319003 | 0.004941057 | -3.363111961 | Higher in NT |
| TGM1 | 0.05811 | 2.391423953 | 3.11659 | 0.00232551 | 0.004947894 | -3.36570984 | Higher in PA |
| HSPG2 | -0.07385 | 3.259663585 | -3.10888 | 0.002382054 | 0.005061021 | -3.387978063 | Higher in NT |
| SPHK2 | -0.03764 | 2.772577606 | -3.10352 | 0.002422064 | 0.00513875 | -3.403413403 | Higher in NT |
| ERAP1 | -0.16582 | 3.770166775 | -3.10156 | 0.002436848 | 0.005162814 | -3.409051534 | Higher in NT |
| TNC | 0.09655 | 3.492910077 | 3.099019 | 0.002456176 | 0.005196423 | -3.41637016 | Higher in PA |
| AXIN2 | 0.09267 | 3.108130316 | 3.09738 | 0.00246871 | 0.005215584 | -3.421085106 | Higher in PA |
| ACTN2 | 0.20933 | 2.970029215 | 3.09625 | 0.002477382 | 0.005226544 | -3.424333177 | Higher in PA |
| IGF2.1 | 0.07964 | 3.186782662 | 3.094293 | 0.002492471 | 0.005250991 | -3.429957105 | Higher in PA |
| STAT5A | -0.07264 | 3.504758871 | -3.07896 | 0.002613672 | 0.005498609 | -3.473916552 | Higher in NT |

| | | | | | | | |
|---|---|---|---|---|---|---|---|
| MICA | 0.11209 | 2.206328428 | 3.075297 | 0.002643404 | 0.005553371 | -3.484384342 | Higher in PA |
| IL11RA | 0.04263 | 2.814112634 | 3.074558 | 0.002649444 | 0.005558275 | -3.48649622 | Higher in PA |
| TMPO | -0.12208 | 3.46384254 | -3.07404 | 0.002653666 | 0.005559357 | -3.487969454 | Higher in NT |
| ADAMTS13 | 0.06123 | 4.315119004 | 3.069695 | 0.002689506 | 0.005626582 | -3.50038124 | Higher in PA |
| TNNI3 | 0.06220 | 2.501972856 | 3.059188 | 0.002777991 | 0.005803602 | -3.530319136 | Higher in PA |
| TFF1 | 0.14227 | 3.126852421 | 3.058014 | 0.002788048 | 0.005816511 | -3.53366037 | Higher in PA |
| PCSK2 | 0.05671 | 2.914489785 | 3.056566 | 0.002800488 | 0.00583435 | -3.537776464 | Higher in PA |
| APEX1 | -0.10635 | 4.163012974 | -3.0537 | 0.002825253 | 0.00587778 | -3.545915448 | Higher in NT |
| THPO | -0.08156 | 2.864248091 | -3.05172 | 0.002842534 | 0.005905541 | -3.551552125 | Higher in NT |
| ANXA1 | 0.10955 | 3.789674272 | 3.047868 | 0.002876338 | 0.005967505 | -3.562478135 | Higher in PA |
| FADD | -0.06837 | 3.513417637 | -3.04522 | 0.002899766 | 0.006007803 | -3.569974471 | Higher in NT |
| VEGFD | 0.11157 | 3.886352356 | 3.030955 | 0.003029281 | 0.006267478 | -3.610335495 | Higher in PA |
| GNAS | -0.05565 | 3.181212651 | -3.02662 | 0.003069695 | 0.006342345 | -3.622571584 | Higher in NT |
| KLK11 | 0.05683 | 3.064215839 | 3.019148 | 0.003140471 | 0.006479652 | -3.643611427 | Higher in PA |
| PRAME | -0.06571 | 3.318171237 | -3.01395 | 0.003190596 | 0.00657403 | -3.658222618 | Higher in NT |
| COMP | -0.04988 | 2.937144949 | -3.01329 | 0.003197007 | 0.006578203 | -3.660074552 | Higher in NT |
| UST | 0.05185 | 2.838930625 | 3.010298 | 0.003226267 | 0.006629316 | -3.668479314 | Higher in PA |
| CD7 | 0.10929 | 2.692213675 | 3.009746 | 0.003231694 | 0.006631383 | -3.670029592 | Higher in PA |
| DSG2 | -0.05949 | 2.781804212 | -3.00174 | 0.003311333 | 0.006785519 | -3.692480885 | Higher in NT |
| ANGPTL4 | -0.05288 | 3.558934331 | -2.99759 | 0.003353267 | 0.006862075 | -3.704083088 | Higher in NT |
| PRNP | -0.04266 | 2.987599917 | -2.99189 | 0.003411716 | 0.006972171 | -3.720010726 | Higher in NT |
| GFAP | -0.05862 | 2.690818111 | -2.98107 | 0.003525253 | 0.007194393 | -3.750172766 | Higher in NT |
| HTRA1 | 0.06116 | 4.198558021 | 2.977752 | 0.00356076 | 0.007256983 | -3.75940307 | Higher in PA |
| ITGA5 | 0.03972 | 3.230761005 | 2.971471 | 0.003628884 | 0.007385788 | -3.776853284 | Higher in PA |
| APCS | 0.04342 | 4.806544076 | 2.970904 | 0.00363509 | 0.007388395 | -3.778426485 | Higher in PA |
| CRABP2 | 0.09522 | 4.12849345 | 2.966117 | 0.003687901 | 0.007485591 | -3.791703502 | Higher in PA |
| IFNGR1 | 0.07354 | 2.669744688 | 2.963995 | 0.003711538 | 0.007523388 | -3.797583406 | Higher in PA |
| C6 | 0.02670 | 4.785479497 | 2.955698 | 0.003805291 | 0.007692951 | -3.820536502 | Higher in PA |
| MXRA7 | 0.06640 | 2.943412627 | 2.955684 | 0.003805447 | 0.007692951 | -3.820574014 | Higher in PA |
| VAV3 | -0.08468 | 3.136752806 | -2.95298 | 0.003836468 | 0.007745224 | -3.828041967 | Higher in NT |
| GRB14 | -0.06679 | 2.882033886 | -2.95109 | 0.003858243 | 0.007778716 | -3.833247519 | Higher in NT |
| GHR | 0.07113 | 3.021427654 | 2.94518 | 0.003927276 | 0.007907267 | -3.849554743 | Higher in PA |
| RET | 0.09625 | 3.155327815 | 2.942303 | 0.00396126 | 0.007964999 | -3.857475795 | Higher in PA |
| ITGB3\|ITGAV | 0.10273 | 3.388055885 | 2.935911 | 0.004037753 | 0.008107938 | -3.875055046 | Higher in PA |

| | | | | | | | |
|---|---|---|---|---|---|---|---|
| ACVR1 | -0.12549 | 2.824929853 | -2.92042 | 0.004228722 | 0.008480057 | -3.917504102 | Higher in NT |
| FGF1 | 0.04990 | 2.176368991 | 2.918617 | 0.004251548 | 0.008514449 | -3.922446996 | Higher in PA |
| PPM1L | -0.06131 | 2.795852852 | -2.91116 | 0.004346849 | 0.008693697 | -3.942795758 | Higher in NT |
| CSTB | -0.06405 | 3.082644818 | -2.9056 | 0.004419237 | 0.008826704 | -3.95795099 | Higher in NT |
| MASP1.1 | 0.14502 | 2.44764561 | 2.9003 | 0.004489298 | 0.008954717 | -3.972380377 | Higher in PA |
| AMBP | 0.03466 | 5.008014089 | 2.891934 | 0.004601895 | 0.009167122 | -3.995096427 | Higher in PA |
| IL4R | 0.07415 | 2.307867273 | 2.888833 | 0.004644274 | 0.009239272 | -4.003499915 | Higher in PA |
| PPARGC1A | -0.03306 | 2.635746585 | -2.8853 | 0.004693027 | 0.009323895 | -4.0130714 | Higher in NT |
| LDLR | 0.03298 | 2.193746546 | 2.873197 | 0.004863533 | 0.009649867 | -4.045766927 | Higher in PA |
| HSPA8 | -0.04098 | 3.399938007 | -2.86389 | 0.004998549 | 0.009904654 | -4.070839032 | Higher in NT |
| VCAM1 | 0.06603 | 3.77382426 | 2.853857 | 0.00514785 | 0.010187038 | -4.097772787 | Higher in PA |
| COLEC11 | 0.09369 | 3.412041283 | 2.851576 | 0.005182364 | 0.010241826 | -4.103885765 | Higher in PA |
| TF | 0.03466 | 5.31873815 | 2.850512 | 0.005198539 | 0.010260273 | -4.106736299 | Higher in PA |
| DGKB | 0.06505 | 3.254376175 | 2.849527 | 0.00521354 | 0.01027636 | -4.109371989 | Higher in PA |
| MYL4 | 0.03367 | 4.19851713 | 2.841578 | 0.005336172 | 0.010491453 | -4.130632494 | Higher in PA |
| ANGPTL3 | -0.05908 | 3.237650106 | -2.84155 | 0.005336652 | 0.010491453 | -4.130714737 | Higher in NT |
| RGS5 | 0.04431 | 3.130779242 | 2.840896 | 0.005346803 | 0.010497649 | -4.132451916 | Higher in PA |
| PDE3A | -0.03455 | 2.666697686 | -2.83885 | 0.005378792 | 0.010546651 | -4.137904938 | Higher in NT |
| TNFRSF9 | 0.02892 | 2.652593545 | 2.834466 | 0.005448113 | 0.010668629 | -4.149609028 | Higher in PA |
| PSMA6 | -0.06211 | 3.033046646 | -2.81827 | 0.005711023 | 0.011168884 | -4.192658727 | Higher in NT |
| PEX26 | 0.03871 | 2.820251509 | 2.811222 | 0.005829109 | 0.011384979 | -4.211340929 | Higher in PA |
| MFAP5 | 0.08384 | 3.312480516 | 2.809047 | 0.005865985 | 0.011442103 | -4.217095938 | Higher in PA |
| LPIN1 | -0.12070 | 3.165848429 | -2.80428 | 0.005947486 | 0.011586012 | -4.229685561 | Higher in NT |
| INHBB | 0.12284 | 3.909819406 | 2.799701 | 0.006026838 | 0.011725366 | -4.241775143 | Higher in PA |
| GC | 0.08351 | 4.212024868 | 2.79483 | 0.006112281 | 0.011876193 | -4.254612432 | Higher in PA |
| KITLG | 0.06435 | 3.322369929 | 2.778279 | 0.006410863 | 0.012428713 | -4.29807721 | Higher in PA |
| PNPLA2 | 0.05692 | 2.847954486 | 2.778151 | 0.006413216 | 0.012428713 | -4.298411408 | Higher in PA |
| PCK2 | -0.16420 | 2.810392223 | -2.77658 | 0.00644233 | 0.012469026 | -4.302536931 | Higher in NT |
| GAL | -0.03932 | 3.088940422 | -2.77519 | 0.006468096 | 0.012502763 | -4.306172172 | Higher in NT |
| SPINK2 | 0.06850 | 2.572292417 | 2.761586 | 0.006725432 | 0.012983459 | -4.341688265 | Higher in PA |
| PDGFD | -0.12115 | 3.547360077 | -2.75949 | 0.006765854 | 0.013044705 | -4.347140692 | Higher in NT |
| LEFTY2 | 0.09669 | 2.978801565 | 2.758336 | 0.006788301 | 0.013071183 | -4.350154237 | Higher in PA |
| PCBD1 | 0.08709 | 3.147496935 | 2.75259 | 0.006900725 | 0.013270625 | -4.365095449 | Higher in PA |
| CD46 | 0.04401 | 3.266890912 | 2.743392 | 0.007084247 | 0.013606108 | -4.388959482 | Higher in PA |

| | | | | | | | |
|---|---|---|---|---|---|---|---|
| COL6A1 | -0.15411 | 4.056830629 | -2.73882 | 0.007177106 | 0.013766828 | -4.400794733 | Higher in NT |
| BLK | -0.04859 | 2.608532701 | -2.72656 | 0.007431529 | 0.014236645 | -4.432437229 | Higher in NT |
| BOC | 0.09731 | 3.18768761 | 2.713122 | 0.007720015 | 0.014770437 | -4.467003427 | Higher in PA |
| DDX58 | -0.04839 | 2.699310295 | -2.70613 | 0.007874129 | 0.015046107 | -4.484932053 | Higher in NT |
| CD70 | 0.03156 | 2.506127048 | 2.704827 | 0.007903047 | 0.015082151 | -4.488256144 | Higher in PA |
| CFAP300 | 0.04066 | 3.060824502 | 2.703756 | 0.007926966 | 0.015108575 | -4.490996157 | Higher in PA |
| ACADS | -0.04420 | 3.151454591 | -2.70005 | 0.008010158 | 0.015247763 | -4.500461068 | Higher in NT |
| TGFB2 | -0.04786 | 2.48101646 | -2.69556 | 0.008112221 | 0.015422473 | -4.511936441 | Higher in NT |
| SCN2B | 0.05509 | 2.643331427 | 2.691611 | 0.008202818 | 0.01557497 | -4.521999584 | Higher in PA |
| FGFR1 | 0.03341 | 2.861023777 | 2.68441 | 0.008370475 | 0.015873214 | -4.540325943 | Higher in PA |
| EFEMP1 | -0.05462 | 3.482073698 | -2.67943 | 0.008488146 | 0.016076034 | -4.552965502 | Higher in NT |
| DNAJB13 | -0.08686 | 3.005611912 | -2.67555 | 0.008580944 | 0.016231294 | -4.562807575 | Higher in NT |
| LILRB5 | 0.09773 | 4.037087861 | 2.6735 | 0.008630381 | 0.016304245 | -4.56800634 | Higher in PA |
| UBE2T | -0.04149 | 2.818963997 | -2.67019 | 0.008710785 | 0.016435444 | -4.576397003 | Higher in NT |
| MRE11 | -0.04954 | 3.017510785 | -2.66777 | 0.00876987 | 0.016526138 | -4.582512504 | Higher in NT |
| NDUFB10 | -0.03391 | 2.675394466 | -2.65532 | 0.009079904 | 0.017088904 | -4.613925946 | Higher in NT |
| PSAP | 0.07854 | 3.267399507 | 2.64412 | 0.009367075 | 0.017607284 | -4.642058173 | Higher in PA |
| GALNS | 0.06709 | 3.291897171 | 2.642313 | 0.009414188 | 0.017673694 | -4.646588984 | Higher in PA |
| TLE5 | 0.03831 | 2.691812024 | 2.633762 | 0.009640067 | 0.018071594 | -4.667994067 | Higher in PA |
| APOC2 | 0.06476 | 2.713775701 | 2.633381 | 0.009650231 | 0.018071594 | -4.668945163 | Higher in PA |
| PKD2.1 | 0.03527 | 2.51539832 | 2.631954 | 0.009688437 | 0.018120519 | -4.672510987 | Higher in PA |
| RXRA | 0.02830 | 2.736564439 | 2.631269 | 0.009706828 | 0.018132306 | -4.674222272 | Higher in PA |
| CYCS | -0.05687 | 3.171298424 | -2.63078 | 0.009719985 | 0.018134301 | -4.675444554 | Higher in NT |
| MSTN\|GDF11 | 0.05869 | 2.982072906 | 2.629628 | 0.009751003 | 0.018169571 | -4.678319369 | Higher in PA |
| HK1 | -0.04631 | 3.363066049 | -2.62499 | 0.009876878 | 0.018381287 | -4.689890284 | Higher in NT |
| GDI1 | 0.06511 | 2.654049038 | 2.623529 | 0.009916807 | 0.018416658 | -4.69352908 | Higher in PA |
| ST6GAL1 | 0.02860 | 2.758630264 | 2.623396 | 0.00992044 | 0.018416658 | -4.693859468 | Higher in PA |
| KHK | -0.04787 | 3.180189253 | -2.61617 | 0.010120201 | 0.018764279 | -4.711835139 | Higher in NT |
| NPPB | 0.15671 | 3.234749855 | 2.609885 | 0.010296918 | 0.019068366 | -4.727436499 | Higher in PA |
| APOA2 | 0.06868 | 3.523160142 | 2.606942 | 0.010380608 | 0.019199645 | -4.734729566 | Higher in PA |
| CYB5A | 0.08499 | 3.057452184 | 2.598193 | 0.010633064 | 0.019642359 | -4.75636958 | Higher in PA |
| ESRRA | -0.04844 | 2.963285105 | -2.59075 | 0.010852125 | 0.020022371 | -4.774724319 | Higher in NT |
| RETN | 0.08952 | 4.28508518 | 2.583649 | 0.011064984 | 0.020390019 | -4.79219886 | Higher in PA |
| VHL | -0.04084 | 2.568490514 | -2.58015 | 0.01117132 | 0.020560712 | -4.800799748 | Higher in NT |

| | | | | | | | |
|---|---|---|---|---|---|---|---|
| B3GALT6 | 0.03012 | 2.63853999 | 2.576889 | 0.011271045 | 0.020718833 | -4.808789852 | Higher in PA |
| KLK8 | 0.05305 | 3.370957846 | 2.567589 | 0.011560215 | 0.021224385 | -4.831554841 | Higher in PA |
| CELA2A | 0.07792 | 3.293970023 | 2.565238 | 0.011634368 | 0.021334416 | -4.837298428 | Higher in PA |
| IL7R | 0.03645 | 2.642653167 | 2.551237 | 0.01208485 | 0.022133425 | -4.871402847 | Higher in PA |
| PHKA1 | -0.04438 | 2.835920045 | -2.54594 | 0.012259431 | 0.022425789 | -4.884270009 | Higher in NT |
| CDON | 0.04044 | 3.821547652 | 2.540415 | 0.012443728 | 0.022735191 | -4.897650612 | Higher in PA |
| RCAN1 | -0.08335 | 2.93489495 | -2.53703 | 0.012557994 | 0.022916047 | -4.905844851 | Higher in NT |
| FCGR2B | 0.10965 | 3.051078418 | 2.532182 | 0.01272312 | 0.023189161 | -4.917552056 | Higher in PA |
| COL18A1 | -0.06313 | 4.358192761 | -2.53072 | 0.012773282 | 0.023252334 | -4.921077584 | Higher in NT |
| FST | -0.04615 | 2.867439464 | -2.52744 | 0.012886613 | 0.023430206 | -4.928990667 | Higher in NT |
| PSMB5 | 0.03110 | 2.857827724 | 2.525061 | 0.012969303 | 0.02354977 | -4.93471928 | Higher in PA |
| CD38 | 0.02905 | 2.753234894 | 2.524646 | 0.012983773 | 0.02354977 | -4.935717887 | Higher in PA |
| DES | 0.04883 | 3.126770408 | 2.513792 | 0.013367629 | 0.024216719 | -4.961798805 | Higher in PA |
| MAD2L2 | -0.04273 | 2.887949675 | -2.5109 | 0.01347142 | 0.024375308 | -4.96871864 | Higher in NT |
| ALDH5A1 | -0.10620 | 2.97078031 | -2.51015 | 0.013498519 | 0.024394914 | -4.97051634 | Higher in NT |
| GP1BA | -0.05194 | 3.148296611 | -2.49956 | 0.013886451 | 0.025065797 | -4.995851699 | Higher in NT |
| POMC.1 | 0.06112 | 2.686171605 | 2.49859 | 0.013922285 | 0.025100273 | -4.998155101 | Higher in PA |
| IL11 | -0.04027 | 2.550111127 | -2.496 | 0.014018801 | 0.025243939 | -5.00432901 | Higher in NT |
| STX3 | 0.07896 | 2.678860607 | 2.482423 | 0.014534692 | 0.026141532 | -5.036604885 | Higher in PA |
| NR4A1 | -0.02191 | 2.678058855 | -2.46998 | 0.015022157 | 0.02696761 | -5.066036659 | Higher in NT |
| MAP2K2 | 0.07242 | 3.617828835 | 2.469788 | 0.015029948 | 0.02696761 | -5.066499042 | Higher in PA |
| ACVRL1 | 0.05719 | 2.539333404 | 2.460018 | 0.015423168 | 0.027640087 | -5.089520523 | Higher in PA |
| TBX5 | -0.03127 | 2.723735364 | -2.44904 | 0.015876013 | 0.028417684 | -5.115295168 | Higher in NT |
| CKM\|CKB | 0.15422 | 2.962066896 | 2.445114 | 0.016040721 | 0.028678285 | -5.124482436 | Higher in PA |
| CKM | 0.09917 | 2.973031114 | 2.442374 | 0.016156619 | 0.028851106 | -5.130889072 | Higher in PA |
| PSMB7 | -0.07901 | 2.823309805 | -2.43692 | 0.016389656 | 0.029208901 | -5.143628704 | Higher in NT |
| KCNE5 | 0.03531 | 2.471350774 | 2.436772 | 0.01639593 | 0.029208901 | -5.143969084 | Higher in PA |
| GDF2 | 0.04381 | 3.363240096 | 2.434362 | 0.016499843 | 0.029343893 | -5.149587493 | Higher in PA |
| UROS | 0.06447 | 3.207456324 | 2.434108 | 0.016510831 | 0.029343893 | -5.150179446 | Higher in PA |
| TAT | 0.02560 | 2.988806168 | 2.41985 | 0.017137995 | 0.030422476 | -5.183310729 | Higher in PA |
| SCT | 0.04169 | 2.252912926 | 2.413169 | 0.017439098 | 0.030920386 | -5.198775359 | Higher in PA |
| PAX4 | -0.04533 | 2.669490315 | -2.41144 | 0.017517923 | 0.031023476 | -5.202778303 | Higher in NT |
| JUP | 0.07640 | 2.755448111 | 2.408801 | 0.017638505 | 0.031200185 | -5.208865982 | Higher in PA |
| COG8 | 0.03703 | 2.6315185 | 2.406963 | 0.017723019 | 0.031312754 | -5.213107163 | Higher in PA |

| | | | | | | | |
|---|---|---|---|---|---|---|---|
| OCLN | -0.02872 | 2.641987732 | -2.4054 | 0.01779492 | 0.0314028 | -5.216699022 | Higher in NT |
| SAMHD1 | 0.07039 | 2.952250457 | 2.402767 | 0.017917266 | 0.03158155 | -5.222776547 | Higher in PA |
| TGFBR3 | 0.03726 | 4.329695205 | 2.390949 | 0.01847462 | 0.03252574 | -5.249931464 | Higher in PA |
| HPRT1 | -0.07479 | 3.456237277 | -2.38517 | 0.018752734 | 0.032976672 | -5.263166713 | Higher in NT |
| TFR2 | -0.04376 | 2.684370376 | -2.38363 | 0.018827478 | 0.033069341 | -5.266689166 | Higher in NT |
| ACVR2B | 0.02981 | 2.581777247 | 2.381994 | 0.01890714 | 0.033170421 | -5.270427516 | Higher in PA |
| NR3C2 | -0.11751 | 2.767675243 | -2.37334 | 0.01933342 | 0.033878656 | -5.290159267 | Higher in NT |
| MERTK | 0.10531 | 2.824761439 | 2.371361 | 0.019432361 | 0.034006355 | -5.294674809 | Higher in PA |
| TNFSF4 | -0.05141 | 2.807704086 | -2.37098 | 0.019451635 | 0.034006355 | -5.295551669 | Higher in NT |
| IL31RA | 0.02787 | 2.979393382 | 2.370197 | 0.01949063 | 0.034034861 | -5.297323002 | Higher in PA |
| CTRC | -0.03637 | 2.505116368 | -2.36778 | 0.019612238 | 0.034207392 | -5.302823545 | Higher in NT |
| UNG | 0.09713 | 3.255851349 | 2.361868 | 0.019912116 | 0.034690098 | -5.316238306 | Higher in PA |
| CD80 | -0.06000 | 2.321615292 | -2.3605 | 0.019982217 | 0.034771839 | -5.319344078 | Higher in NT |
| TNFRSF10A | -0.05291 | 2.891762637 | -2.35388 | 0.020324105 | 0.035325791 | -5.334331479 | Higher in NT |
| KIT | 0.05619 | 4.171138132 | 2.350624 | 0.020493935 | 0.035579749 | -5.341679676 | Higher in PA |
| ASNS | 0.08582 | 3.512286211 | 2.340051 | 0.021054667 | 0.036500552 | -5.365501516 | Higher in PA |
| NTN4 | 0.04555 | 3.676584057 | 2.339709 | 0.021072985 | 0.036500552 | -5.366268645 | Higher in PA |
| ICAM2 | 0.06633 | 3.763832003 | 2.338691 | 0.021127748 | 0.036553198 | -5.368557822 | Higher in PA |
| KRT19 | -0.05219 | 3.213423714 | -2.33365 | 0.021400675 | 0.036979798 | -5.379875786 | Higher in NT |
| IFIH1 | 0.02337 | 2.909263139 | 2.333207 | 0.021424767 | 0.036979798 | -5.380867686 | Higher in PA |
| FCGR3B | 0.05497 | 3.49842347 | 2.332776 | 0.021448283 | 0.036979798 | -5.381834736 | Higher in PA |
| BMPR1B | -0.02885 | 2.560307898 | -2.32943 | 0.02163144 | 0.037252767 | -5.389329397 | Higher in NT |
| HJV | 0.03191 | 2.80592411 | 2.326079 | 0.021816384 | 0.037528184 | -5.396830802 | Higher in PA |
| COL5A1 | 0.06576 | 3.30779009 | 2.324257 | 0.021917472 | 0.037623664 | -5.400903159 | Higher in PA |
| NDRG2 | -0.04510 | 2.899147316 | -2.32417 | 0.021922055 | 0.037623664 | -5.401087325 | Higher in NT |
| EDN1 | 0.03926 | 2.722662008 | 2.319055 | 0.022208428 | 0.038071591 | -5.412516784 | Higher in PA |
| PRSS2 | 0.09011 | 3.222141807 | 2.3117 | 0.022625609 | 0.038742481 | -5.428896564 | Higher in PA |
| MRAP2 | -0.05536 | 2.972610034 | -2.3107 | 0.022682675 | 0.038795909 | -5.431112795 | Higher in NT |
| TBK1 | -0.05320 | 2.573615731 | -2.30996 | 0.022725109 | 0.038824218 | -5.432757032 | Higher in NT |
| FAH | 0.06095 | 3.751016292 | 2.296794 | 0.023492262 | 0.040089183 | -5.46194586 | Higher in PA |
| ADA2 | 0.09376 | 4.008133673 | 2.282436 | 0.024354536 | 0.041513413 | -5.493596586 | Higher in PA |
| PLAU | 0.03809 | 3.27335423 | 2.273391 | 0.024911909 | 0.042415283 | -5.513442868 | Higher in PA |
| IDO1 | 0.09453 | 2.590709679 | 2.269793 | 0.025136676 | 0.04274945 | -5.521316013 | Higher in PA |
| SERPIND1 | 0.03566 | 3.701800385 | 2.264306 | 0.02548298 | 0.043289321 | -5.533304404 | Higher in PA |

| | | | | | | | |
|---|---|---|---|---|---|---|---|
| GNMT | 0.02287 | 3.14552157 | 2.262425 | 0.025602658 | 0.043443425 | -5.537408132 | Higher in PA |
| PTK2 | -0.07614 | 2.133224001 | -2.26117 | 0.025682612 | 0.043529851 | -5.540138653 | Higher in NT |
| UNC13A | 0.03688 | 2.548911854 | 2.257139 | 0.025941552 | 0.043919106 | -5.548921493 | Higher in PA |
| IFNA2 | 0.02581 | 1.926160553 | 2.255227 | 0.026065099 | 0.044078521 | -5.553079993 | Higher in PA |
| CLEC11A | -0.08139 | 3.55673047 | -2.24983 | 0.026416351 | 0.044622214 | -5.564791877 | Higher in NT |
| CALB2 | 0.09531 | 2.608980858 | 2.244287 | 0.026782 | 0.045188976 | -5.57681311 | Higher in PA |
| SPN | 0.06499 | 2.821106903 | 2.243427 | 0.026839044 | 0.045234345 | -5.578673149 | Higher in PA |
| RUNX3 | -0.05769 | 2.856913981 | -2.24064 | 0.027024744 | 0.045496203 | -5.584699837 | Higher in NT |
| FGF2 | -0.14984 | 3.155968437 | -2.23861 | 0.027160522 | 0.045673524 | -5.589079181 | Higher in NT |
| BAG3 | 0.03085 | 2.800679402 | 2.237665 | 0.027224248 | 0.045729421 | -5.591126739 | Higher in PA |
| ANPEP | 0.08829 | 2.983893552 | 2.232488 | 0.027574476 | 0.0462659 | -5.602291532 | Higher in PA |
| F2 | 0.07213 | 3.216906818 | 2.230569 | 0.027705256 | 0.04643339 | -5.606422837 | Higher in PA |
| F13B | 0.06373 | 4.438967343 | 2.226723 | 0.027969083 | 0.046816027 | -5.61469569 | Higher in PA |
| CD2 | 0.15718 | 2.836675853 | 2.226332 | 0.027995984 | 0.046816027 | -5.615534661 | Higher in PA |
| PRKCQ | -0.09787 | 3.295310213 | -2.22545 | 0.028057046 | 0.04686589 | -5.617435931 | Higher in NT |
| AGER | 0.11885 | 3.382806752 | 2.216201 | 0.028701979 | 0.047889842 | -5.637257879 | Higher in PA |
| GNRH1 | 0.04931 | 3.126783791 | 2.211169 | 0.029058377 | 0.048430628 | -5.648013791 | Higher in PA |
| IGF1 | 0.04365 | 2.275273198 | 2.207412 | 0.029327029 | 0.048824133 | -5.65603118 | Higher in PA |
| GSTA4 | 0.06311 | 2.931204031 | 2.206815 | 0.029369887 | 0.048841275 | -5.657303085 | Higher in PA |
| IGFBP3 | 0.05200 | 4.308146603 | 2.202277 | 0.029697673 | 0.049331682 | -5.666967593 | Higher in PA |
