## Supplemental Table 3 for "The Evolution of Primary Aldosteronism and the Role of Norrin"

| GeneID | logFC | AveExpr | t | P.Value | adj.P.Val | B | Significance |
|---|---|---|---|---|---|---|---|
| CYBRD1 | 1.433217 | 2.845971 | 44.15096 | 1.19E-68 | 1.78E-65 | 144.9169 | Higher in PA |
| F11 | 0.757012 | 3.148904 | 33.00602 | 1.6E-56 | 8.02E-54 | 118.0272 | Higher in PA |
| ITGA3\|ITG | 0.431353 | 2.574196 | 7.869355 | 3.76E-12 | 1.53E-10 | 16.89407 | Higher in PA |
| S100A6 | 0.441123 | 3.500188 | 7.05122 | 2.11E-10 | 4.65E-09 | 12.91725 | Higher in PA |
| NDP | 0.272997 | 2.603316 | 7.002161 | 2.67E-10 | 5.81E-09 | 12.68319 | Higher in PA |
| BPGM | 0.484501 | 3.020993 | 6.891746 | 4.55E-10 | 9.61E-09 | 12.15854 | Higher in PA |
| GLO1 | 0.399091 | 3.770293 | 6.68309 | 1.23E-09 | 2.23E-08 | 11.17555 | Higher in PA |
| PRL | 0.494507 | 3.765626 | 6.431358 | 4.05E-09 | 6.2E-08 | 10.00564 | Higher in PA |
| PKLR | 0.479743 | 3.914814 | 5.960736 | 3.56E-08 | 4.42E-07 | 7.871574 | Higher in PA |
| CA1 | 0.358504 | 4.413494 | 5.599378 | 1.8E-07 | 1.93E-06 | 6.286706 | Higher in PA |
| LSP1 | 0.212609 | 2.769777 | 5.57335 | 2.02E-07 | 2.12E-06 | 6.174537 | Higher in PA |
| PCNA | 0.371582 | 2.703826 | 5.442909 | 3.58E-07 | 3.6E-06 | 5.616652 | Higher in PA |
| HMBS | 0.415891 | 4.536682 | 5.360495 | 5.12E-07 | 4.79E-06 | 5.267929 | Higher in PA |
| HMGB2 | 0.251615 | 2.68962 | 5.359518 | 5.14E-07 | 4.79E-06 | 5.263813 | Higher in PA |
| TARDBP | 0.121913 | 2.658376 | 5.234372 | 8.8E-07 | 7.77E-06 | 4.740085 | Higher in PA |
| NCAM1 | 0.142612 | 3.895053 | 5.102508 | 1.54E-06 | 1.26E-05 | 4.196023 | Higher in PA |
| OLR1 | 0.236031 | 3.05336 | 5.09778 | 1.57E-06 | 1.27E-05 | 4.176665 | Higher in PA |
| HK2 | 0.315469 | 2.800946 | 4.978883 | 2.59E-06 | 1.99E-05 | 3.693476 | Higher in PA |
| TNFRSF4 | 0.096586 | 2.819179 | 4.923003 | 3.26E-06 | 2.44E-05 | 3.46878 | Higher in PA |
| ARG1 | 0.431227 | 3.155933 | 4.896688 | 3.63E-06 | 2.7E-05 | 3.363509 | Higher in PA |
| PLXND1 | 0.145029 | 3.722776 | 4.836871 | 4.65E-06 | 3.37E-05 | 3.125525 | Higher in PA |
| COMT | 0.165336 | 3.101728 | 4.649254 | 9.93E-06 | 6.71E-05 | 2.391161 | Higher in PA |
| CCL2 | 0.100064 | 2.456059 | 4.628407 | 1.08E-05 | 7.2E-05 | 2.31072 | Higher in PA |
| SELE | 0.223272 | 4.277172 | 4.613493 | 1.15E-05 | 7.6E-05 | 2.253317 | Higher in PA |
| HBB\|HBA1 | 0.494288 | 2.933846 | 4.564997 | 1.39E-05 | 8.98E-05 | 2.067505 | Higher in PA |
| PDLIM4 | 0.24691 | 2.754032 | 4.564903 | 1.39E-05 | 8.98E-05 | 2.067149 | Higher in PA |
| KDR | 0.113414 | 3.755407 | 4.541923 | 1.52E-05 | 9.71E-05 | 1.979553 | Higher in PA |
| CA3 | 0.291726 | 3.25419 | 4.504644 | 1.76E-05 | 0.000112 | 1.838086 | Higher in PA |
| NOTCH1 | 0.094365 | 4.105544 | 4.470821 | 2.01E-05 | 0.000126 | 1.710406 | Higher in PA |
| CNTN1 | 0.11614 | 4.460893 | 4.442944 | 2.24E-05 | 0.000138 | 1.605657 | Higher in PA |
| MMP7 | 0.329751 | 3.919776 | 4.432746 | 2.33E-05 | 0.000143 | 1.56745 | Higher in PA |
| ACE | 0.151244 | 2.774368 | 4.418559 | 2.47E-05 | 0.000149 | 1.514396 | Higher in PA |
| ABCC6 | 0.086823 | 2.835004 | 4.396067 | 2.69E-05 | 0.000161 | 1.430522 | Higher in PA |
| EPHA3 | 0.082276 | 2.122722 | 4.344197 | 3.29E-05 | 0.000193 | 1.238212 | Higher in PA |
| MET | 0.101063 | 3.198886 | 4.30717 | 3.79E-05 | 0.000221 | 1.101897 | Higher in PA |
| SFRP1 | 0.249351 | 3.172295 | 4.303499 | 3.84E-05 | 0.000223 | 1.088426 | Higher in PA |
| THSD1 | 0.141188 | 2.801717 | 4.292291 | 4.01E-05 | 0.000232 | 1.047346 | Higher in PA |
| ALCAM | 0.090784 | 3.996595 | 4.278219 | 4.23E-05 | 0.000243 | 0.995879 | Higher in PA |
| LRIG1 | 0.118888 | 2.922697 | 4.246443 | 4.78E-05 | 0.000271 | 0.880085 | Higher in PA |
| CD14 | 0.221909 | 4.119502 | 4.216485 | 5.35E-05 | 0.000302 | 0.771472 | Higher in PA |
| HIP1R | 0.062779 | 2.561424 | 4.04197 | 0.000103 | 0.000555 | 0.149666 | Higher in PA |
| S100A9\|S1 | 0.273024 | 2.887864 | 3.965251 | 0.000136 | 0.000722 | -0.11767 | Higher in PA |
| AMN | 0.107023 | 2.216262 | 3.868721 | 0.000192 | 0.000987 | -0.44868 | Higher in PA |
| NODAL | 0.075759 | 2.439367 | 3.783347 | 0.00026 | 0.001281 | -0.73635 | Higher in PA |
| CDH5 | 0.106174 | 4.118198 | 3.783227 | 0.00026 | 0.001281 | -0.73675 | Higher in PA |
| TTN | 0.116309 | 2.622652 | 3.738998 | 0.000304 | 0.001476 | -0.88388 | Higher in PA |

| | | | | | | | |
|---|---|---|---|---|---|---|---|
| C1orf185 | 0.084083 | 2.185035 | 3.692391 | 0.000358 | 0.001703 | -1.03749 | Higher in PA |
| SCARB1 | 0.164091 | 3.035075 | 3.680939 | 0.000372 | 0.001767 | -1.07501 | Higher in PA |
| EGFR | 0.082384 | 4.129976 | 3.67187 | 0.000384 | 0.001817 | -1.10466 | Higher in PA |
| FANCF | 0.087225 | 2.706221 | 3.648452 | 0.000416 | 0.001952 | -1.18096 | Higher in PA |
| CXCL16 | 0.096013 | 3.674528 | 3.604937 | 0.000483 | 0.002245 | -1.32174 | Higher in PA |
| ANXA1 | 0.225224 | 3.792205 | 3.591892 | 0.000505 | 0.002325 | -1.36368 | Higher in PA |
| CD38 | 0.071559 | 2.753092 | 3.58349 | 0.00052 | 0.002378 | -1.39064 | Higher in PA |
| NOTCH3 | 0.097832 | 4.574639 | 3.5536 | 0.000576 | 0.002624 | -1.48613 | Higher in PA |
| IL31RA | 0.07203 | 2.979594 | 3.527898 | 0.000628 | 0.002853 | -1.56775 | Higher in PA |
| GAA | 0.120565 | 4.040479 | 3.474203 | 0.000751 | 0.003325 | -1.73675 | Higher in PA |
| EPHA4 | 0.098101 | 4.18976 | 3.473125 | 0.000754 | 0.003327 | -1.74012 | Higher in PA |
| IL2 | 0.112055 | 3.013666 | 3.448693 | 0.000818 | 0.003566 | -1.81632 | Higher in PA |
| RNASEH1 | 0.139931 | 2.166408 | 3.436041 | 0.000853 | 0.003708 | -1.85562 | Higher in PA |
| SPR | 0.141277 | 2.619903 | 3.387996 | 0.000999 | 0.004294 | -2.00379 | Higher in PA |
| PLEKHM2 | 0.208155 | 3.487695 | 3.382842 | 0.001016 | 0.004354 | -2.01959 | Higher in PA |
| GALNT3 | 0.085232 | 2.748136 | 3.34496 | 0.00115 | 0.004885 | -2.13509 | Higher in PA |
| ITGB1\|ITG | 0.188422 | 2.440568 | 3.33163 | 0.0012 | 0.005044 | -2.17549 | Higher in PA |
| SPP1 | 0.134024 | 2.79932 | 3.304589 | 0.00131 | 0.005428 | -2.25704 | Higher in PA |
| GPHN | 0.106222 | 2.365056 | 3.293807 | 0.001356 | 0.005587 | -2.28941 | Higher in PA |
| LIF | 0.074349 | 2.767381 | 3.292408 | 0.001362 | 0.005587 | -2.2936 | Higher in PA |
| NPPA | 0.155904 | 3.09976 | 3.292177 | 0.001363 | 0.005587 | -2.29429 | Higher in PA |
| PNPLA2 | 0.118271 | 2.848701 | 3.259696 | 0.001513 | 0.006166 | -2.39124 | Higher in PA |
| SERPINA7 | 0.056534 | 4.850335 | 3.253987 | 0.001541 | 0.006246 | -2.4082 | Higher in PA |
| A2M | 0.110248 | 4.086724 | 3.247044 | 0.001575 | 0.006368 | -2.4288 | Higher in PA |
| IGF2.1 | 0.135484 | 3.18373 | 3.244404 | 0.001588 | 0.006387 | -2.43662 | Higher in PA |
| TEK | 0.088057 | 3.029054 | 3.243586 | 0.001592 | 0.006387 | -2.43904 | Higher in PA |
| SPARCL1 | 0.10447 | 3.915669 | 3.181799 | 0.001936 | 0.007622 | -2.62056 | Higher in PA |
| CAV3 | 0.074208 | 2.542742 | 3.180271 | 0.001945 | 0.007638 | -2.62501 | Higher in PA |
| GH1 | 0.091892 | 2.576002 | 3.179284 | 0.001951 | 0.007642 | -2.62789 | Higher in PA |
| MMP2 | 0.099542 | 4.183886 | 3.160527 | 0.002069 | 0.00802 | -2.68239 | Higher in PA |
| IGFBP7 | 0.071776 | 4.484231 | 3.158182 | 0.002084 | 0.008059 | -2.68919 | Higher in PA |
| CR2 | 0.165398 | 3.775273 | 3.145788 | 0.002167 | 0.008333 | -2.72503 | Higher in PA |
| C4B\|C4A | 0.076515 | 5.142369 | 3.13477 | 0.002242 | 0.008602 | -2.7568 | Higher in PA |
| KLK11 | 0.095416 | 3.062718 | 3.111493 | 0.00241 | 0.009136 | -2.82362 | Higher in PA |
| SURF1 | 0.163874 | 3.817088 | 3.111243 | 0.002412 | 0.009136 | -2.82433 | Higher in PA |
| TNXB | 0.123726 | 3.953885 | 3.105335 | 0.002456 | 0.009281 | -2.84122 | Higher in PA |
| GDF2 | 0.093827 | 3.362419 | 3.080468 | 0.002652 | 0.009944 | -2.91203 | Higher in PA |
| IL6ST | 0.081336 | 3.837616 | 3.079418 | 0.00266 | 0.009951 | -2.91501 | Higher in PA |
| ST6GAL1 | 0.059071 | 2.758329 | 3.063475 | 0.002794 | 0.010424 | -2.96014 | Higher in PA |
| JAG1 | 0.145061 | 3.199483 | 3.060518 | 0.002819 | 0.010492 | -2.96849 | Higher in PA |
| MYBPC3 | 0.105721 | 2.323946 | 3.058079 | 0.00284 | 0.010544 | -2.97537 | Higher in PA |
| RFX5 | 0.097219 | 2.92494 | 3.034618 | 0.00305 | 0.011298 | -3.04134 | Higher in PA |
| STX3 | 0.169621 | 2.679003 | 3.033769 | 0.003058 | 0.011299 | -3.04372 | Higher in PA |
| MALT1 | 0.091727 | 2.442503 | 3.026337 | 0.003128 | 0.011529 | -3.06452 | Higher in PA |
| PON1 | 0.082796 | 2.545795 | 3.012236 | 0.003265 | 0.011973 | -3.10387 | Higher in PA |
| AGGF1 | 0.096899 | 2.132619 | 3.007968 | 0.003307 | 0.012099 | -3.11576 | Higher in PA |
| LOXL2 | 0.060767 | 2.459772 | 3.003839 | 0.003348 | 0.012221 | -3.12723 | Higher in PA |

| | | | | | | | |
|---|---|---|---|---|---|---|---|
| FAM20A | 0.125981 | 3.093377 | 2.992041 | 0.00347 | 0.012632 | -3.15996 | Higher in PA |
| IL7R | 0.074769 | 2.642401 | 2.978126 | 0.003618 | 0.01314 | -3.19843 | Higher in PA |
| RRAS2 | 0.085146 | 2.778306 | 2.958805 | 0.003833 | 0.013855 | -3.25159 | Higher in PA |
| BMP6 | 0.098022 | 3.13678 | 2.945633 | 0.003987 | 0.014341 | -3.28766 | Higher in PA |
| LMNB2 | 0.163745 | 3.301235 | 2.939173 | 0.004064 | 0.014515 | -3.3053 | Higher in PA |
| F2 | 0.165494 | 3.218188 | 2.932191 | 0.004149 | 0.014784 | -3.32434 | Higher in PA |
| AGRP | 0.159275 | 3.004313 | 2.928003 | 0.004201 | 0.014933 | -3.33573 | Higher in PA |
| RXRA | 0.0557 | 2.737023 | 2.922392 | 0.004272 | 0.015139 | -3.35098 | Higher in PA |
| RPS6KB1 | 0.132766 | 2.869212 | 2.921785 | 0.004279 | 0.015139 | -3.35263 | Higher in PA |
| B4GALT1 | 0.067993 | 4.306422 | 2.909331 | 0.00444 | 0.01567 | -3.38638 | Higher in PA |
| LTA | 0.0707 | 2.914383 | 2.902417 | 0.004531 | 0.015942 | -3.40506 | Higher in PA |
| TNFAIP6 | 0.144641 | 3.052425 | 2.901906 | 0.004538 | 0.015942 | -3.40644 | Higher in PA |
| PDGFRB | 0.158975 | 3.36351 | 2.852308 | 0.005248 | 0.018221 | -3.53934 | Higher in PA |
| CNDP1 | 0.14326 | 3.879215 | 2.822925 | 0.005715 | 0.019661 | -3.61715 | Higher in PA |
| CCL3 | 0.089615 | 2.79669 | 2.80569 | 0.006006 | 0.020569 | -3.66247 | Higher in PA |
| TFRC.1 | 0.112637 | 3.710776 | 2.80288 | 0.006055 | 0.020688 | -3.66984 | Higher in PA |
| CETP | 0.101768 | 2.804202 | 2.788572 | 0.006309 | 0.021459 | -3.70725 | Higher in PA |
| INSR | 0.17411 | 3.775284 | 2.788558 | 0.006309 | 0.021459 | -3.70728 | Higher in PA |
| CANX | 0.065946 | 2.40787 | 2.770727 | 0.006639 | 0.022531 | -3.75367 | Higher in PA |
| TMPRSS15 | 0.110838 | 2.568906 | 2.758024 | 0.006884 | 0.023152 | -3.78657 | Higher in PA |
| MPO | 0.144868 | 4.006152 | 2.747575 | 0.007091 | 0.023796 | -3.81353 | Higher in PA |
| NOS3 | 0.063224 | 2.782238 | 2.743956 | 0.007164 | 0.023988 | -3.82284 | Higher in PA |
| CDKN1B | 0.183278 | 2.870065 | 2.74104 | 0.007224 | 0.024084 | -3.83034 | Higher in PA |
| PYCR1 | 0.127167 | 2.563647 | 2.736719 | 0.007313 | 0.024211 | -3.84144 | Higher in PA |
| NCF1 | 0.224122 | 2.86291 | 2.735985 | 0.007328 | 0.024211 | -3.84333 | Higher in PA |
| TF | 0.058464 | 5.318387 | 2.734916 | 0.00735 | 0.024231 | -3.84607 | Higher in PA |
| PRTN3 | 0.19462 | 3.197174 | 2.732032 | 0.00741 | 0.024376 | -3.85346 | Higher in PA |
| ANTXR2 | 0.123598 | 3.764774 | 2.728141 | 0.007492 | 0.024591 | -3.86343 | Higher in PA |
| SMPD1 | 0.10009 | 2.970209 | 2.715558 | 0.007763 | 0.025313 | -3.89558 | Higher in PA |
| KL | 0.179539 | 3.38324 | 2.711647 | 0.007849 | 0.025449 | -3.90555 | Higher in PA |
| IFIH1 | 0.047109 | 2.909214 | 2.699637 | 0.008118 | 0.02613 | -3.93608 | Higher in PA |
| EWSR1 | 0.100497 | 3.334664 | 2.695148 | 0.00822 | 0.026404 | -3.94746 | Higher in PA |
| FSTL1 | 0.066776 | 4.281515 | 2.693311 | 0.008263 | 0.026446 | -3.95211 | Higher in PA |
| GBA | 0.162159 | 3.630314 | 2.693049 | 0.008269 | 0.026446 | -3.95277 | Higher in PA |
| CTHRC1 | 0.095753 | 4.645674 | 2.678111 | 0.008621 | 0.027339 | -3.9905 | Higher in PA |
| CHST14 | 0.082132 | 2.648596 | 2.671208 | 0.008788 | 0.027694 | -4.00787 | Higher in PA |
| TIMP2 | 0.064654 | 4.595673 | 2.631539 | 0.009808 | 0.030524 | -4.10694 | Higher in PA |
| LAMC1\|LA | 0.081821 | 4.180569 | 2.630097 | 0.009847 | 0.030582 | -4.11052 | Higher in PA |
| GM2A | 0.076776 | 3.03063 | 2.624431 | 0.010002 | 0.030998 | -4.12455 | Higher in PA |
| PLEKHA1 | 0.054261 | 2.456796 | 2.616839 | 0.010213 | 0.03152 | -4.14332 | Higher in PA |
| CD163 | 0.11218 | 3.325114 | 2.605915 | 0.010523 | 0.032278 | -4.17025 | Higher in PA |
| HAVCR1 | 0.150308 | 2.957685 | 2.604973 | 0.01055 | 0.032296 | -4.17256 | Higher in PA |
| CELA2A | 0.131294 | 3.291536 | 2.594806 | 0.010847 | 0.03307 | -4.19752 | Higher in PA |
| SERPINA1 | 0.085704 | 4.51434 | 2.575775 | 0.011423 | 0.034685 | -4.24401 | Higher in PA |
| UROS | 0.119892 | 3.206537 | 2.568635 | 0.011646 | 0.035079 | -4.26138 | Higher in PA |
| TCN2 | 0.093205 | 3.839028 | 2.551879 | 0.012186 | 0.036557 | -4.30196 | Higher in PA |
| CNTF | 0.045112 | 2.226477 | 2.541371 | 0.012535 | 0.037455 | -4.3273 | Higher in PA |

| | | | | | | | |
|---|---|---|---|---|---|---|---|
| MASP1.1 | 0.222035 | 2.441473 | 2.530542 | 0.012905 | 0.038483 | -4.35331 | Higher in PA |
| ACP2 | 0.217199 | 2.51215 | 2.524784 | 0.013105 | 0.039004 | -4.3671 | Higher in PA |
| KLK3\|SERP | 0.111565 | 3.927046 | 2.510342 | 0.01362 | 0.040383 | -4.40156 | Higher in PA |
| TCAP | 0.053688 | 3.061689 | 2.50957 | 0.013648 | 0.040383 | -4.4034 | Higher in PA |
| SFRP4 | 0.102608 | 3.87666 | 2.509544 | 0.013649 | 0.040383 | -4.40346 | Higher in PA |
| CAMK2A | 0.102029 | 2.928991 | 2.490245 | 0.014368 | 0.042341 | -4.44923 | Higher in PA |
| PTPRS | 0.064724 | 4.649383 | 2.482526 | 0.014664 | 0.04313 | -4.46745 | Higher in PA |
| TNFRSF10B | 0.09988 | 2.694832 | 2.475854 | 0.014925 | 0.043765 | -4.48316 | Higher in PA |
| INS | 0.121018 | 2.154833 | 2.469489 | 0.015178 | 0.044241 | -4.49811 | Higher in PA |
| TAGLN | 0.092628 | 3.268291 | 2.469196 | 0.015189 | 0.044241 | -4.4988 | Higher in PA |
| ANPEP | 0.17497 | 2.983604 | 2.466095 | 0.015314 | 0.044345 | -4.50607 | Higher in PA |
| PROC | 0.062574 | 4.874483 | 2.463934 | 0.015401 | 0.044512 | -4.51113 | Higher in PA |
| ROBO3 | 0.060281 | 2.715431 | 2.460068 | 0.015558 | 0.04488 | -4.52018 | Higher in PA |
| MGAT2 | 0.053666 | 3.004646 | 2.451927 | 0.015894 | 0.045761 | -4.53918 | Higher in PA |
| CHL1 | 0.073267 | 3.959709 | 2.44932 | 0.016003 | 0.045899 | -4.54526 | Higher in PA |
| FGF5 | 0.077271 | 2.681903 | 2.432497 | 0.016723 | 0.047779 | -4.58432 | Higher in PA |
| CCL20 | -1.18381 | 2.872118 | -34.1282 | 6.86E-58 | 5.14E-55 | 121.0928 | Higher in NT |
| ITGB1\|ITGA | -0.94618 | 3.122226 | -20.4259 | 5.68E-38 | 2.13E-35 | 76.00181 | Higher in NT |
| PTPN6 | -1.17014 | 3.35546 | -14.2568 | 4.24E-26 | 1.27E-23 | 48.82381 | Higher in NT |
| PSPH | -0.47409 | 3.538479 | -12.5619 | 1.72E-22 | 4.31E-20 | 40.55055 | Higher in NT |
| ATP5PF | -1.15942 | 3.517835 | -11.449 | 4.65E-20 | 9.96E-18 | 34.97756 | Higher in NT |
| NAMPT | -0.34462 | 3.197433 | -11.2529 | 1.26E-19 | 2.36E-17 | 33.98694 | Higher in NT |
| ECHS1 | -1.12928 | 3.958903 | -11.086 | 2.94E-19 | 4.9E-17 | 33.14273 | Higher in NT |
| MPI | -0.67168 | 3.447956 | -10.9348 | 6.35E-19 | 9.52E-17 | 32.3763 | Higher in NT |
| IVD | -1.114 | 4.020764 | -10.5332 | 4.93E-18 | 6.72E-16 | 30.33759 | Higher in NT |
| SYNE2 | -0.57083 | 3.431189 | -9.77784 | 2.35E-16 | 2.86E-14 | 26.49602 | Higher in NT |
| ACADM | -1.16007 | 3.171038 | -9.76735 | 2.48E-16 | 2.86E-14 | 26.44272 | Higher in NT |
| CHKB | -0.86607 | 3.452631 | -9.69074 | 3.67E-16 | 3.75E-14 | 26.05322 | Higher in NT |
| HSD17B4 | -0.8174 | 3.777537 | -9.68656 | 3.75E-16 | 3.75E-14 | 26.03198 | Higher in NT |
| DECR1 | -1.07494 | 4.071283 | -9.4381 | 1.34E-15 | 1.25E-13 | 24.76984 | Higher in NT |
| MYL9 | -0.93395 | 4.15582 | -9.39239 | 1.69E-15 | 1.49E-13 | 24.53786 | Higher in NT |
| SERPINH1 | -0.7867 | 3.137084 | -9.25183 | 3.46E-15 | 2.88E-13 | 23.82501 | Higher in NT |
| HMGCL | -0.96325 | 3.334281 | -8.77384 | 3.95E-14 | 3.12E-12 | 21.40892 | Higher in NT |
| GFPT1 | -0.44381 | 3.575983 | -8.60881 | 9.12E-14 | 6.84E-12 | 20.57861 | Higher in NT |
| CSRP3 | -0.42458 | 2.948106 | -8.51998 | 1.43E-13 | 1.02E-11 | 20.1327 | Higher in NT |
| PRKAR1A | -0.59182 | 4.176046 | -8.45372 | 2E-13 | 1.36E-11 | 19.80061 | Higher in NT |
| CTSC | -0.33589 | 3.956238 | -8.44569 | 2.08E-13 | 1.36E-11 | 19.76038 | Higher in NT |
| AKT1S1 | -0.6288 | 3.4372 | -8.38678 | 2.8E-13 | 1.75E-11 | 19.46554 | Higher in NT |
| NDUFV2 | -0.49078 | 3.40392 | -8.25722 | 5.39E-13 | 3.22E-11 | 18.81845 | Higher in NT |
| LTA4H | -0.3919 | 3.491416 | -8.24627 | 5.69E-13 | 3.22E-11 | 18.76388 | Higher in NT |
| ITGB3\|ITGA | -0.85463 | 4.496261 | -8.24278 | 5.79E-13 | 3.22E-11 | 18.74649 | Higher in NT |
| GRHPR | -0.41094 | 3.456518 | -8.2348 | 6.03E-13 | 3.23E-11 | 18.70671 | Higher in NT |
| STIM1 | -0.64574 | 3.997089 | -8.21854 | 6.54E-13 | 3.38E-11 | 18.62566 | Higher in NT |
| BOLA3 | -0.74567 | 3.299959 | -8.03108 | 1.68E-12 | 8.38E-11 | 17.69394 | Higher in NT |
| COX5A | -0.90583 | 3.684205 | -7.98544 | 2.11E-12 | 1.02E-10 | 17.46784 | Higher in NT |
| CCDC92 | -0.67635 | 3.431017 | -7.9786 | 2.18E-12 | 1.02E-10 | 17.434 | Higher in NT |
| DBT | -0.81244 | 3.200905 | -7.97231 | 2.25E-12 | 1.02E-10 | 17.40283 | Higher in NT |

| | | | | | | | |
|---|---|---|---|---|---|---|---|
| OXCT1 | -0.67277 | 4.194299 | -7.9424 | 2.61E-12 | 1.15E-10 | 17.25485 | Higher in NT |
| HSPA1A | -0.28725 | 3.361648 | -7.93546 | 2.7E-12 | 1.16E-10 | 17.22057 | Higher in NT |
| HSPB1 | -0.75261 | 3.517917 | -7.89865 | 3.25E-12 | 1.35E-10 | 17.03867 | Higher in NT |
| JAK2 | -0.61361 | 3.818189 | -7.8616 | 3.91E-12 | 1.54E-10 | 16.85583 | Higher in NT |
| BCAT2 | -0.74508 | 3.367205 | -7.83595 | 4.44E-12 | 1.71E-10 | 16.72931 | Higher in NT |
| SEPTIN11 | -0.29439 | 3.478142 | -7.79877 | 5.35E-12 | 2.01E-10 | 16.54621 | Higher in NT |
| PRKAR2B | -0.61294 | 3.469649 | -7.76217 | 6.41E-12 | 2.35E-10 | 16.3661 | Higher in NT |
| DUSP6 | -0.165 | 3.200525 | -7.6872 | 9.31E-12 | 3.32E-10 | 15.99796 | Higher in NT |
| AMT | -0.80024 | 3.675339 | -7.664 | 1.04E-11 | 3.64E-10 | 15.88418 | Higher in NT |
| NME4 | -0.75994 | 3.771477 | -7.64581 | 1.14E-11 | 3.79E-10 | 15.79511 | Higher in NT |
| FH | -0.75493 | 3.214726 | -7.64338 | 1.16E-11 | 3.79E-10 | 15.7832 | Higher in NT |
| HIBCH | -0.85109 | 3.919703 | -7.64257 | 1.16E-11 | 3.79E-10 | 15.7792 | Higher in NT |
| CIRBP | -0.63399 | 4.244109 | -7.63529 | 1.2E-11 | 3.84E-10 | 15.74358 | Higher in NT |
| TFAM | -0.90825 | 4.005182 | -7.58391 | 1.55E-11 | 4.85E-10 | 15.49228 | Higher in NT |
| UBQLN2 | -0.4769 | 3.76224 | -7.51538 | 2.18E-11 | 6.67E-10 | 15.15782 | Higher in NT |
| MCEE | -0.60398 | 3.565518 | -7.50271 | 2.32E-11 | 6.95E-10 | 15.09611 | Higher in NT |
| UFD1 | -0.77418 | 3.8259 | -7.4814 | 2.57E-11 | 7.57E-10 | 14.99229 | Higher in NT |
| MESD | -0.54099 | 3.913544 | -7.45038 | 3E-11 | 8.65E-10 | 14.8414 | Higher in NT |
| HSPE1 | -0.77396 | 3.973182 | -7.41244 | 3.61E-11 | 1.02E-09 | 14.65711 | Higher in NT |
| RAB21 | -0.62226 | 4.647803 | -7.4082 | 3.69E-11 | 1.03E-09 | 14.63653 | Higher in NT |
| PYGB | -0.69562 | 4.386486 | -7.36939 | 4.47E-11 | 1.22E-09 | 14.44832 | Higher in NT |
| AKT2 | -0.64629 | 3.905174 | -7.35581 | 4.77E-11 | 1.28E-09 | 14.3825 | Higher in NT |
| PTP4A2 | -0.43069 | 3.080519 | -7.3447 | 5.04E-11 | 1.33E-09 | 14.32872 | Higher in NT |
| GOT2 | -0.64982 | 3.645119 | -7.31928 | 5.71E-11 | 1.48E-09 | 14.20576 | Higher in NT |
| HADH | -0.86511 | 4.187938 | -7.30319 | 6.18E-11 | 1.57E-09 | 14.12795 | Higher in NT |
| DARS2 | -1.03704 | 4.093085 | -7.28797 | 6.65E-11 | 1.66E-09 | 14.05442 | Higher in NT |
| FKBP14 | -0.44024 | 3.127506 | -7.24121 | 8.36E-11 | 2.06E-09 | 13.82888 | Higher in NT |
| UBQLN4 | -0.49947 | 3.635118 | -7.2144 | 9.53E-11 | 2.31E-09 | 13.69971 | Higher in NT |
| PUS1 | -0.34669 | 2.406378 | -7.20646 | 9.91E-11 | 2.36E-09 | 13.66153 | Higher in NT |
| CCM2 | -0.25421 | 3.51178 | -7.17056 | 1.18E-10 | 2.77E-09 | 13.48893 | Higher in NT |
| DDOST | -0.44079 | 3.297936 | -7.15953 | 1.25E-10 | 2.87E-09 | 13.43594 | Higher in NT |
| NDUFAF2 | -0.79366 | 3.440431 | -7.10347 | 1.64E-10 | 3.72E-09 | 13.16713 | Higher in NT |
| ALDH6A1 | -0.74238 | 3.683011 | -7.0654 | 1.97E-10 | 4.4E-09 | 12.98499 | Higher in NT |
| HTRA2 | -0.40661 | 3.490058 | -6.95213 | 3.4E-10 | 7.29E-09 | 12.44509 | Higher in NT |
| VPS26A | -0.45356 | 4.321792 | -6.88631 | 4.67E-10 | 9.63E-09 | 12.1328 | Higher in NT |
| GSK3B | -0.76036 | 3.553861 | -6.8854 | 4.69E-10 | 9.63E-09 | 12.12848 | Higher in NT |
| BCL2L1 | -0.29028 | 3.498661 | -6.87873 | 4.84E-10 | 9.81E-09 | 12.09689 | Higher in NT |
| CS | -0.75867 | 4.650421 | -6.86737 | 5.11E-10 | 1.02E-08 | 12.04313 | Higher in NT |
| PRDX3 | -0.67856 | 3.923414 | -6.86518 | 5.17E-10 | 1.02E-08 | 12.03276 | Higher in NT |
| ENO1 | -0.70708 | 4.782625 | -6.82249 | 6.34E-10 | 1.24E-08 | 11.83101 | Higher in NT |
| ABHD5 | -0.56536 | 2.982369 | -6.79893 | 7.1E-10 | 1.36E-08 | 11.71988 | Higher in NT |
| NDUFA2 | -0.49803 | 2.755822 | -6.77 | 8.15E-10 | 1.54E-08 | 11.58357 | Higher in NT |
| CD69 | -0.54601 | 2.44127 | -6.76825 | 8.22E-10 | 1.54E-08 | 11.57536 | Higher in NT |
| PDIA3 | -0.64484 | 3.754986 | -6.7529 | 8.84E-10 | 1.64E-08 | 11.50317 | Higher in NT |
| MAPK1 | -0.49661 | 3.274129 | -6.70014 | 1.14E-09 | 2.08E-08 | 11.25543 | Higher in NT |
| DLG4 | -0.35863 | 2.629863 | -6.66564 | 1.34E-09 | 2.39E-08 | 11.09389 | Higher in NT |
| MMAB | -0.56759 | 3.623234 | -6.65499 | 1.41E-09 | 2.49E-08 | 11.04407 | Higher in NT |

| | | | | | | | |
|---|---|---|---|---|---|---|---|
| FIS1 | -0.29055 | 3.33462 | -6.61436 | 1.71E-09 | 2.98E-08 | 10.85434 | Higher in NT |
| PDHX | -0.64368 | 3.404476 | -6.60442 | 1.79E-09 | 3.09E-08 | 10.808 | Higher in NT |
| HSPD1 | -0.72173 | 4.22604 | -6.59649 | 1.86E-09 | 3.17E-08 | 10.77101 | Higher in NT |
| TALDO1 | -0.29416 | 4.010107 | -6.58632 | 1.95E-09 | 3.29E-08 | 10.72365 | Higher in NT |
| GHRL | -0.70401 | 3.417436 | -6.55188 | 2.3E-09 | 3.83E-08 | 10.56347 | Higher in NT |
| COPB2 | -0.67302 | 4.25478 | -6.54847 | 2.34E-09 | 3.85E-08 | 10.54763 | Higher in NT |
| SMAD5 | -0.66282 | 3.775867 | -6.52995 | 2.55E-09 | 4.12E-08 | 10.46165 | Higher in NT |
| ACADVL | -0.85766 | 3.880437 | -6.52937 | 2.56E-09 | 4.12E-08 | 10.45895 | Higher in NT |
| UROD | -0.39019 | 3.613028 | -6.48873 | 3.09E-09 | 4.94E-08 | 10.27062 | Higher in NT |
| FLT3 | -0.34138 | 3.336863 | -6.4525 | 3.67E-09 | 5.79E-08 | 10.10317 | Higher in NT |
| RRAS | -0.20997 | 2.983355 | -6.44455 | 3.81E-09 | 5.95E-08 | 10.0665 | Higher in NT |
| CDK8\|CCN | -0.22565 | 3.228877 | -6.43166 | 4.05E-09 | 6.2E-08 | 10.00703 | Higher in NT |
| ACADSB | -0.47268 | 3.157992 | -6.42359 | 4.2E-09 | 6.37E-08 | 9.969817 | Higher in NT |
| AKT1 | -0.5178 | 3.270907 | -6.38321 | 5.08E-09 | 7.62E-08 | 9.784017 | Higher in NT |
| PPP3R1\|PI | -0.51981 | 4.300512 | -6.35737 | 5.73E-09 | 8.5E-08 | 9.665334 | Higher in NT |
| EPHX2 | -0.13085 | 2.415519 | -6.35388 | 5.82E-09 | 8.5E-08 | 9.649324 | Higher in NT |
| LZTFL1 | -0.4245 | 3.327001 | -6.35338 | 5.84E-09 | 8.5E-08 | 9.647072 | Higher in NT |
| ALG2 | -0.3521 | 3.152735 | -6.33716 | 6.3E-09 | 9.08E-08 | 9.572685 | Higher in NT |
| PRKCA | -0.69456 | 4.257375 | -6.32433 | 6.68E-09 | 9.55E-08 | 9.513931 | Higher in NT |
| FTH1 | -0.19959 | 3.348626 | -6.3209 | 6.79E-09 | 9.61E-08 | 9.498233 | Higher in NT |
| SMAD3 | -0.42473 | 3.784477 | -6.3018 | 7.42E-09 | 1.04E-07 | 9.410901 | Higher in NT |
| RHOC | -0.63068 | 3.888681 | -6.29358 | 7.71E-09 | 1.07E-07 | 9.373329 | Higher in NT |
| RASA1 | -0.30403 | 2.762923 | -6.2919 | 7.77E-09 | 1.07E-07 | 9.365655 | Higher in NT |
| ACAT1 | -0.64671 | 3.66679 | -6.27053 | 8.58E-09 | 1.17E-07 | 9.268127 | Higher in NT |
| HSPA2 | -0.27793 | 3.513019 | -6.25744 | 9.12E-09 | 1.23E-07 | 9.208483 | Higher in NT |
| SLC9A3R1 | -0.55723 | 4.446402 | -6.19693 | 1.21E-08 | 1.62E-07 | 8.93343 | Higher in NT |
| IL18 | -0.29332 | 2.931996 | -6.17503 | 1.34E-08 | 1.77E-07 | 8.834139 | Higher in NT |
| SLMAP | -0.47697 | 2.973141 | -6.17242 | 1.35E-08 | 1.78E-07 | 8.822353 | Higher in NT |
| FLNA.1 | -0.43415 | 4.340428 | -6.11601 | 1.75E-08 | 2.29E-07 | 8.567474 | Higher in NT |
| DDX6 | -0.58608 | 4.554016 | -6.10583 | 1.84E-08 | 2.38E-07 | 8.521565 | Higher in NT |
| ADSL | -0.41855 | 3.649857 | -6.09312 | 1.95E-08 | 2.5E-07 | 8.464337 | Higher in NT |
| ILK | -0.68507 | 4.020176 | -6.06975 | 2.17E-08 | 2.76E-07 | 8.359243 | Higher in NT |
| CRK | -0.34783 | 3.834998 | -5.97558 | 3.33E-08 | 4.2E-07 | 7.937721 | Higher in NT |
| PLA2G4A | -0.66178 | 3.515182 | -5.96504 | 3.5E-08 | 4.37E-07 | 7.890764 | Higher in NT |
| SNTA1 | -0.68767 | 3.924687 | -5.92959 | 4.11E-08 | 5.05E-07 | 7.733042 | Higher in NT |
| GBP2 | -0.22026 | 3.242095 | -5.89264 | 4.85E-08 | 5.92E-07 | 7.569117 | Higher in NT |
| HGD | -0.41349 | 2.987982 | -5.87272 | 5.31E-08 | 6.43E-07 | 7.48098 | Higher in NT |
| SUMO4 | -0.29264 | 3.192467 | -5.85988 | 5.63E-08 | 6.72E-07 | 7.424204 | Higher in NT |
| GRK2 | -0.52624 | 4.195185 | -5.85939 | 5.64E-08 | 6.72E-07 | 7.422064 | Higher in NT |
| MEIS2 | -0.47436 | 3.14042 | -5.8519 | 5.84E-08 | 6.89E-07 | 7.388984 | Higher in NT |
| CBR1 | -0.21057 | 3.377474 | -5.7973 | 7.46E-08 | 8.74E-07 | 7.148569 | Higher in NT |
| TPM4 | -0.79399 | 4.49953 | -5.79454 | 7.55E-08 | 8.78E-07 | 7.136408 | Higher in NT |
| PGAM2 | -0.42917 | 3.159187 | -5.77162 | 8.37E-08 | 9.66E-07 | 7.03587 | Higher in NT |
| PPCS | -0.45923 | 4.005475 | -5.76131 | 8.77E-08 | 1E-06 | 6.990687 | Higher in NT |
| PFN1 | -0.53341 | 4.605056 | -5.75598 | 8.98E-08 | 1.02E-06 | 6.967363 | Higher in NT |
| ELK3 | -0.46354 | 3.128715 | -5.73634 | 9.8E-08 | 1.11E-06 | 6.881471 | Higher in NT |
| CES3 | -0.52239 | 4.611756 | -5.70631 | 1.12E-07 | 1.25E-06 | 6.750395 | Higher in NT |

| | | | | | | | |
|---|---|---|---|---|---|---|---|
| XRCC4 | -0.26803 | 2.718075 | -5.6694 | 1.32E-07 | 1.47E-06 | 6.589825 | Higher in NT |
| ANXA11 | -0.52447 | 4.449391 | -5.66241 | 1.36E-07 | 1.5E-06 | 6.559482 | Higher in NT |
| AIFM1 | -0.32039 | 3.52619 | -5.64718 | 1.46E-07 | 1.6E-06 | 6.49343 | Higher in NT |
| GALK1 | -0.42802 | 3.866108 | -5.63317 | 1.55E-07 | 1.69E-06 | 6.432731 | Higher in NT |
| CASP3 | -0.55037 | 4.011921 | -5.62154 | 1.63E-07 | 1.76E-06 | 6.382435 | Higher in NT |
| HSPA9 | -0.37226 | 3.574627 | -5.58821 | 1.89E-07 | 2.01E-06 | 6.238524 | Higher in NT |
| CRKL | -0.62668 | 3.507771 | -5.57612 | 2E-07 | 2.11E-06 | 6.186443 | Higher in NT |
| SCFD1 | -0.361 | 3.035306 | -5.56945 | 2.06E-07 | 2.14E-06 | 6.157744 | Higher in NT |
| PLEK | -0.55511 | 4.880768 | -5.53947 | 2.35E-07 | 2.43E-06 | 6.028967 | Higher in NT |
| MAPRE1 | -0.66065 | 4.338874 | -5.52469 | 2.5E-07 | 2.57E-06 | 5.965581 | Higher in NT |
| GATM | -0.55097 | 3.863729 | -5.48036 | 3.04E-07 | 3.1E-06 | 5.776095 | Higher in NT |
| KRAS | -0.28082 | 2.168047 | -5.45458 | 3.4E-07 | 3.45E-06 | 5.666267 | Higher in NT |
| RAB7A | -0.50123 | 4.402492 | -5.43768 | 3.66E-07 | 3.66E-06 | 5.594445 | Higher in NT |
| AK2 | -0.48982 | 4.521613 | -5.42399 | 3.89E-07 | 3.86E-06 | 5.536323 | Higher in NT |
| KLF9 | -0.43282 | 3.506799 | -5.41708 | 4.01E-07 | 3.95E-06 | 5.50705 | Higher in NT |
| ENSA | -0.51226 | 3.471135 | -5.39204 | 4.47E-07 | 4.38E-06 | 5.40106 | Higher in NT |
| NFU1 | -0.30259 | 2.951919 | -5.38031 | 4.7E-07 | 4.58E-06 | 5.351493 | Higher in NT |
| STAT3 | -0.43857 | 4.225568 | -5.37847 | 4.74E-07 | 4.58E-06 | 5.343747 | Higher in NT |
| GNE | -0.20819 | 2.890045 | -5.37559 | 4.8E-07 | 4.61E-06 | 5.331574 | Higher in NT |
| TBCE | -0.53971 | 3.869782 | -5.36629 | 4.99E-07 | 4.75E-06 | 5.292361 | Higher in NT |
| POR | -0.30155 | 3.494791 | -5.36427 | 5.04E-07 | 4.75E-06 | 5.283859 | Higher in NT |
| FXN | -0.21197 | 2.84623 | -5.36407 | 5.04E-07 | 4.75E-06 | 5.283006 | Higher in NT |
| NRAS | -0.19633 | 3.247516 | -5.35029 | 5.35E-07 | 4.95E-06 | 5.224948 | Higher in NT |
| PTEN | -0.30345 | 2.798794 | -5.34339 | 5.51E-07 | 5.07E-06 | 5.195925 | Higher in NT |
| ARHGDIB | -0.45989 | 4.548703 | -5.31034 | 6.36E-07 | 5.81E-06 | 5.057165 | Higher in NT |
| NEIL1 | -0.21088 | 4.015663 | -5.30922 | 6.39E-07 | 5.81E-06 | 5.052472 | Higher in NT |
| MAPK14 | -0.47692 | 3.560111 | -5.30737 | 6.44E-07 | 5.82E-06 | 5.044714 | Higher in NT |
| SMAD4 | -0.33952 | 2.633926 | -5.27996 | 7.24E-07 | 6.5E-06 | 4.930036 | Higher in NT |
| PYGL | -0.47867 | 4.220356 | -5.26418 | 7.75E-07 | 6.92E-06 | 4.864175 | Higher in NT |
| ELMO2 | -0.36214 | 3.464183 | -5.2493 | 8.26E-07 | 7.33E-06 | 4.802173 | Higher in NT |
| ID1 | -0.21225 | 2.601633 | -5.22968 | 8.98E-07 | 7.88E-06 | 4.720589 | Higher in NT |
| PYCARD | -0.17508 | 4.152515 | -5.21281 | 9.65E-07 | 8.42E-06 | 4.650581 | Higher in NT |
| GNAQ | -0.4096 | 3.428888 | -5.20949 | 9.79E-07 | 8.44E-06 | 4.636803 | Higher in NT |
| MAPK3 | -0.39765 | 4.127178 | -5.20944 | 9.79E-07 | 8.44E-06 | 4.636608 | Higher in NT |
| MAPK8 | -0.25219 | 2.847524 | -5.18578 | 1.08E-06 | 9.28E-06 | 4.538642 | Higher in NT |
| SARS2 | -0.46338 | 3.429363 | -5.18151 | 1.1E-06 | 9.4E-06 | 4.520989 | Higher in NT |
| CD84 | -0.16884 | 3.613852 | -5.17058 | 1.15E-06 | 9.79E-06 | 4.475874 | Higher in NT |
| PKM | -0.63728 | 4.460963 | -5.16466 | 1.18E-06 | 9.98E-06 | 4.451453 | Higher in NT |
| STAR | -0.22647 | 3.099565 | -5.1517 | 1.25E-06 | 1.05E-05 | 4.398041 | Higher in NT |
| PDLIM5 | -0.41199 | 4.411586 | -5.14823 | 1.27E-06 | 1.06E-05 | 4.38375 | Higher in NT |
| LDHA | -0.26006 | 4.601985 | -5.13257 | 1.36E-06 | 1.12E-05 | 4.319349 | Higher in NT |
| NONO | -0.27903 | 2.875833 | -5.13059 | 1.37E-06 | 1.13E-05 | 4.311192 | Higher in NT |
| VCP | -0.50762 | 4.420597 | -5.11221 | 1.48E-06 | 1.21E-05 | 4.23579 | Higher in NT |
| ROCK2 | -0.48884 | 3.129632 | -5.07238 | 1.75E-06 | 1.41E-05 | 4.072884 | Higher in NT |
| EIF2A | -0.14329 | 2.80459 | -5.0677 | 1.78E-06 | 1.43E-05 | 4.053767 | Higher in NT |
| ASAH1 | -0.32999 | 3.112542 | -5.05761 | 1.86E-06 | 1.48E-05 | 4.012646 | Higher in NT |
| CFAP298 | -0.25654 | 2.674172 | -5.05069 | 1.92E-06 | 1.52E-05 | 3.984477 | Higher in NT |

| | | | | | | | |
|---|---|---|---|---|---|---|---|
| APRT | -0.64817 | 3.765548 | -5.0339 | 2.05E-06 | 1.62E-05 | 3.916197 | Higher in NT |
| FLNA | -0.5216 | 3.089036 | -5.03283 | 2.06E-06 | 1.62E-05 | 3.911866 | Higher in NT |
| ACOX1 | -0.60287 | 4.326953 | -5.02364 | 2.14E-06 | 1.68E-05 | 3.874556 | Higher in NT |
| ATOX1 | -0.30196 | 2.975046 | -4.99716 | 2.4E-06 | 1.86E-05 | 3.76731 | Higher in NT |
| SDHB | -0.19923 | 3.201088 | -4.98135 | 2.56E-06 | 1.98E-05 | 3.70342 | Higher in NT |
| STX6 | -0.13732 | 3.328042 | -4.96208 | 2.77E-06 | 2.12E-05 | 3.625729 | Higher in NT |
| STAT1 | -0.41765 | 4.474648 | -4.95079 | 2.91E-06 | 2.21E-05 | 3.580312 | Higher in NT |
| SHC1 | -0.36626 | 3.818167 | -4.94846 | 2.93E-06 | 2.22E-05 | 3.570958 | Higher in NT |
| YES1 | -0.20019 | 2.82882 | -4.9274 | 3.2E-06 | 2.41E-05 | 3.486396 | Higher in NT |
| CBL | -0.3631 | 4.121751 | -4.90282 | 3.54E-06 | 2.64E-05 | 3.388017 | Higher in NT |
| EGF.1 | -0.40694 | 3.201238 | -4.88571 | 3.8E-06 | 2.81E-05 | 3.31969 | Higher in NT |
| CAVIN1 | -0.30205 | 3.419554 | -4.88323 | 3.84E-06 | 2.82E-05 | 3.309819 | Higher in NT |
| SDHAF1 | -0.14532 | 3.040777 | -4.87955 | 3.9E-06 | 2.85E-05 | 3.295144 | Higher in NT |
| ERBB4 | -0.12614 | 2.24804 | -4.85794 | 4.26E-06 | 3.1E-05 | 3.209145 | Higher in NT |
| G6PD | -0.59133 | 4.150777 | -4.83425 | 4.7E-06 | 3.39E-05 | 3.115133 | Higher in NT |
| NR0B2 | -0.44436 | 2.316191 | -4.82295 | 4.92E-06 | 3.53E-05 | 3.070415 | Higher in NT |
| TAF15 | -0.26797 | 3.478308 | -4.82095 | 4.96E-06 | 3.54E-05 | 3.062501 | Higher in NT |
| GFER | -0.27684 | 3.069988 | -4.77973 | 5.87E-06 | 4.17E-05 | 2.899915 | Higher in NT |
| BRAF | -0.16507 | 3.140553 | -4.75877 | 6.39E-06 | 4.52E-05 | 2.817559 | Higher in NT |
| PRKCB | -0.50315 | 4.610262 | -4.75066 | 6.6E-06 | 4.65E-05 | 2.785781 | Higher in NT |
| LONP1 | -0.35593 | 2.984055 | -4.71628 | 7.58E-06 | 5.3E-05 | 2.651378 | Higher in NT |
| ALDOA | -0.44772 | 3.171984 | -4.71609 | 7.59E-06 | 5.3E-05 | 2.650638 | Higher in NT |
| CHMP2B | -0.52989 | 4.121215 | -4.70301 | 8E-06 | 5.56E-05 | 2.59966 | Higher in NT |
| GLB1 | -0.38314 | 1.333193 | -4.70009 | 8.1E-06 | 5.6E-05 | 2.588301 | Higher in NT |
| SULT1A1 | -0.48147 | 3.581977 | -4.68698 | 8.53E-06 | 5.87E-05 | 2.537335 | Higher in NT |
| TACO1 | -0.45529 | 3.32825 | -4.68034 | 8.77E-06 | 6E-05 | 2.511547 | Higher in NT |
| PDPK1 | -0.37593 | 2.878096 | -4.66419 | 9.35E-06 | 6.38E-05 | 2.44892 | Higher in NT |
| LDLRAP1 | -0.58032 | 4.368011 | -4.66021 | 9.5E-06 | 6.45E-05 | 2.433511 | Higher in NT |
| STAMBP | -0.25686 | 3.334359 | -4.64813 | 9.97E-06 | 6.71E-05 | 2.386818 | Higher in NT |
| DIABLO | -0.13329 | 3.513386 | -4.63501 | 1.05E-05 | 7.04E-05 | 2.336171 | Higher in NT |
| BAD | -0.34449 | 3.647383 | -4.6116 | 1.15E-05 | 7.63E-05 | 2.246033 | Higher in NT |
| CD68 | -0.1907 | 3.217203 | -4.60468 | 1.19E-05 | 7.8E-05 | 2.219474 | Higher in NT |
| S100A5 | -0.36374 | 3.393892 | -4.58153 | 1.3E-05 | 8.52E-05 | 2.130719 | Higher in NT |
| IL20 | -0.23389 | 2.621989 | -4.5692 | 1.37E-05 | 8.91E-05 | 2.083577 | Higher in NT |
| RAB1B | -0.67392 | 3.600242 | -4.55258 | 1.46E-05 | 9.39E-05 | 2.020144 | Higher in NT |
| VDR | -0.12338 | 2.698912 | -4.54737 | 1.49E-05 | 9.55E-05 | 2.000276 | Higher in NT |
| ITPA | -0.29011 | 3.73467 | -4.52183 | 1.65E-05 | 0.000105 | 1.9032 | Higher in NT |
| DSTN | -0.52522 | 3.629219 | -4.50356 | 1.77E-05 | 0.000112 | 1.833986 | Higher in NT |
| PRKAG1\|P | -0.39901 | 3.514539 | -4.47254 | 2E-05 | 0.000125 | 1.716872 | Higher in NT |
| YWHAH | -0.33832 | 4.847978 | -4.45924 | 2.11E-05 | 0.000131 | 1.666818 | Higher in NT |
| LAMA4 | -0.08931 | 2.805234 | -4.45699 | 2.12E-05 | 0.000132 | 1.658398 | Higher in NT |
| CTNNA1 | -0.37351 | 3.229129 | -4.44975 | 2.18E-05 | 0.000135 | 1.631192 | Higher in NT |
| ITGB5\|ITG | -0.16547 | 3.023994 | -4.42883 | 2.37E-05 | 0.000145 | 1.552779 | Higher in NT |
| FOXO3 | -0.19019 | 2.935498 | -4.4274 | 2.38E-05 | 0.000145 | 1.547434 | Higher in NT |
| FN1.1 | -0.79705 | 4.605993 | -4.4136 | 2.51E-05 | 0.000151 | 1.495868 | Higher in NT |
| CYP3A4 | -0.16713 | 3.581232 | -4.3971 | 2.68E-05 | 0.000161 | 1.434365 | Higher in NT |
| IRAK4 | -0.26197 | 2.824118 | -4.37676 | 2.9E-05 | 0.000173 | 1.358772 | Higher in NT |

| | | | | | | | |
|---|---|---|---|---|---|---|---|
| NISCH | -0.11194 | 3.359292 | -4.3718 | 2.96E-05 | 0.000175 | 1.340339 | Higher in NT |
| SRC | -0.6906 | 3.905691 | -4.35661 | 3.13E-05 | 0.000185 | 1.284106 | Higher in NT |
| SMAD1 | -0.21658 | 3.093871 | -4.34691 | 3.25E-05 | 0.000191 | 1.248238 | Higher in NT |
| PPM1B | -0.28943 | 3.029555 | -4.2917 | 4.02E-05 | 0.000232 | 1.045196 | Higher in NT |
| FN1 | -0.7348 | 3.971223 | -4.27618 | 4.27E-05 | 0.000244 | 0.988417 | Higher in NT |
| TK2 | -0.28725 | 3.047255 | -4.27114 | 4.35E-05 | 0.000248 | 0.970017 | Higher in NT |
| LDHB | -0.3139 | 3.765454 | -4.22771 | 5.13E-05 | 0.00029 | 0.812112 | Higher in NT |
| CXCL2 | -0.35316 | 3.789493 | -4.20864 | 5.51E-05 | 0.00031 | 0.743133 | Higher in NT |
| GPT | -0.21523 | 3.484632 | -4.19433 | 5.82E-05 | 0.000326 | 0.691511 | Higher in NT |
| SMAD2 | -0.3618 | 3.658194 | -4.16984 | 6.38E-05 | 0.000356 | 0.603455 | Higher in NT |
| HSPA8 | -0.1004 | 3.39979 | -4.15129 | 6.83E-05 | 0.00038 | 0.537003 | Higher in NT |
| KLRC4 | -0.18344 | 2.764441 | -4.13998 | 7.13E-05 | 0.000395 | 0.496585 | Higher in NT |
| GSN | -0.14148 | 2.983913 | -4.11023 | 7.97E-05 | 0.000439 | 0.390649 | Higher in NT |
| APLN | -0.09125 | 2.774255 | -4.08114 | 8.87E-05 | 0.000488 | 0.287592 | Higher in NT |
| IFNG | -0.17233 | 2.576031 | -4.07421 | 9.1E-05 | 0.000498 | 0.263106 | Higher in NT |
| HRAS | -0.16381 | 3.183486 | -4.07299 | 9.14E-05 | 0.000499 | 0.258806 | Higher in NT |
| SHC1.1 | -0.27194 | 3.632219 | -4.05705 | 9.7E-05 | 0.000527 | 0.202656 | Higher in NT |
| PTPN11 | -0.30285 | 4.458099 | -4.01702 | 0.000112 | 0.000606 | 0.062302 | Higher in NT |
| VWF | -0.39914 | 4.424643 | -4.00266 | 0.000118 | 0.000637 | 0.012229 | Higher in NT |
| IL21R | -0.14874 | 3.03625 | -3.99039 | 0.000124 | 0.000663 | -0.03047 | Higher in NT |
| IL6 | -0.27846 | 2.761713 | -3.97694 | 0.00013 | 0.000694 | -0.07716 | Higher in NT |
| TRAF1 | -0.13525 | 3.271404 | -3.95379 | 0.000141 | 0.00075 | -0.1573 | Higher in NT |
| CAP1 | -0.52142 | 3.931397 | -3.95059 | 0.000143 | 0.000756 | -0.16833 | Higher in NT |
| PGK1 | -0.15734 | 3.42323 | -3.94557 | 0.000146 | 0.000767 | -0.18565 | Higher in NT |
| SPARC | -0.14584 | 3.537664 | -3.93475 | 0.000152 | 0.000795 | -0.22291 | Higher in NT |
| TPI1 | -0.36544 | 4.125748 | -3.89865 | 0.000173 | 0.000902 | -0.3467 | Higher in NT |
| FOXRED1 | -0.15588 | 2.782511 | -3.88933 | 0.000179 | 0.00093 | -0.37851 | Higher in NT |
| NACA | -0.32087 | 3.877302 | -3.88386 | 0.000182 | 0.000945 | -0.39718 | Higher in NT |
| TXNRD1 | -0.30477 | 3.682587 | -3.88175 | 0.000183 | 0.000949 | -0.40436 | Higher in NT |
| MCL1 | -0.14366 | 2.245333 | -3.8729 | 0.000189 | 0.000976 | -0.43449 | Higher in NT |
| SNAP23 | -0.17385 | 3.617743 | -3.86585 | 0.000194 | 0.000994 | -0.45842 | Higher in NT |
| NPY | -0.1479 | 3.226686 | -3.8573 | 0.0002 | 0.001021 | -0.48744 | Higher in NT |
| RAC1 | -0.46158 | 4.430508 | -3.84639 | 0.000208 | 0.001058 | -0.52439 | Higher in NT |
| CYRIB | -0.30352 | 3.351277 | -3.84347 | 0.00021 | 0.001066 | -0.53426 | Higher in NT |
| GNAI3 | -0.11135 | 2.806511 | -3.83033 | 0.00022 | 0.001113 | -0.57863 | Higher in NT |
| MLX | -0.19011 | 3.05958 | -3.82407 | 0.000225 | 0.001134 | -0.59974 | Higher in NT |
| ALAD | -0.2448 | 4.152251 | -3.8223 | 0.000227 | 0.001138 | -0.60568 | Higher in NT |
| RAC2 | -0.38357 | 4.249445 | -3.80711 | 0.000239 | 0.001197 | -0.65678 | Higher in NT |
| QDPR | -0.28836 | 3.827755 | -3.80315 | 0.000243 | 0.001209 | -0.67004 | Higher in NT |
| PGLS | -0.3403 | 3.527531 | -3.79511 | 0.00025 | 0.00124 | -0.69699 | Higher in NT |
| TKT | -0.28888 | 4.63511 | -3.79163 | 0.000253 | 0.001251 | -0.70865 | Higher in NT |
| GP1BB | -0.29917 | 3.406548 | -3.75602 | 0.000287 | 0.001404 | -0.8274 | Higher in NT |
| NME1 | -0.2206 | 3.389943 | -3.75277 | 0.00029 | 0.001416 | -0.83822 | Higher in NT |
| PTPN1 | -0.21794 | 3.127085 | -3.74266 | 0.0003 | 0.001462 | -0.87174 | Higher in NT |
| ETFA | -0.09498 | 3.286146 | -3.73554 | 0.000308 | 0.001489 | -0.89532 | Higher in NT |
| TMED2 | -0.06138 | 2.726639 | -3.72211 | 0.000323 | 0.001555 | -0.93972 | Higher in NT |
| FASN | -0.26813 | 3.320961 | -3.72135 | 0.000323 | 0.001555 | -0.94222 | Higher in NT |

| | | | | | | | |
|---|---|---|---|---|---|---|---|
| PSMD6 | -0.17937 | 3.288174 | -3.71398 | 0.000332 | 0.00159 | -0.96652 | Higher in NT |
| RAB5A | -0.29972 | 4.019239 | -3.70776 | 0.000339 | 0.00162 | -0.987 | Higher in NT |
| DHFR | -0.206 | 3.167528 | -3.6628 | 0.000396 | 0.001869 | -1.13426 | Higher in NT |
| NFKB1 | -0.28207 | 3.693266 | -3.65703 | 0.000404 | 0.001901 | -1.15305 | Higher in NT |
| SPINT2 | -0.17916 | 2.840678 | -3.62211 | 0.000456 | 0.00213 | -1.26633 | Higher in NT |
| FDX2 | -0.19593 | 2.190642 | -3.6167 | 0.000464 | 0.002163 | -1.28382 | Higher in NT |
| PIK3R1\|PIK | -0.1799 | 2.861984 | -3.60184 | 0.000489 | 0.002262 | -1.33171 | Higher in NT |
| FKBP6 | -0.14414 | 3.346497 | -3.59902 | 0.000493 | 0.002277 | -1.34076 | Higher in NT |
| ERAP1 | -0.30572 | 3.78273 | -3.58776 | 0.000513 | 0.002351 | -1.37694 | Higher in NT |
| HIF1A | -0.11427 | 2.542393 | -3.52477 | 0.000634 | 0.002875 | -1.57766 | Higher in NT |
| CMIP | -0.25958 | 3.281279 | -3.51104 | 0.000664 | 0.003001 | -1.62103 | Higher in NT |
| JAM2 | -0.07955 | 3.674191 | -3.50599 | 0.000676 | 0.003043 | -1.63696 | Higher in NT |
| CBS | -0.1505 | 3.164581 | -3.50071 | 0.000688 | 0.003089 | -1.65359 | Higher in NT |
| C1R | -0.17478 | 4.970684 | -3.49586 | 0.000699 | 0.00313 | -1.66883 | Higher in NT |
| SCO1 | -0.09275 | 3.161843 | -3.49437 | 0.000702 | 0.003136 | -1.67353 | Higher in NT |
| UGT1A1 | -0.13474 | 3.011223 | -3.49019 | 0.000712 | 0.003171 | -1.68663 | Higher in NT |
| SDHAF2 | -0.13211 | 2.4016 | -3.47817 | 0.000742 | 0.003291 | -1.72434 | Higher in NT |
| ACVR1 | -0.24827 | 2.829669 | -3.46895 | 0.000765 | 0.003364 | -1.75319 | Higher in NT |
| IL15 | -0.1411 | 3.050802 | -3.46769 | 0.000768 | 0.003368 | -1.75711 | Higher in NT |
| UQCRB | -0.0665 | 2.41928 | -3.4615 | 0.000784 | 0.003428 | -1.77644 | Higher in NT |
| MAP2K1 | -0.14337 | 2.610006 | -3.43428 | 0.000858 | 0.003719 | -1.86108 | Higher in NT |
| NCF4 | -0.2338 | 2.905013 | -3.4046 | 0.000946 | 0.004089 | -1.95277 | Higher in NT |
| ACADS | -0.09475 | 3.152302 | -3.38831 | 0.000998 | 0.004294 | -2.00281 | Higher in NT |
| PET117 | -0.08358 | 2.745858 | -3.35905 | 0.001098 | 0.004693 | -2.09224 | Higher in NT |
| IRF5 | -0.07385 | 3.115951 | -3.3565 | 0.001107 | 0.004718 | -2.10003 | Higher in NT |
| C3 | -0.12607 | 4.616649 | -3.34082 | 0.001165 | 0.004937 | -2.14764 | Higher in NT |
| HNRNPA1 | -0.36771 | 3.602238 | -3.33859 | 0.001174 | 0.004959 | -2.15441 | Higher in NT |
| STAT5A | -0.1359 | 3.505804 | -3.33464 | 0.001189 | 0.005009 | -2.16638 | Higher in NT |
| MLYCD | -0.13889 | 2.848004 | -3.32532 | 0.001225 | 0.005133 | -2.19458 | Higher in NT |
| FXYD2 | -0.11096 | 3.329407 | -3.3215 | 0.00124 | 0.005183 | -2.20611 | Higher in NT |
| P4HB | -0.13812 | 3.630442 | -3.31908 | 0.00125 | 0.005209 | -2.21342 | Higher in NT |
| GLUL | -0.1389 | 3.571255 | -3.31118 | 0.001282 | 0.005329 | -2.2372 | Higher in NT |
| PPP3R1 | -0.1476 | 3.702422 | -3.30086 | 0.001326 | 0.005478 | -2.26823 | Higher in NT |
| PNP | -0.32686 | 3.792386 | -3.2822 | 0.001408 | 0.005754 | -2.32416 | Higher in NT |
| TAPBP | -0.26722 | 3.546055 | -3.25676 | 0.001527 | 0.006207 | -2.39997 | Higher in NT |
| F8 | -0.20036 | 4.36214 | -3.24541 | 0.001583 | 0.006384 | -2.43363 | Higher in NT |
| COASY | -0.16933 | 3.174863 | -3.22476 | 0.001691 | 0.006762 | -2.49464 | Higher in NT |
| TNFSF4 | -0.11935 | 2.807804 | -3.21322 | 0.001753 | 0.006995 | -2.5286 | Higher in NT |
| PHYH | -0.07744 | 3.319407 | -3.21209 | 0.00176 | 0.007001 | -2.53192 | Higher in NT |
| ITGB2\|ITG | -0.10989 | 2.427398 | -3.20751 | 0.001785 | 0.007085 | -2.54536 | Higher in NT |
| SELPLG.1 | -0.18173 | 2.788877 | -3.19592 | 0.001852 | 0.007329 | -2.57933 | Higher in NT |
| SELP | -0.21957 | 4.414891 | -3.19381 | 0.001864 | 0.007358 | -2.58549 | Higher in NT |
| HK1 | -0.09588 | 3.3632 | -3.17748 | 0.001962 | 0.007665 | -2.63313 | Higher in NT |
| GOSR2 | -0.24594 | 2.040583 | -3.17415 | 0.001983 | 0.007726 | -2.64284 | Higher in NT |
| MAPK9 | -0.15608 | 3.750676 | -3.16524 | 0.002039 | 0.007924 | -2.66873 | Higher in NT |
| DLAT | -0.15531 | 2.872098 | -3.15141 | 0.002129 | 0.00821 | -2.7088 | Higher in NT |
| JUN | -0.0994 | 3.088015 | -3.12609 | 0.002303 | 0.008814 | -2.78175 | Higher in NT |

| | | | | | | | |
|---|---|---|---|---|---|---|---|
| FGB | -0.11807 | 3.177334 | -3.12321 | 0.002324 | 0.008871 | -2.79003 | Higher in NT |
| CXCL3 | -0.32621 | 3.650491 | -3.12225 | 0.002331 | 0.008875 | -2.79279 | Higher in NT |
| ENO3 | -0.24041 | 3.174936 | -3.09811 | 0.002512 | 0.009466 | -2.86183 | Higher in NT |
| ME1 | -0.19329 | 3.161888 | -3.08841 | 0.002588 | 0.009729 | -2.88947 | Higher in NT |
| FADD | -0.11266 | 3.517032 | -3.0239 | 0.003151 | 0.011586 | -3.07134 | Higher in NT |
| GCDH | -0.22511 | 2.92599 | -2.96157 | 0.003802 | 0.013774 | -3.24399 | Higher in NT |
| MANBA | -0.11686 | 3.508866 | -2.9573 | 0.003851 | 0.013884 | -3.25573 | Higher in NT |
| NDUFS4 | -0.28841 | 3.445071 | -2.94145 | 0.004037 | 0.014486 | -3.29909 | Higher in NT |
| PLG.2 | -0.0777 | 3.886528 | -2.9393 | 0.004063 | 0.014515 | -3.30495 | Higher in NT |
| CDC25B | -0.06505 | 3.590485 | -2.88822 | 0.004725 | 0.01654 | -3.44331 | Higher in NT |
| SH3PXD2B | -0.28996 | 3.040492 | -2.88781 | 0.00473 | 0.01654 | -3.44442 | Higher in NT |
| GNS | -0.15641 | 3.065138 | -2.87275 | 0.004944 | 0.017246 | -3.4848 | Higher in NT |
| SCO2 | -0.22257 | 2.730026 | -2.85831 | 0.005157 | 0.017947 | -3.52336 | Higher in NT |
| PNPO | -0.1641 | 3.07524 | -2.8499 | 0.005285 | 0.018307 | -3.54574 | Higher in NT |
| PTHLH | -0.06714 | 2.948819 | -2.83614 | 0.0055 | 0.01901 | -3.58223 | Higher in NT |
| SH2B3 | -0.15529 | 3.049821 | -2.82515 | 0.005678 | 0.019579 | -3.61127 | Higher in NT |
| TMPRSS6 | -0.20427 | 2.938884 | -2.80649 | 0.005992 | 0.020569 | -3.66038 | Higher in NT |
| GPD1L | -0.15522 | 3.717416 | -2.76638 | 0.006722 | 0.022761 | -3.76495 | Higher in NT |
| ENTPD6 | -0.08616 | 2.962009 | -2.76239 | 0.006799 | 0.022969 | -3.77529 | Higher in NT |
| HDAC8 | -0.08814 | 3.918491 | -2.75848 | 0.006875 | 0.023152 | -3.7854 | Higher in NT |
| PLAT | -0.25948 | 3.712416 | -2.74098 | 0.007225 | 0.024084 | -3.83051 | Higher in NT |
| FMO3 | -0.08949 | 3.417668 | -2.73678 | 0.007312 | 0.024211 | -3.84129 | Higher in NT |
| BMPR1B | -0.05858 | 2.560687 | -2.73663 | 0.007315 | 0.024211 | -3.84167 | Higher in NT |
| COL18A1 | -0.11505 | 4.356744 | -2.72369 | 0.007587 | 0.024848 | -3.87482 | Higher in NT |
| ATIC | -0.2662 | 3.582847 | -2.71574 | 0.007759 | 0.025313 | -3.89512 | Higher in NT |
| IL2RG | -0.16898 | 3.571738 | -2.71479 | 0.007779 | 0.025313 | -3.89754 | Higher in NT |
| CCDC103 | -0.10342 | 2.415842 | -2.71134 | 0.007855 | 0.025449 | -3.90633 | Higher in NT |
| CBFB | -0.07552 | 3.084423 | -2.70765 | 0.007937 | 0.025659 | -3.91571 | Higher in NT |
| DNAJC19 | -0.12381 | 2.834245 | -2.70395 | 0.00802 | 0.025871 | -3.92513 | Higher in NT |
| CXCL11 | -0.23075 | 3.24483 | -2.68852 | 0.008374 | 0.026726 | -3.96422 | Higher in NT |
| CD36 | -0.19737 | 4.136693 | -2.68476 | 0.008463 | 0.026951 | -3.97373 | Higher in NT |
| CD27 | -0.07587 | 3.585021 | -2.68259 | 0.008514 | 0.027057 | -3.97921 | Higher in NT |
| LPL | -0.08904 | 3.417124 | -2.67424 | 0.008714 | 0.027578 | -4.00024 | Higher in NT |
| MAD2L2 | -0.08099 | 2.888139 | -2.67135 | 0.008785 | 0.027694 | -4.0075 | Higher in NT |
| ZC3H12A | -0.13498 | 3.034303 | -2.66514 | 0.008938 | 0.028107 | -4.02311 | Higher in NT |
| CD320 | -0.25686 | 3.056351 | -2.65036 | 0.009312 | 0.029221 | -4.06009 | Higher in NT |
| EIF2AK4 | -0.24501 | 2.032603 | -2.6469 | 0.009401 | 0.02944 | -4.06873 | Higher in NT |
| TGFBR1 | -0.16761 | 2.628418 | -2.64039 | 0.009572 | 0.029912 | -4.08494 | Higher in NT |
| BTG1 | -0.08192 | 3.289334 | -2.63389 | 0.009745 | 0.03039 | -4.1011 | Higher in NT |
| COL6A1 | -0.26178 | 4.052282 | -2.61802 | 0.01018 | 0.031483 | -4.1404 | Higher in NT |
| RGS7 | -0.14587 | 2.681849 | -2.61458 | 0.010276 | 0.031651 | -4.1489 | Higher in NT |
| PRAME | -0.10085 | 3.319035 | -2.61318 | 0.010316 | 0.031708 | -4.15236 | Higher in NT |
| PIK3CG | -0.11033 | 3.566944 | -2.60018 | 0.010689 | 0.032655 | -4.18435 | Higher in NT |
| SERAC1 | -0.09206 | 3.120629 | -2.59287 | 0.010904 | 0.033177 | -4.20226 | Higher in NT |
| SULT1E1 | -0.05712 | 3.239245 | -2.57487 | 0.011451 | 0.034701 | -4.24622 | Higher in NT |
| FEV | -0.14143 | 2.865804 | -2.57152 | 0.011556 | 0.034947 | -4.25438 | Higher in NT |
| TYK2 | -0.11796 | 3.441656 | -2.57039 | 0.011591 | 0.034983 | -4.25712 | Higher in NT |

| | | | | | | | |
|---|---|---|---|---|---|---|---|
| GALE | -0.10924 | 2.835491 | -2.56517 | 0.011756 | 0.035338 | -4.26978 | Higher in NT |
| FBLN5 | -0.16576 | 3.831534 | -2.54794 | 0.012315 | 0.036873 | -4.31147 | Higher in NT |
| COMP | -0.06989 | 2.938435 | -2.50133 | 0.013951 | 0.041194 | -4.42299 | Higher in NT |
| MDH1 | -0.17022 | 4.388178 | -2.47551 | 0.014938 | 0.043765 | -4.48396 | Higher in NT |
| STAT5B | -0.05797 | 2.958674 | -2.47212 | 0.015073 | 0.044072 | -4.49193 | Higher in NT |
| STK4 | -0.14931 | 3.611681 | -2.46701 | 0.015277 | 0.044345 | -4.50392 | Higher in NT |
| TSC22D3 | -0.07743 | 2.955573 | -2.46659 | 0.015294 | 0.044345 | -4.5049 | Higher in NT |
| MARCO | -0.09156 | 1.849203 | -2.44993 | 0.015978 | 0.045899 | -4.54384 | Higher in NT |
| GUSB | -0.19193 | 3.528864 | -2.44833 | 0.016045 | 0.04593 | -4.54756 | Higher in NT |
| PDGFB | -0.33245 | 4.442498 | -2.41828 | 0.017353 | 0.049485 | -4.61714 | Higher in NT |

| plot_p |
|---|
| 1.78E-65 |
| 8.02E-54 |
| 1.53E-10 |
| 4.65E-09 |
| 5.81E-09 |
| 9.61E-09 |
| 2.23E-08 |
| 6.2E-08 |
| 4.42E-07 |
| 1.93E-06 |
| 2.12E-06 |
| 3.6E-06 |
| 4.79E-06 |
| 4.79E-06 |
| 7.77E-06 |
| 1.26E-05 |
| 1.27E-05 |
| 1.99E-05 |
| 2.44E-05 |
| 2.7E-05 |
| 3.37E-05 |
| 6.71E-05 |
| 7.2E-05 |
| 7.6E-05 |
| 8.98E-05 |
| 8.98E-05 |
| 9.71E-05 |
| 0.000112 |
| 0.000126 |
| 0.000138 |
| 0.000143 |
| 0.000149 |
| 0.000161 |
| 0.000193 |
| 0.000221 |
| 0.000223 |
| 0.000232 |
| 0.000243 |
| 0.000271 |
| 0.000302 |
| 0.000555 |
| 0.000722 |
| 0.000987 |
| 0.001281 |
| 0.001281 |
| 0.001476 |

| |
|---|
| 0.001703 |
| 0.001767 |
| 0.001817 |
| 0.001952 |
| 0.002245 |
| 0.002325 |
| 0.002378 |
| 0.002624 |
| 0.002853 |
| 0.003325 |
| 0.003327 |
| 0.003566 |
| 0.003708 |
| 0.004294 |
| 0.004354 |
| 0.004885 |
| 0.005044 |
| 0.005428 |
| 0.005587 |
| 0.005587 |
| 0.005587 |
| 0.006166 |
| 0.006246 |
| 0.006368 |
| 0.006387 |
| 0.006387 |
| 0.007622 |
| 0.007638 |
| 0.007642 |
| 0.00802 |
| 0.008059 |
| 0.008333 |
| 0.008602 |
| 0.009136 |
| 0.009136 |
| 0.009281 |
| 0.009944 |
| 0.009951 |
| 0.010424 |
| 0.010492 |
| 0.010544 |
| 0.011298 |
| 0.011299 |
| 0.011529 |
| 0.011973 |
| 0.012099 |
| 0.012221 |

| |
|---|
| 0.012632 |
| 0.01314 |
| 0.013855 |
| 0.014341 |
| 0.014515 |
| 0.014784 |
| 0.014933 |
| 0.015139 |
| 0.015139 |
| 0.01567 |
| 0.015942 |
| 0.015942 |
| 0.018221 |
| 0.019661 |
| 0.020569 |
| 0.020688 |
| 0.021459 |
| 0.021459 |
| 0.022531 |
| 0.023152 |
| 0.023796 |
| 0.023988 |
| 0.024084 |
| 0.024211 |
| 0.024211 |
| 0.024231 |
| 0.024376 |
| 0.024591 |
| 0.025313 |
| 0.025449 |
| 0.02613 |
| 0.026404 |
| 0.026446 |
| 0.026446 |
| 0.027339 |
| 0.027694 |
| 0.030524 |
| 0.030582 |
| 0.030998 |
| 0.03152 |
| 0.032278 |
| 0.032296 |
| 0.03307 |
| 0.034685 |
| 0.035079 |
| 0.036557 |
| 0.037455 |

| |
|---|
| 0.038483 |
| 0.039004 |
| 0.040383 |
| 0.040383 |
| 0.040383 |
| 0.042341 |
| 0.04313 |
| 0.043765 |
| 0.044241 |
| 0.044241 |
| 0.044345 |
| 0.044512 |
| 0.04488 |
| 0.045761 |
| 0.045899 |
| 0.047779 |
| 5.14E-55 |
| 2.13E-35 |
| 1.27E-23 |
| 4.31E-20 |
| 9.96E-18 |
| 2.36E-17 |
| 4.9E-17 |
| 9.52E-17 |
| 6.72E-16 |
| 2.86E-14 |
| 2.86E-14 |
| 3.75E-14 |
| 3.75E-14 |
| 1.25E-13 |
| 1.49E-13 |
| 2.88E-13 |
| 3.12E-12 |
| 6.84E-12 |
| 1.02E-11 |
| 1.36E-11 |
| 1.36E-11 |
| 1.75E-11 |
| 3.22E-11 |
| 3.22E-11 |
| 3.22E-11 |
| 3.23E-11 |
| 3.38E-11 |
| 8.38E-11 |
| 1.02E-10 |
| 1.02E-10 |
| 1.02E-10 |

| |
|---|
| 1.15E-10 |
| 1.16E-10 |
| 1.35E-10 |
| 1.54E-10 |
| 1.71E-10 |
| 2.01E-10 |
| 2.35E-10 |
| 3.32E-10 |
| 3.64E-10 |
| 3.79E-10 |
| 3.79E-10 |
| 3.79E-10 |
| 3.84E-10 |
| 4.85E-10 |
| 6.67E-10 |
| 6.95E-10 |
| 7.57E-10 |
| 8.65E-10 |
| 1.02E-09 |
| 1.03E-09 |
| 1.22E-09 |
| 1.28E-09 |
| 1.33E-09 |
| 1.48E-09 |
| 1.57E-09 |
| 1.66E-09 |
| 2.06E-09 |
| 2.31E-09 |
| 2.36E-09 |
| 2.77E-09 |
| 2.87E-09 |
| 3.72E-09 |
| 4.4E-09 |
| 7.29E-09 |
| 9.63E-09 |
| 9.63E-09 |
| 9.81E-09 |
| 1.02E-08 |
| 1.02E-08 |
| 1.24E-08 |
| 1.36E-08 |
| 1.54E-08 |
| 1.54E-08 |
| 1.64E-08 |
| 2.08E-08 |
| 2.39E-08 |
| 2.49E-08 |

| |
|---|
| 2.98E-08 |
| 3.09E-08 |
| 3.17E-08 |
| 3.29E-08 |
| 3.83E-08 |
| 3.85E-08 |
| 4.12E-08 |
| 4.12E-08 |
| 4.94E-08 |
| 5.79E-08 |
| 5.95E-08 |
| 6.2E-08 |
| 6.37E-08 |
| 7.62E-08 |
| 8.5E-08 |
| 8.5E-08 |
| 8.5E-08 |
| 9.08E-08 |
| 9.55E-08 |
| 9.61E-08 |
| 1.04E-07 |
| 1.07E-07 |
| 1.07E-07 |
| 1.17E-07 |
| 1.23E-07 |
| 1.62E-07 |
| 1.77E-07 |
| 1.78E-07 |
| 2.29E-07 |
| 2.38E-07 |
| 2.5E-07 |
| 2.76E-07 |
| 4.2E-07 |
| 4.37E-07 |
| 5.05E-07 |
| 5.92E-07 |
| 6.43E-07 |
| 6.72E-07 |
| 6.72E-07 |
| 6.89E-07 |
| 8.74E-07 |
| 8.78E-07 |
| 9.66E-07 |
| 1E-06 |
| 1.02E-06 |
| 1.11E-06 |
| 1.25E-06 |

| |
|---|
| 1.47E-06 |
| 1.5E-06 |
| 1.6E-06 |
| 1.69E-06 |
| 1.76E-06 |
| 2.01E-06 |
| 2.11E-06 |
| 2.14E-06 |
| 2.43E-06 |
| 2.57E-06 |
| 3.1E-06 |
| 3.45E-06 |
| 3.66E-06 |
| 3.86E-06 |
| 3.95E-06 |
| 4.38E-06 |
| 4.58E-06 |
| 4.58E-06 |
| 4.61E-06 |
| 4.75E-06 |
| 4.75E-06 |
| 4.75E-06 |
| 4.95E-06 |
| 5.07E-06 |
| 5.81E-06 |
| 5.81E-06 |
| 5.82E-06 |
| 6.5E-06 |
| 6.92E-06 |
| 7.33E-06 |
| 7.88E-06 |
| 8.42E-06 |
| 8.44E-06 |
| 8.44E-06 |
| 9.28E-06 |
| 9.4E-06 |
| 9.79E-06 |
| 9.98E-06 |
| 1.05E-05 |
| 1.06E-05 |
| 1.12E-05 |
| 1.13E-05 |
| 1.21E-05 |
| 1.41E-05 |
| 1.43E-05 |
| 1.48E-05 |
| 1.52E-05 |

| |
|---|
| 1.62E-05 |
| 1.62E-05 |
| 1.68E-05 |
| 1.86E-05 |
| 1.98E-05 |
| 2.12E-05 |
| 2.21E-05 |
| 2.22E-05 |
| 2.41E-05 |
| 2.64E-05 |
| 2.81E-05 |
| 2.82E-05 |
| 2.85E-05 |
| 3.1E-05 |
| 3.39E-05 |
| 3.53E-05 |
| 3.54E-05 |
| 4.17E-05 |
| 4.52E-05 |
| 4.65E-05 |
| 5.3E-05 |
| 5.3E-05 |
| 5.56E-05 |
| 5.6E-05 |
| 5.87E-05 |
| 6E-05 |
| 6.38E-05 |
| 6.45E-05 |
| 6.71E-05 |
| 7.04E-05 |
| 7.63E-05 |
| 7.8E-05 |
| 8.52E-05 |
| 8.91E-05 |
| 9.39E-05 |
| 9.55E-05 |
| 0.000105 |
| 0.000112 |
| 0.000125 |
| 0.000131 |
| 0.000132 |
| 0.000135 |
| 0.000145 |
| 0.000145 |
| 0.000151 |
| 0.000161 |
| 0.000173 |

0.000175
0.000185
0.000191
0.000232
0.000244
0.000248
0.00029
0.00031
0.000326
0.000356
0.00038
0.000395
0.000439
0.000488
0.000498
0.000499
0.000527
0.000606
0.000637
0.000663
0.000694
0.00075
0.000756
0.000767
0.000795
0.000902
0.00093
0.000945
0.000949
0.000976
0.000994
0.001021
0.001058
0.001066
0.001113
0.001134
0.001138
0.001197
0.001209
0.00124
0.001251
0.001404
0.001416
0.001462
0.001489
0.001555
0.001555

| |
|---|
| 0.00159 |
| 0.00162 |
| 0.001869 |
| 0.001901 |
| 0.00213 |
| 0.002163 |
| 0.002262 |
| 0.002277 |
| 0.002351 |
| 0.002875 |
| 0.003001 |
| 0.003043 |
| 0.003089 |
| 0.00313 |
| 0.003136 |
| 0.003171 |
| 0.003291 |
| 0.003364 |
| 0.003368 |
| 0.003428 |
| 0.003719 |
| 0.004089 |
| 0.004294 |
| 0.004693 |
| 0.004718 |
| 0.004937 |
| 0.004959 |
| 0.005009 |
| 0.005133 |
| 0.005183 |
| 0.005209 |
| 0.005329 |
| 0.005478 |
| 0.005754 |
| 0.006207 |
| 0.006384 |
| 0.006762 |
| 0.006995 |
| 0.007001 |
| 0.007085 |
| 0.007329 |
| 0.007358 |
| 0.007665 |
| 0.007726 |
| 0.007924 |
| 0.00821 |
| 0.008814 |

| |
|---|
| 0.008871 |
| 0.008875 |
| 0.009466 |
| 0.009729 |
| 0.011586 |
| 0.013774 |
| 0.013884 |
| 0.014486 |
| 0.014515 |
| 0.01654 |
| 0.01654 |
| 0.017246 |
| 0.017947 |
| 0.018307 |
| 0.01901 |
| 0.019579 |
| 0.020569 |
| 0.022761 |
| 0.022969 |
| 0.023152 |
| 0.024084 |
| 0.024211 |
| 0.024211 |
| 0.024848 |
| 0.025313 |
| 0.025313 |
| 0.025449 |
| 0.025659 |
| 0.025871 |
| 0.026726 |
| 0.026951 |
| 0.027057 |
| 0.027578 |
| 0.027694 |
| 0.028107 |
| 0.029221 |
| 0.02944 |
| 0.029912 |
| 0.03039 |
| 0.031483 |
| 0.031651 |
| 0.031708 |
| 0.032655 |
| 0.033177 |
| 0.034701 |
| 0.034947 |
| 0.034983 |

| |
|---|
| 0.035338 |
| 0.036873 |
| 0.041194 |
| 0.043765 |
| 0.044072 |
| 0.044345 |
| 0.044345 |
| 0.045899 |
| 0.04593 |
| 0.049485 |
