## Supplemental Table 4 for "The Evolution of Primary Aldosteronism and the Role of Norrin"

| Protein | DST Replication | | ACTHstim I |
|---|---|---|---|
| | Slope | p-value | Slope |
| BPGM | 0.125 | 0.016 | - |
| HBB\|HBA1 | 0.187 | 0.038 | - |
| PKLR | 0.077 | 0.197 | - |
| GLO1 | - | - | - |
| S100A6 | - | - | - |
| HMBS | - | - | - |
| CA1 | 0.081 | 0.082 | - |
| CA3 | 0.056 | 0.157 | - |
| HPX | 0.059 | 0.18 | 0.076 |
| NDP | 0.04 | 0.027 | 0.046 |
| NCF1 | 0.012 | 0.86 | - |
| PI3 | 0.014 | 0.644 | 0.035 |
| COMT | - | - | - |
| SURF1 | 0.004 | 0.889 | 0.031 |
| GAA | - | - | 0.033 |
| TFF3 | - | - | 0.02 |
| CKM | 0.069 | 0.061 | 0.064 |
| CFHR5 | - | - | 0.04 |
| MMP9 | - | - | - |
| STX3 | - | - | 0.041 |
| UROS | 0.051 | 0.163 | - |
| FBP2 | - | - | - |
| PRDX1 | - | - | - |
| ITGB1\|ITGAV | 0.038 | 0.231 | 0.076 |
| CCL3 | 0.011 | 0.389 | 0.007 |
| GH1 | 0.022 | 0.322 | - |
| CHD7 | - | - | 0.021 |
| MGAT2 | - | - | 0.008 |
| RPIA | - | - | - |
| RPS6KB1 | 0.03 | 0.153 | - |
| TNFRSF4 | - | - | - |
| PNPLA2 | - | - | - |
| RGS5 | - | - | -0.017 |
| CCL2 | - | - | - |
| CDON | - | - | 0.019 |
| DTNBP1 | 0.002 | 0.742 | - |
| HJV | - | - | 0.006 |
| AMBP | - | - | 0.014 |
| ODAD4 | 0.018 | 0.134 | 0.016 |
| F10 | - | - | - |
| CD38 | - | - | - |
| TAT | - | - | 0.015 |
| RXRA | - | - | - |
| BMPR1B | -0.01 | 0.265 | - |
| TBX5 | - | - | - |
| ACADS | -0.007 | 0.609 | - |
| APOA4 | - | - | - |
| HSPA8 | - | - | - |

| | | | |
|---|---|---|---|
| GPX3 | - | - | - |
| CYCS | -0.009 | 0.702 | 0.013 |
| MAD2L2 | -0.021 | 0.302 | - |
| ETFA | -0.021 | 0.191 | - |
| CBFB | -0.015 | 0.234 | - |
| ANGPTL3 | -0.042 | 0.057 | -0.026 |
| DSG2 | -0.023 | 0.169 | -0.032 |
| EPHX2 | -0.006 | 0.717 | - |
| IRF5 | -0.013 | 0.355 | - |
| VEGFA.1 | - | - | - |
| BIRC3 | -0.022 | 0.161 | - |
| GCG | -0.023 | 0.425 | - |
| PTK2 | -0.051 | 0.22 | -0.048 |
| GNAI3 | - | - | - |
| ITGB1\|ITGA1 | - | - | - |
| STX6 | -0.016 | 0.365 | - |
| ACVR1 | -0.027 | 0.228 | - |
| LHB\|CGA | -0.07 | 0.076 | -0.061 |
| ICOSLG | - | - | - |
| UGT1A1 | -0.035 | 0.172 | - |
| DUSP6 | -0.027 | 0.064 | - |
| CD36 | - | - | -0.062 |
| ID1 | -0.049 | 0.026 | -0.032 |
| ITGB5\|ITGAV | - | - | -0.012 |
| CD68 | - | - | - |
| COL6A1 | - | - | -0.062 |
| KRAS | - | - | - |
| HSPA1A | -0.03 | 0.281 | - |
| TALDO1 | - | - | -0.011 |
| IL18 | - | - | - |
| SEPTIN11 | - | - | - |
| CTSC | -0.031 | 0.358 | - |
| RASA1 | - | - | - |
| GLB1 | - | - | -0.048 |

*Footnote: Proteins that did not exhibit a consistent trend upwards or downwards within a re*

| ɛplication | Upright Sodium Restricted Replication | |
|---|---|---|
| p-value | Slope | p-value |
| - | 0.043 | 0.159 |
| - | 0.057 | 0.152 |
| - | 0.036 | 0.252 |
| - | 0.052 | 0.039 |
| - | 0.047 | 0.057 |
| - | - | - |
| - | 0.03 | 0.349 |
| - | 0.007 | 0.749 |
| 0.049 | - | - |
| 0.011 | 0.034 | 0.071 |
| - | - | - |
| 0.161 | - | - |
| - | - | - |
| 0.16 | 0.019 | 0.396 |
| 0.107 | 0.013 | 0.474 |
| 0.535 | 0.008 | 0.814 |
| 0.076 | 0.034 | 0.261 |
| 0.014 | 0.025 | 0.14 |
| - | 0.051 | 0.219 |
| 0.268 | - | - |
| - | 0.038 | 0.05 |
| - | - | - |
| - | - | - |
| 0.005 | 0.053 | 0.044 |
| 0.614 | - | - |
| - | 0.012 | 0.375 |
| 0.097 | -0.005 | 0.662 |
| 0.453 | - | - |
| - | 0.015 | 0.355 |
| - | 0.041 | 0.08 |
| - | - | - |
| - | - | - |
| 0.249 | - | - |
| - | - | - |
| 0.174 | - | - |
| - | - | - |
| 0.558 | 0.014 | 0.163 |
| 0.057 | 0.016 | 0.032 |
| 0.129 | - | - |
| - | 0.026 | 0.004 |
| - | - | - |
| 0.124 | - | - |
| - | - | - |
| - | -0.01 | 0.353 |
| - | - | - |
| - | - | - |
| - | -0.022 | 0.052 |
| - | - | - |

| | | |
|---|---|---|
| - | - | - |
| 0.553 | - | - |
| - | - | - |
| - | -0.017 | 0.096 |
| - | -0.009 | 0.416 |
| 0.084 | - | - |
| 0.027 | -0.012 | 0.456 |
| - | -0.011 | 0.25 |
| - | -0.009 | 0.363 |
| - | -0.035 | 0.19 |
| - | - | - |
| - | - | - |
| 0.237 | -0.046 | 0.274 |
| - | 0.007 | 0.658 |
| - | 0.047 | 0.276 |
| - | - | - |
| - | -0.043 | 0.054 |
| 0.118 | - | - |
| - | - | - |
| - | -0.022 | 0.381 |
| - | -0.003 | 0.772 |
| 0.092 | - | - |
| 0.205 | -0.05 | 0.037 |
| 0.553 | - | - |
| - | - | - |
| 0.223 | - | - |
| - | 0.018 | 0.425 |
| - | - | - |
| 0.667 | - | - |
| - | - | - |
| - | - | - |
| - | - | - |
| - | 0.031 | 0.266 |
| 0.218 | - | - |

*plication condition are shown as '-'.*
